# Human genetic risk for major depressive disorder implicates motor cortex PVALB⁺ inhibitory neurons

**DOI:** 10.64898/2026.09.08.26362568

**Authors:** Yunlong Ma, Haijun Han, Cheng Chen, Xiaoqin Huang, Cheng Gao, Wei Dai, Fei Qiu, Jingjing Li, Yijun Zhou, Dingping Jiang, Gongwei Zheng, Zhengbiao Zhu, Chunyu Deng, Yizhou Huang, Jianhong Zhou, Connor Jops, Jade England, Daniel Vo, Xiang Cai, Michael J. Gandal, Jianzhong Su

## Abstract

Major depressive disorder (MDD) is highly polygenic, but the cellular and anatomical contexts through which inherited risk contributes to disease biology remain incompletely understood. Here we integrated large-scale MDD genome-wide association studies with an adult human brain single-cell discovery atlas and five independent replication atlases spanning brain regions, developmental stages, species, and disease states, comprising more than five million cells and nuclei. Across complementary polygenic mapping frameworks, MDD genetic signals were preferentially enriched in neuronal populations and consistently prioritized the primary motor cortex (M1) as a key anatomical context of risk. Subtype-level analyses highlighted PVALB⁺ inhibitory neurons and Ex-L2/4 excitatory neurons, with integrative gene prioritization nominating *CNNM2* as a leading PVALB⁺-linked MDD risk-gene candidate. *CNNM2*⁺ PVALB⁺ inhibitory neurons showed elevated genetic risk scores and were enriched for synaptic, membrane-potential, and activity-related programs. In chronic-stress mouse models, Cnnm2 protein levels were reduced in M1 PV⁺ and SST⁺ inhibitory neurons, and optogenetic activation of M1 PV⁺ neurons rescued stress-induced depression-like behaviours. Anatomical tracing and activity mapping further implicated an M1–l/vlPAG circuit axis in exercise-associated antidepressant-like effects. These findings connect MDD polygenic risk to motor-cortical inhibitory-neuron biology and experimentally tractable circuit mechanisms and provide scDepBrain as a resource for exploring MDD-associated cellular programs across the brain.

## INTRODUCTION

Major depressive disorder (MDD) affects more than 300 million people worldwide^1^ and is the leading contributor to non-fatal health loss globally^2^. Twin studies estimate the heritability of MDD to be approximately 40%^3^, and recent large-scale genome-wide association studies (GWAS) have shown that this liability is highly polygenic, involving hundreds of common variants of small effect^4–11^. The latest PGC trans-ancestry depression GWAS has substantially expanded the genetic landscape of MDD and prioritized 308 high-confidence risk genes through fine-mapping and functional genomic analyses^12^. A central challenge is to move from these statistical associations to mechanistic insight into disease biology^13–18^. Because most MDD risk variants reside in non-coding regions, their effects are expected to depend on the cellular, anatomical, circuit and developmental contexts in which regulatory elements are active^19–21^. Resolving these contexts is essential for defining the cellular etiology of MDD and for linking polygenic risk to disease-relevant molecular programs and neural circuits.

Single-cell RNA sequencing provides an unprecedented opportunity to resolve the cellular composition, transcriptional states and disease-associated molecular programs of the human brain at cellular resolution^20,22–27^. In MDD, however, existing single-cell studies^28–31^ have remained limited in anatomical breadth, cohort diversity, and genetic interpretability. snRNA-seq analysis of the dorsolateral prefrontal cortex implicated deep-layer excitatory neurons and oligodendrocyte progenitor cells^30^; subsequent work refined sex- and gene-specific transcriptional effects^31^; and macaque spatial transcriptomics nominated a depression-associated microglial subtype^28^. Although these studies have revealed important region-specific transcriptional alterations, their relationship to the broader cellular architecture of inherited risk remains unresolved. Thus, neither anatomically restricted single-cell studies nor GWAS alone can localize genetic risk to its relevant cellular and anatomical contexts across the human brain. Integrating these complementary data types provides a route to a genetically informed, brain-wide map of the cell populations and anatomical contexts associated with MDD risk.

Computational approaches^32–36^ have increasingly enabled the integration of GWAS summary statistics with cell-type-resolved transcriptomic maps. Early studies^37,38^ leveraged bulk tissue expression or FACS-isolated cell populations to nominate disease-relevant tissues and broad cellular classes, whereas subsequent frameworks^39–44^, including our recently developed scPagwas^39^ and complementary methods such as scDRS^44^, extended this strategy to single-cell and single-nucleus datasets. Applied to neuropsychiatric disorders, including MDD, these approaches have so far been used predominantly for cell-type-level enrichment, nominating broad neuronal populations across mouse and human brain datasets^12,45–51^. However, their capacity to resolve trait-relevant cells at single-cell resolution—and to detect risk-enriched cellular states that transcend predefined annotations—has not yet been systematically exploited for MDD. More broadly, most applications have relied on selected single-cell datasets, predefined cell-type labels, or individual analytical frameworks. As a result, the regional, developmental, and species-conserved cellular architecture underlying MDD genetic risk remains only partially resolved. Moreover, computationally prioritized cell populations have rarely been linked to candidate genes, anatomical projections, or experimentally tractable circuits. Closing this gap requires a unified framework that moves from genetically informed cellular discovery to molecular and circuit-level validation in disease-relevant models.

In this context, the motor cortex represents an underexplored but biologically plausible site for circuit-level investigation. Although the primary motor cortex (M1/M1C) is classically viewed as an output node for voluntary movement, it also participates in sensorimotor integration, action selection and motivational control^52,53^. M1 modulation has been explored primarily for pain relief^54–56^, with emerging evidence that motor-cortical neuromodulation may also influence depressive symptoms^57–59^. In parallel, physical exercise has robust antidepressant and anxiolytic effects^60^, yet the cortical circuits linking motor activity to mood regulation remain poorly understood. The lateral and ventrolateral periaqueductal grey (l/vlPAG), which has established roles in pain, defensive responses, anxiety and depression-like behaviours^61–63^, provides a plausible downstream node for linking motor-cortical activity to affective regulation. These observations motivated us to test whether M1-to-l/vlPAG circuits regulate depression-like behaviour and contribute to the antidepressant effects of exercise.

Here, we map the cellular architecture of MDD risk by developing a curated adult human brain single-cell and single-nucleus transcriptomic discovery atlas and integrating it with three large-scale MDD GWAS datasets spanning European-ancestry and trans-ancestry populations^5,12^, followed by validation across independent brain atlases comprising more than five million cells and nuclei. Using complementary polygenic-scoring frameworks operating at cell-type and single-cell resolution^39–41,44^, we implicate neuronal populations within M1 as key contexts of MDD polygenic burden, with subtype-level analyses highlighting PVALB⁺ inhibitory neurons and Ex-L2/4 excitatory neurons. We then integrated gene-level association, transcriptome-wide association, and cell-state-specific expression evidence to prioritize *CNNM2* as a leading PVALB⁺-linked MDD risk-gene candidate and identify a *CNNM2*-centered inhibitory-neuron state in M1. In chronically stressed mice, Cnnm2 protein levels were reduced in M1 PV⁺/SST⁺ inhibitory neurons. Combining viral tracing, activity mapping and optogenetic perturbation, we further link M1 PV⁺ neurons and the M1 → l/vlPAG pathway to stress-induced depression-like behaviour and exercise-associated antidepressant effects. Together, these findings connect human MDD polygenic risk to disease-relevant cells, genes and circuits, and establish scDepBrain (single-cell Depression Brain Map; https://scdepbrain.su-lab.org) as an interactive platform for exploring MDD-associated cellular programs across the brain.

## RESULTS

### scDepBrain integrates multi-context brain single-cell atlases for MDD genetic mapping

We designed an integrative discovery-to-validation framework spanning atlas construction, genetically informed cellular and gene prioritization, experimental validation, and community resource development (**Fig. 1**). First, we curated and harmonized large-scale brain single-cell and single-nucleus transcriptomic datasets into an adult human discovery atlas and five independent replication datasets spanning adult human, developing human, mouse, cerebral organoid and human MDD case-control brain samples (**Fig. 1a,b**). Second, we integrated the discovery atlas with three large-scale MDD GWAS resources, including the Howard et al. European-ancestry GWAS^5^ and the updated PGC European-ancestry and trans-ancestry depression GWAS datasets^12^, to prioritize MDD-relevant cell types, cell states and candidate genes and to assess robustness across GWAS sample size and ancestry composition. These genetic resources were analyzed through complementary polygenic-scoring frameworks operating at both cell-type and single-cell resolution, together with gene-level association, transcriptome-wide association and eQTL-informed gene-prioritization analyses (**Fig. 1c,d**). The replication single-cell datasets were then used to evaluate the robustness, developmental timing, cross-species conservation and disease-state relevance of the prioritized cell populations and risk genes. Third, we experimentally tested the prioritized molecular and anatomical findings through immunofluorescence profiling, viral tracing, and circuit-level perturbations in chronic-stress mouse models (**Fig. 1e**). Finally, we made the resulting MDD-relevant brain cellular maps accessible through scDepBrain, a public resource designed to support exploration and reuse by the community (**Fig. 1f**).

**Figure 1.**
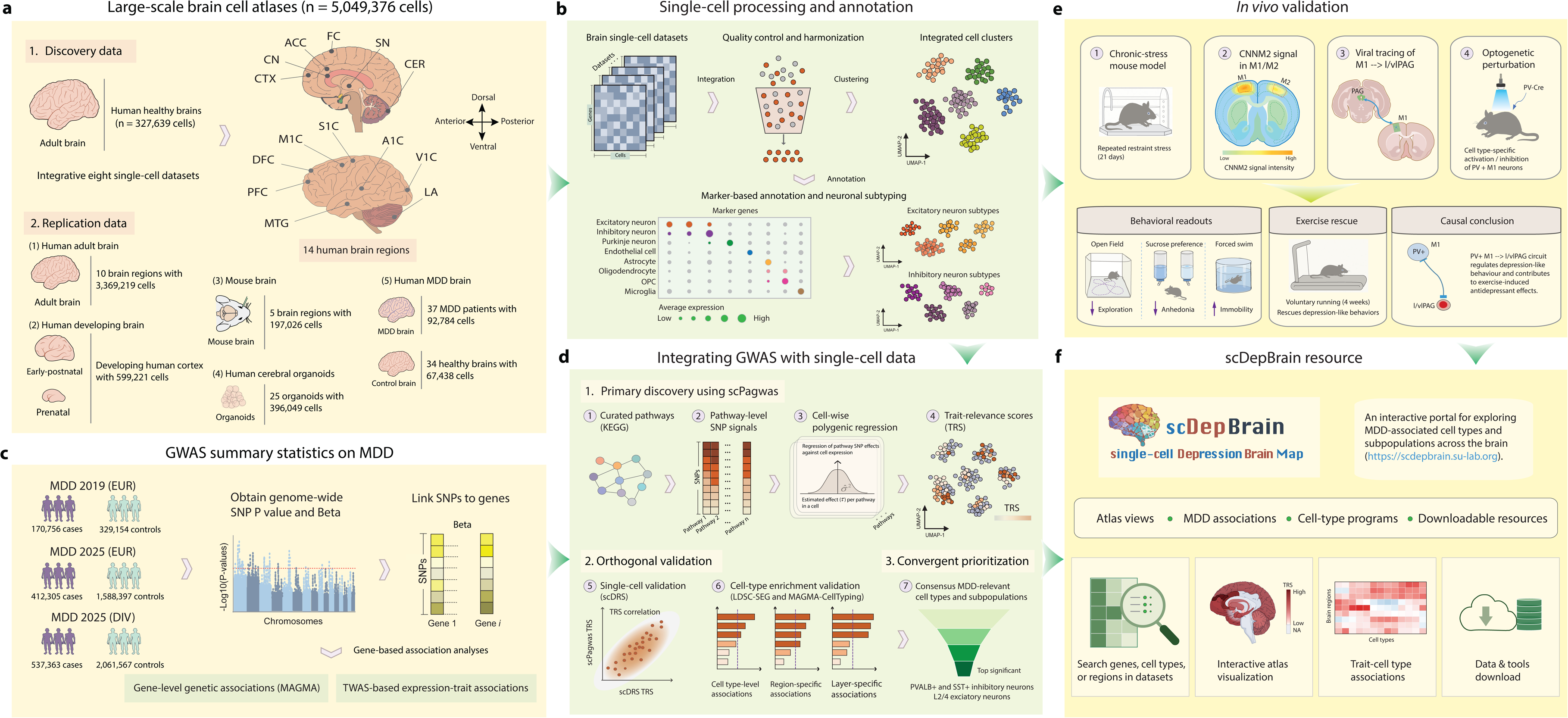
Integrative genomics framework for mapping MDD genetic risk to brain cell types, molecular programs and circuits. **a.** Overview of the single-cell and single-nucleus brain atlases used in this study. The discovery atlas comprised eight adult human brain datasets spanning 14 anatomical regions and 327,639 cells or nuclei. Five independent replication atlases were assembled across the adult human brain, developing human brain, mouse brain, human cerebral organoids and human MDD case–control brain tissue, together comprising 5,049,376 cells and nuclei. **b.** Single-cell processing and annotation workflow. Public datasets were harmonized through quality control, integration, batch-effect correction, clustering and marker-based cell-type annotation, followed by neuronal subtype annotation of excitatory and inhibitory populations. **c.** GWAS summary statistics for MDD were integrated with transcriptomic data. SNP-level association statistics from three large-scale MDD GWASs were mapped to genes and used for gene-based association analysis and transcriptome-wide association analysis. **d.** Genetic prioritization strategy. scPagwas was used as the primary discovery framework to integrate MDD GWAS with single-cell transcriptomes and compute single-cell-level and cell-type-level trait-relevance scores (TRSs). Orthogonal methods, including scDRS, LDSC-SEG and MAGMA-CellTyping, were used for validation at single-cell, cell-type, regional and layer-specific levels. Convergent prioritization nominated MDD-relevant neuronal populations, anatomical contexts and candidate risk genes. **e.** In vivo validation strategy. Computationally prioritized molecular and anatomical findings were tested in chronic-stress mouse models using behavioural assays, immunofluorescence profiling of CNNM2 in motor-cortical inhibitory neurons, viral tracing of M1→l/vlPAG projections and cell-type-specific optogenetic perturbation. **f.** scDepBrain resource. Processed atlases, MDD association results, cell-type programs and downloadable resources were integrated into scDepBrain, an interactive platform for exploring MDD-associated cellular programs across brain contexts.

For discovery, we assembled eight published healthy adult human brain snRNAseq and, after standardized preprocessing and stringent quality control, retained 327,639 high-quality nuclei across 14 anatomical regions (**Fig. 1a,b**; **Supplementary Table 1** and **Supplementary Fig. 1**). Uniform integration and clustering of the discovery atlas resolved 29 clusters (**Supplementary Fig. 2**), which were annotated into eight major brain cell classes—excitatory neurons, inhibitory neurons, Purkinje neurons, astrocytes, oligodendrocytes, OPCs, microglia and endothelial cells—using canonical markers (**Fig. 2a,b** and **Supplementary Fig. 3**). Second-round clustering further resolved excitatory and inhibitory neurons into layer- and subtype-specific populations, including eight excitatory neuronal subtypes and six inhibitory neuronal subtypes, each displaying distinct marker profiles and cortical-layer signatures (**Fig. 2c, d**; **Supplementary Figs. 4–5**).

**Figure 2.**
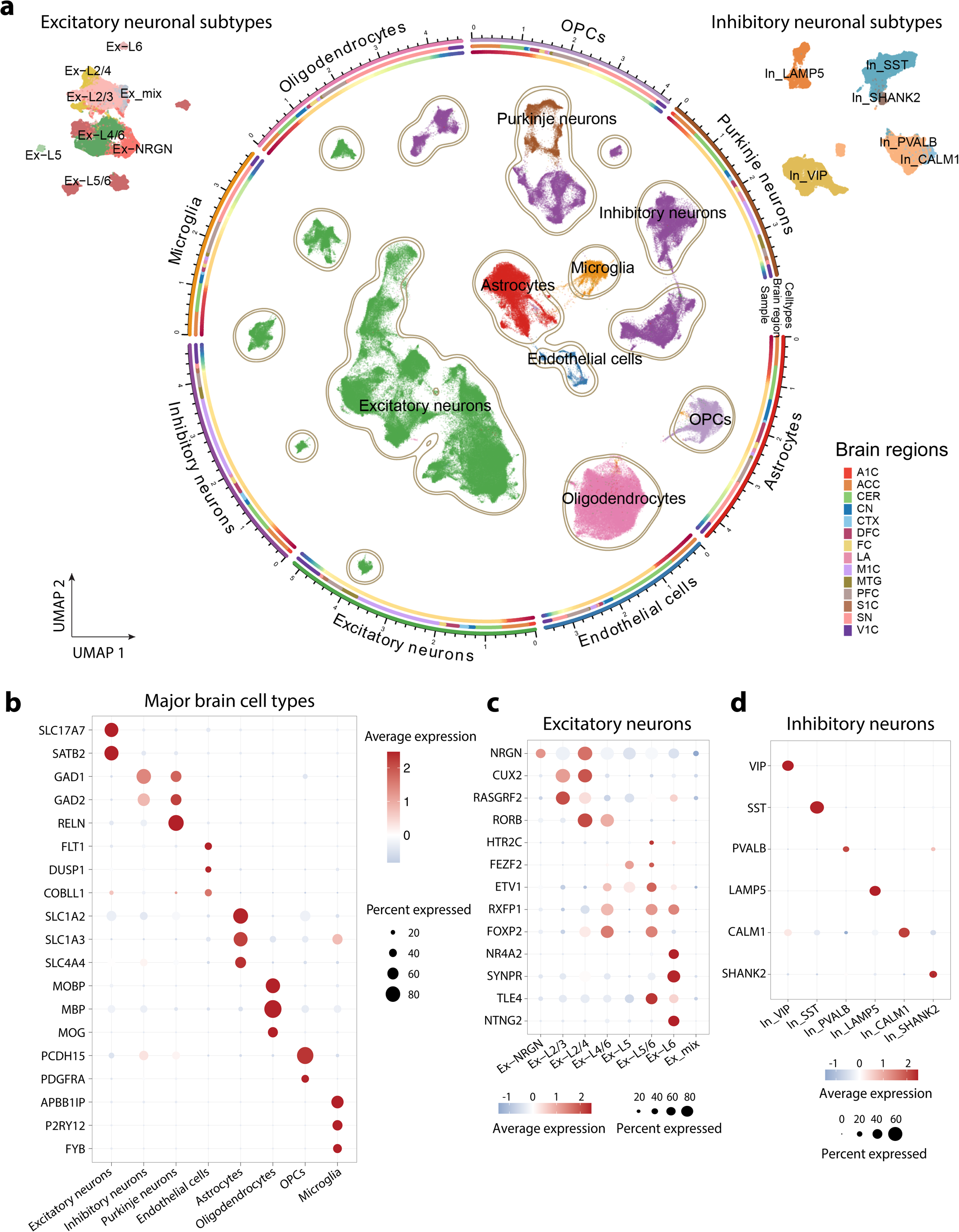
Construction and annotation of the adult human brain single-cell discovery atlas. **a.** Integrated UMAP embedding of the adult human brain discovery atlas showing major brain cell classes and neuronal subtypes (n = 327,639 cells). Cells are coloured by major cell type, with excitatory and inhibitory neuronal subtypes shown separately in the insets. Outer tracks indicate the anatomical brain-region composition of each major cell class. **b.** Dot plot showing expression of canonical marker genes across major brain cell classes, including excitatory neurons, inhibitory neurons, Purkinje neurons, endothelial cells, astrocytes, oligodendrocytes, oligodendrocyte precursor cells (OPCs) and microglia. **c.** Dot plot showing representative marker-gene expression across excitatory neuronal subtypes. **d.** Dot plot showing representative marker-gene expression across inhibitory neuronal subtypes. In b– d, dot size indicates the percentage of cells expressing each gene, and colour indicates scaled average expression.

The annotated neuronal and non-neuronal populations were broadly distributed across brain regions, ages, donors, and source datasets, indicating that batch effects and technical biases were effectively mitigated by the integration strategy (**Supplementary Figs. 6–7**). The integrated atlas preserved expected biological organization across anatomical and developmental axes. Hierarchical clustering of regional cell composition revealed relatedness among anatomically adjacent brain regions, whereas age-stratified analyses across five postnatal periods^66^—childhood, adolescence, young adulthood, middle adulthood and late adulthood—captured dynamic shifts in neuronal and non-neuronal cell proportions (**Supplementary Figs. 8–9**). Collectively, these analyses establish a harmonized, biologically structured adult human brain discovery atlas for subsequent single-cell genetic mapping of MDD risk.

To enable replication and assess generalizability, we further curated five independent single-cell and single-nucleus datasets—adult human brain, developing human brain, mouse brain, human cerebral organoids and human MDD case–control brain samples (**Supplementary Figs. 10–14**). These resources included 3,369,219 adult-human brain cells across 10 regions; 599,221 developing-human brain cells spanning gestational week 6 to postnatal month 8; 197,026 mouse-brain cells across five regions; 396,049 human cerebral organoid cells from 25 organoids; and 160,222 nuclei from 37 MDD patients and 34 controls (**Supplementary Tables 2–3**). In total, the discovery and replication resources comprised 5,049,376 single-cell and single-nucleus transcriptomes from 343 individuals or biological samples, represented across 1,599 region-by-sample profiles and 34 independent studies (**Supplementary Fig. 1**; **Supplementary Table 4**), providing a broad, multi-context reference for downstream genetic analyses.

### MDD genetic risk preferentially maps to neuronal populations

To identify cell populations relevant to MDD, we integrated the adult human brain discovery atlas with three large-scale MDD GWAS resources (**Supplementary Table 5**): the Howard et al. European-ancestry GWAS^5^ and the latest PGC European-ancestry and trans-ancestry depression GWAS datasets^12^. Using the scPagwas polygenic regression framework^39^, the initial Howard et al. analysis prioritized three neuronal populations at the major cell-type level: excitatory neurons (P = 3.35 × 10^-9^), inhibitory neurons (P = 3.14 × 10^-6^) and Purkinje neurons (P = 5.98 × 10^-4^; FDR < 0.05; **Fig. 3a**), whereas non-neuronal cell types showed no significant enrichment (FDR > 0.05; **Fig. 3a** and **Supplementary Table 6**). The larger updated PGC European-ancestry and trans-ancestry GWAS datasets recapitulated this neuronal enrichment, supporting the stability of the cell-type prioritization results across GWAS sample size and ancestry composition (**Supplementary Fig. 15a**).

**Figure 3.**
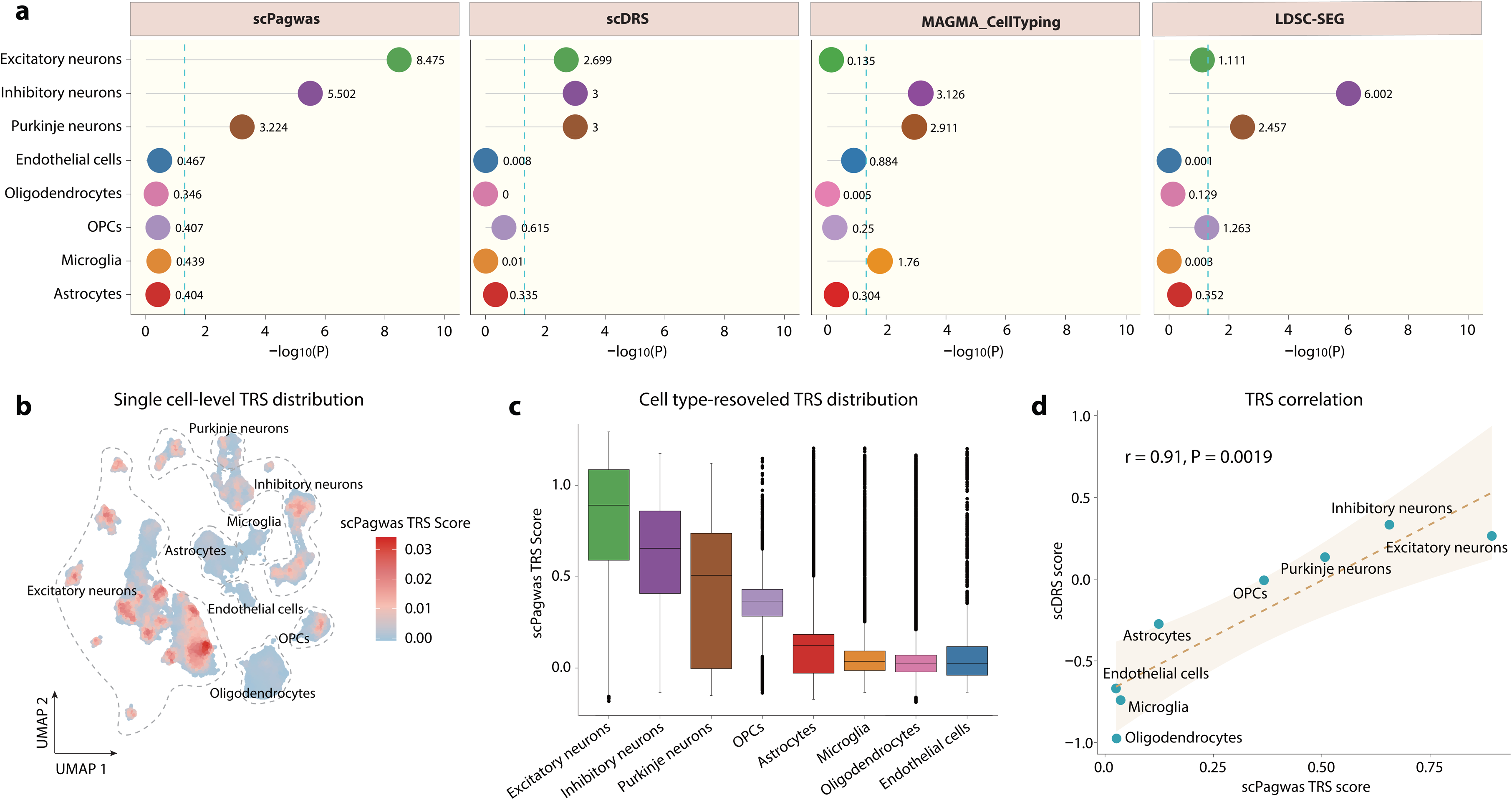
Single-cell genetic frameworks prioritize neuronal populations for MDD genetic risk. **a.** Cell-type-level prioritization of MDD genetic risk in the adult human brain discovery atlas using four complementary frameworks: scPagwas, scDRS, MAGMA-CellTyping and LDSC-SEG. Dot plots show the significance of MDD association across major brain cell types. The x axis shows −log₁₀(P), and the dashed line indicates the nominal significance threshold. Neuronal populations, including excitatory neurons, inhibitory neurons and Purkinje neurons, showed convergent enrichment across methods. **b.** UMAP visualization of single-cell scPagwas trait-relevance scores (TRSs) across the discovery atlas. Red indicates higher MDD trait relevance, and light blue indicates lower trait relevance. **c.** Box plots showing the distribution of scPagwas TRSs across major brain cell types. Neuronal populations showed higher TRSs than non-neuronal cell types. Box plots show median and interquartile range; whiskers indicate 1.5× interquartile range. **d.** Correlation between cell-type-level scPagwas TRSs and scDRS scores across major brain cell types. Each point represents one annotated cell type. The dashed line indicates the fitted linear regression, and the shaded region denotes the confidence interval. Pearson correlation coefficient and *P* value are shown.

To further evaluate this neuronal signal, we applied three complementary approaches—scDRS^44^, LDSC-SEG^41^, and MAGMA-CellTyping^40^—which converged on the same neuron-dominant pattern despite method-specific differences in effect strength (**Fig. 3a**; **Supplementary Fig. 15a** and **Supplementary Table 6**). The 308 high-confidence MDD genes prioritized by the latest PGC depression GWAS also showed elevated module activity in neuronal populations, particularly excitatory and inhibitory neurons, in the adult human brain discovery atlas (**Supplementary Fig. 15b**). As an additional gene-set-based sensitivity analysis, scDRS using this 308-gene set again prioritized neuronal populations, including excitatory, inhibitory and Purkinje neurons (**Supplementary Fig. 15c**). Following previous studies^44,46^, we used standing height as a negative-control trait and observed no enrichment of neuronal populations, supporting the specificity of the MDD-associated signal (**Supplementary Table 7**).

We next assessed the reproducibility of this pattern across independent single-cell datasets. In the adult-human replication dataset, scPagwas prioritized multiple region- and layer-specific neuron populations, including hippocampal CA1–3 neurons (P = 7.66 × 10^-8^), amygdala excitatory neurons (P = 2.77 × 10^-7^), and medium spiny neuron (P = 4.26 × 10^-3^), whereas non-neuronal populations were not significant (FDR > 0.05; **Supplementary Fig. 16** and **Supplementary Table 8**). Consistent neuronal enrichment was also observed in mouse brain and human cerebral organoid datasets (FDR < 0.05; **Supplementary Fig. 17a,b** and **Supplementary Tables 9-10**). By contrast, the developing human brain dataset showed a distinct pattern, with oligodendrocytes and astrocytes showing the strongest enrichment, while excitatory neurons reached only nominal significance (P = 0.038) and inhibitory neurons a suggestive trend (P = 0.15; **Supplementary Fig. 18**; **Supplementary Table 11**). These results suggest that MDD genetic risk maps predominantly to neuronal populations in the adult brain, while developmental datasets may capture additional glial-lineage or maturation-associated programs.

To refine these associations at single-cell resolution, we assigned each cell a scPagwas trait relevance score (TRS; **Methods**). High-TRS cells were concentrated within neuronal populations in the discovery atlas, and cell-type-resolved TRS distributions confirmed higher scores in excitatory, inhibitory and Purkinje neurons than in non-neuronal populations (**Fig. 3b-c**). An independent single-cell-level analysis using scDRS produced a similar neuron-dominant hierarchy (**Supplementary Fig. 19a**), and scPagwas TRSs correlated strongly with scDRS scores across both cell types and individual cells (**Fig. 3d**; **Supplementary Fig. 19b**). Replication datasets showed concordant TRS patterns, with neuronal populations prioritized in adult human, mouse and organoid atlases, and relatively stronger glial signals in the developing human brain (**Supplementary Fig. 20a-e**).

### M1C emerges as a key anatomical context for neuronal MDD risk

Having established that MDD genetic risk preferentially maps to neuronal populations, we next used stratified analyses to identify the anatomical and developmental contexts in which this neuronal signal was most pronounced. In sex-stratified scPagwas analyses, neuronal enrichment was broadly concordant in males and females, indicating that the major neuron-level association was not driven by one sex alone; orthogonal cell-type-level methods yielded consistent patterns (**Supplementary Fig. 21a,b**; **Supplementary Table 12**). Age-stratified analyses showed neuronal enrichment across postnatal periods, with the strongest signals observed in early postnatal and adolescent stages (P1-P2) and persistent, although weaker, neuronal signals in later periods (P3-P5). Complementary methods showed a broadly consistent neuronal-dominant pattern (**Supplementary Fig. 22a–e**; **Supplementary Table 13**). This pattern suggests that MDD genetic liability intersects with neuronal programs across postnatal developments, including developmental windows that precede or overlap the typical age-of-onset for depressive and other mental disorders^67,68^.

Region-stratified scPagwas analyses revealed substantial anatomical heterogeneity in the neuronal MDD genetic signal (**Supplementary Fig. 23**; **Supplementary Table 14**), with consistent evidence recapitulated by complementary methods (**Supplementary Figs. 24-26; Supplementary Tables 15-17**). Single-cell-resolved scPagwas inference identified primary motor cortex (M1C) as the region with the highest TRSs, with additional elevated signals in cortical regions including middle temporal gyrus (MTG), primary somatosensory cortex (S1C) and anterior cingulate cortex (ACC) (**Supplementary Fig. 27a**). To control for regional differences in cell abundance, we repeated the analysis after subsampling an equal number of cells from each region (n = 1,000 cells per region); the M1C-leading pattern was preserved, arguing against confounding by regional cell number (**Supplementary Fig. 27b,c**). Notably, M1C ranked above prefrontal cortical regions, which have been strongly emphasized in prior genetic, neuroimaging and transcriptomic studies of MDD^5,31,69^.

Independent validation using per-cell scDRS scoring further supported this regional pattern, with M1C consistently ranking among the highest-scoring regions in the Howard et al. GWAS^5^ and in both the European-ancestry and trans-ancestry meta-analyses from the latest PGC depression GWAS^12^ (**Supplementary Fig. 28a-c**). Across these GWAS resources, cortical regions showed higher MDD TRSs than subcortical and cerebellar regions, with M1C showing a reproducible motor-cortical signal. This M1C-prioritized pattern was further corroborated in an independent mouse brain atlas across selected depression-relevant regions (**Supplementary Fig. 29**). Collectively, these analyses nominate M1C as a data-driven anatomical context for MDD-relevant neuronal risk and motivate further dissection of the underlying neuronal subpopulations.

### Subtype-level analyses implicate PVALB⁺ inhibitory and Ex-L2/4 excitatory neurons

Using the neuronal subtype annotation established above (**Fig. 2c,d**), we next asked which excitatory and inhibitory subpopulations contributed to the broad neuronal enrichment of MDD genetic risk. To assess subtype-level associations, we integrated four complementary enrichment frameworks—scPagwas, MAGMA-CellTyping, LDSC-SEG and scDRS—across the three MDD GWAS resources described above. Across these analyses, PVALB⁺ inhibitory neurons emerged as the most consistent inhibitory-neuron subtype, showing the strongest convergence in the primary European-ancestry discovery GWAS and remaining the top-ranked inhibitory subtype in both updated PGC GWAS datasets (**Fig. 4a**; **Supplementary Table 18**). Among excitatory subtypes, Ex-L2/4 neurons also showed recurrent enrichment across all three GWAS resources (**Fig. 4b**; **Supplementary Table 18**).

**Figure 4.**
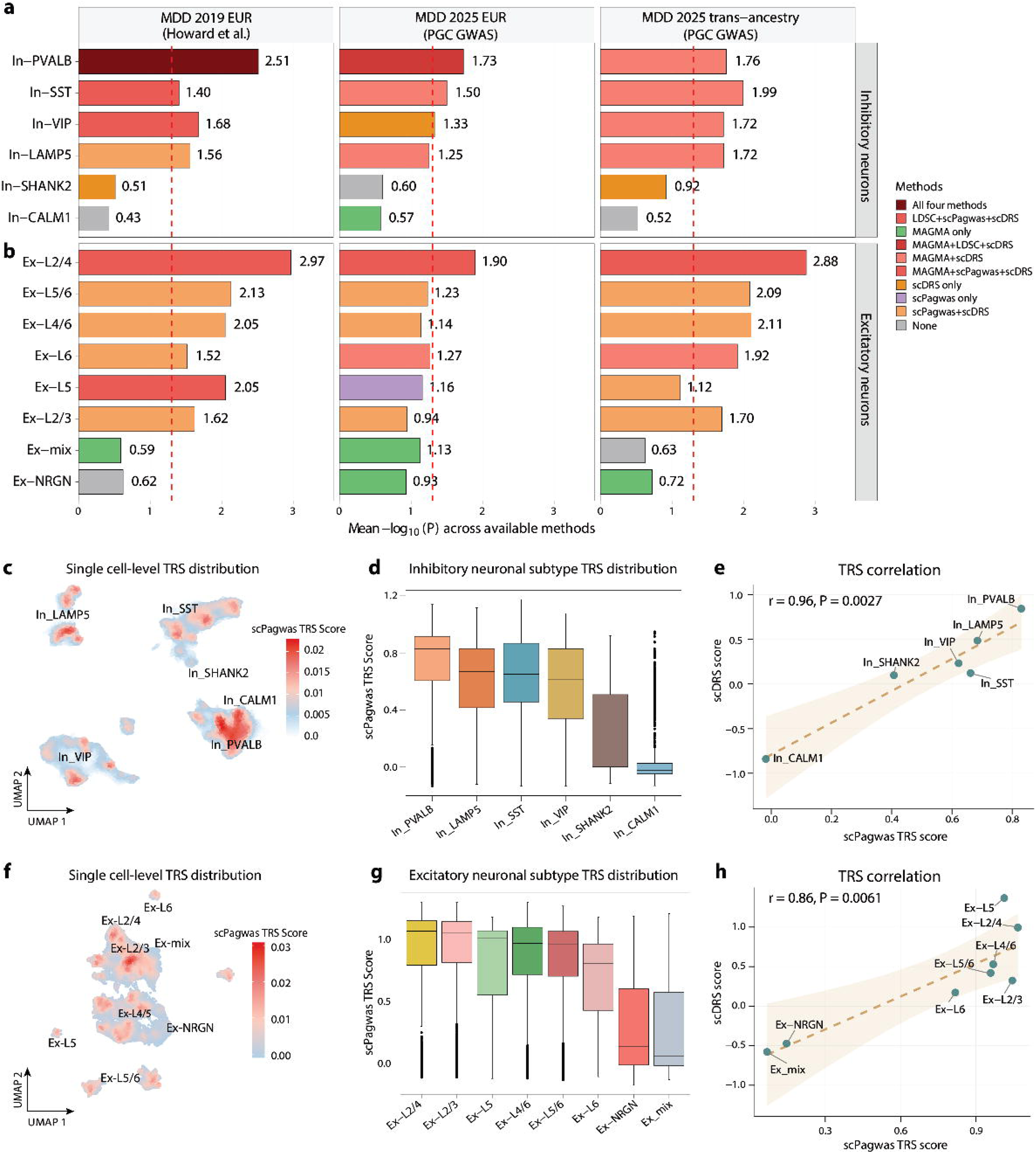
Subtype-level analyses nominate PVALB⁺ inhibitory neurons and L2/4 excitatory neurons as recurrent MDD-associated neuronal populations. **a,b,** Integrated subtype-level enrichment of MDD genetic risk across a, inhibitory neuronal subtypes and b, cortical excitatory neuronal subtypes in the adult human brain discovery atlas. Enrichment was assessed using four complementary approaches—scPagwas, MAGMA-CellTyping, LDSC-SEG and scDRS—across three MDD GWAS resources: Howard et al. 2019 European-ancestry GWAS, PGC MDD 2025 European-ancestry GWAS and PGC MDD 2025 trans-ancestry GWAS. LDSC-SEG was not applied to the trans-ancestry GWAS because of ancestry–LD-reference mismatch. For each neuronal subtype, horizontal bars show the mean −log₁₀(P) across available methods, with values indicated at the bar ends. Bar colours denote the combination of methods reaching nominal significance (P < 0.05; colour key). The dashed red vertical line indicates P = 0.05. **c.** UMAP visualization of inhibitory neuron subtypes, with cells coloured by single-cell scPagwas trait-relevance score (TRS) using the Howard et al. 2019 GWAS. Red indicates higher TRS and light blue indicates lower TRS. **d.** Distribution of single-cell scPagwas TRSs across inhibitory neuron subtypes. Box plots show the median, interquartile range and 1.5 × interquartile range; individual outlier cells are shown as points. PVALB⁺ inhibitory neurons showed the highest TRS distribution among inhibitory subtypes. **e.** Correlation between median subtype-level scPagwas TRS and scDRS score across inhibitory neuron subtypes. Each point represents one subtype; the dashed line indicates the linear regression fit and shading indicates the 95% confidence interval. scPagwas and scDRS showed concordant inhibitory subtype rankings. **f.** UMAP visualization of excitatory neuron subtypes, with cells coloured by single-cell scPagwas MDD TRS as in **c**. **g.** Distribution of single-cell scPagwas TRSs across excitatory neuron subtypes. Ex-L2/4 and Ex-L2/3 neurons showed the highest TRS distributions among excitatory subtypes. **h.** Correlation between median subtype-level scPagwas TRS and scDRS score across excitatory neuron subtypes. scPagwas and scDRS showed concordant excitatory subtype rankings.

We further refined these subtype-level associations at single-cell resolution. Single-cell-resolved scPagwas analyses prioritized PVALB⁺ inhibitory neurons as the highest-scoring inhibitory population, followed by LAMP5⁺ and SST⁺ inhibitory neurons (**Fig. 4c,d**). These findings were further supported by scDRS analyses across the three GWAS datasets (**Fig. 4e**; **Supplementary Fig. 30a-d**) and are consistent with prior evidence linking cortical PVALB⁺/SST⁺ inhibitory neurons and GABAergic dysfunction to MDD pathophysiology^70^. In parallel, Ex-L2/4 neurons emerged as the top excitatory-neuron subpopulation associated with MDD risk (**Fig. 4f-h**), a pattern that was consistently supported across all three GWAS datasets (**Supplementary Fig. 31a-d**).

We next asked whether the subtype-level neuron signals were reflected in the 308 high-confidence gene associations prioritized by the latest PGC depression GWAS^12^. Per-cell module scoring showed that this PGC-derived gene set had elevated activity in PVALB⁺ inhibitory neurons, which displayed the highest module activity among inhibitory subtypes (**Supplementary Fig. 32a**). An independent scDRS analysis using the same 308-gene set further supported PVALB⁺ inhibitory-neuron associations across the three MDD GWASs (**Supplementary Fig. 32b**). Ex-L2/4 excitatory neurons also showed elevated module activity and recurrent scDRS associations across GWAS resources (**Supplementary Fig. 32a-b**). These results show that MDD genetic risk is concentrated in specific neuronal subtypes rather than uniformly distributed across broad neuronal classes.

Consistent with the brain-wide regional scPagwas analysis, M1C showed elevated TRS distributions within inhibitory neurons, excitatory neurons and the prioritized PVALB⁺ and Ex-L2/4 subtypes (**Supplementary Fig. 33a-d**), indicating that the M1C signal persisted within the neuronal populations prioritized by subtype-level analyses. This regional pattern was independently supported by per-cell scDRS scoring using the Howard et al. GWAS, which showed higher disease-relevance scores in cortical regions, including M1C, across the same broad neuronal classes and prioritized subtypes (**Supplementary Fig. 34a**). Similar regional distributions were observed using the updated PGC European-ancestry and trans-ancestry GWAS datasets (**Supplementary Fig. 34b, c**). Taken together, these analyses identify PVALB⁺ inhibitory neurons as a reproducible context of MDD genetic risk, with Ex-L2/4 neurons representing a parallel excitatory signal.

### Integrative gene prioritization identifies a CNNM2-linked PVALB⁺ inhibitory-neuron state

To nominate candidate MDD risk genes acting within PVALB⁺ inhibitory neurons, we integrated MAGMA gene-level association^64^, FUSION transcriptome-wide association (TWAS) ^65^, and single-cell expression evidence. MAGMA identified 907 MDD-associated genes and FUSION identified 218 transcriptome-wide associated genes, with 21 genes shared between the two analyses (**Fig. 5a**; **Supplementary Tables 19–21**). Evaluation of these risk genes across three PVALB⁺-focused expression criteria—single-cell TRS association, enrichment relative to major brain cell classes and specificity relative to other inhibitory-neuron subtypes—identified *CNNM2* as the top-ranked PVALB⁺-linked candidate (**Fig. 5a**; **Methods**). *CNNM2* encodes a divalent cation transporter involved in neuronal function and magnesium homeostasis, has been linked to neuropsychiatric phenotypes including MDD^71–75^, and belongs to the 308 high-confidence MDD risk genes prioritized by the latest PGC study^12^. Multiple depression GWASs have also identified genome-wide significant associations at the *CNNM2* locus^4,12,76,77^.

**Figure 5.**
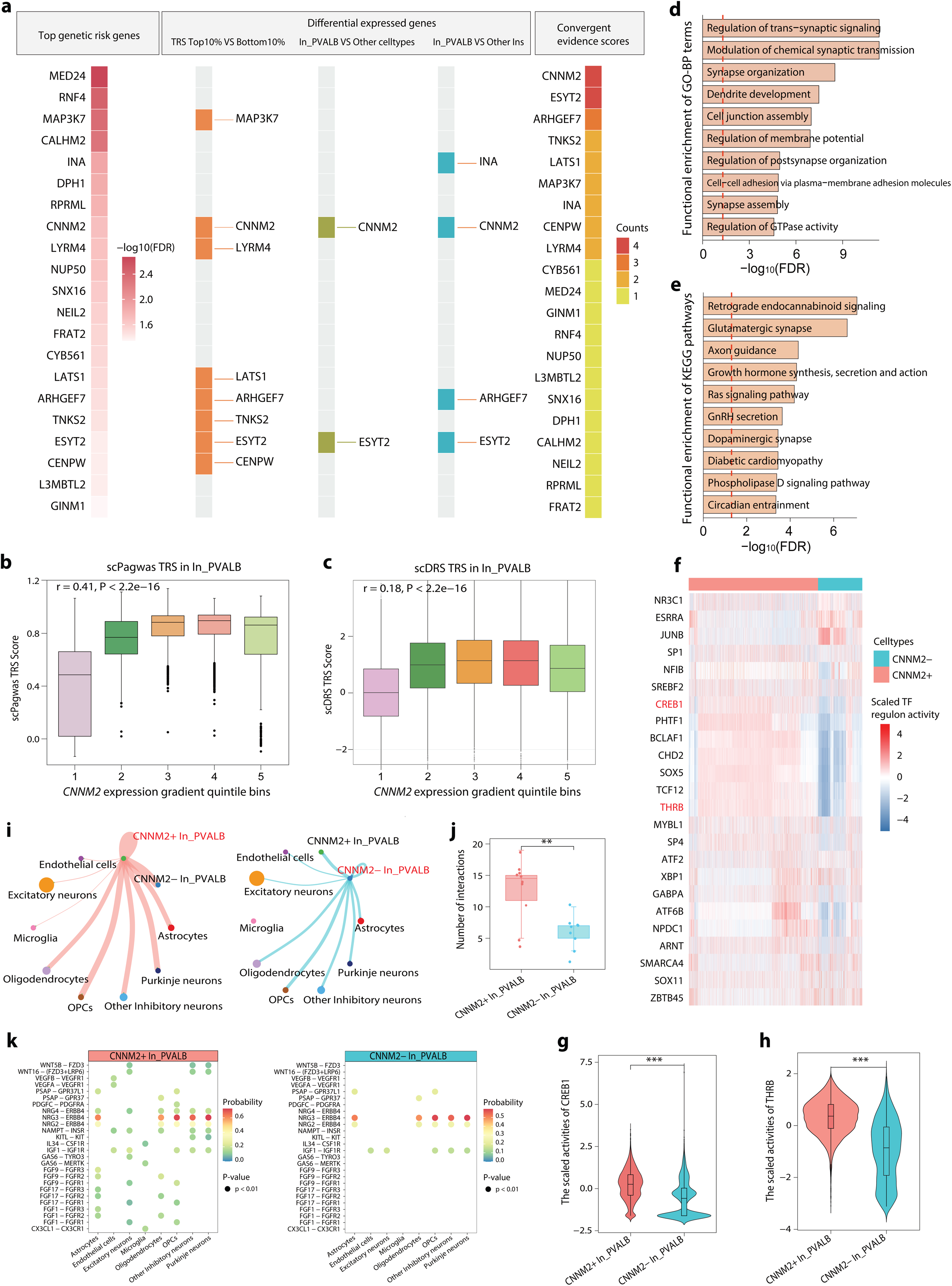
Prioritizing a *CNNM2*-centered PVALB⁺ inhibitory-neuron state associated with MDD risk. **a.** Integrative prioritization of candidate MDD risk genes in PVALB⁺ inhibitory neurons. The first evidence layer shows top genetic risk genes defined by MAGMA gene-wise association and overlap with FUSION TWAS-prioritized genes, with FDR values derived from MAGMA. Additional evidence layers include differential expression between cells with top and bottom 10% TRSs, differential expression between PVALB⁺ inhibitory neurons and other major brain cell classes, and differential expression between PVALB⁺ inhibitory neurons and other inhibitory-neuron subtypes. The right panel summarizes convergent evidence scores across evidence layers. **b.** Distribution of scPagwas TRSs across *CNNM2* expression-gradient quintile bins in PVALB⁺ inhibitory neurons. **c.** Distribution of scDRS TRSs across *CNNM2* expression-gradient quintile bins in PVALB⁺ inhibitory neurons. **d,e.** Functional enrichment analysis comparing *CNNM2*⁺ and *CNNM2*⁻ PVALB⁺ inhibitory neurons, showing enriched Gene Ontology biological process terms (d) and KEGG pathways (e). Bar length indicates −log₁₀(FDR). **f.** Heatmap of transcription factor (TF) regulon activities inferred by pySCENIC in *CNNM2*⁺ and *CNNM2*⁻ PVALB⁺ inhibitory neurons. Rows indicate TF-regulons and columns indicate single cells grouped by *CNNM2* expression status. Colour indicates scaled TF-regulon activity. **g,h.** Violin plots showing regulon activity scores for representative transcription factors CREB1 **(h)** and THRB **(i)** in *CNNM2*⁺ versus *CNNM2*⁻ PVALB⁺ inhibitory neurons. **i.** CellChat-inferred intercellular communication networks involving *CNNM2*⁺ and *CNNM2*⁻ PVALB⁺ inhibitory neurons and other annotated brain cell populations. Edge width indicates the number or strength of predicted interactions. **j.** Quantification of the number of predicted ligand–receptor interactions involving *CNNM2*⁺ and *CNNM2*⁻ PVALB⁺ inhibitory neurons. **k.** Dot plots showing representative CellChat-inferred ligand–receptor interactions between *CNNM2*⁺ or *CNNM2*⁻ PVALB⁺ inhibitory neurons and other annotated brain cell populations. Dot colour denotes inferred communication probability, and dot size indicates statistical significance; only significant interactions are shown. Box plots show median and interquartile range; whiskers indicate 1.5× interquartile range. Statistical significance was assessed using two-sided Wilcoxon rank-sum tests unless otherwise indicated. P < 0.01, *P < 0.001.

To further define the molecular state of *CNNM2*⁺ PVALB⁺ inhibitory neurons, we examined the relationship between *CNNM2* expression and single-cell genetic risk scores. In analyses based on the primary Howard et al. GWAS, *CNNM2* expression was positively associated with scPagwas TRS, with the strongest associations observed within inhibitory neurons and PVALB⁺ inhibitory neurons (**Supplementary Fig. 35a–c**). Higher *CNNM2* expression quintiles consistently showed progressively higher TRS values (**Fig. 5b**; **Supplementary Fig. 35d, e**). This relationship was further supported by scDRS scores derived from the same GWAS (**Fig. 5c**; **Supplementary Fig. 35f-j**) and was recapitulated using the updated PGC European-ancestry and trans-ancestry MDD GWAS datasets (**Supplementary Fig. 36a-l**), supporting *CNNM2* as a marker of a reproducible high-risk PVALB⁺ inhibitory-neuron state. Given the regional prioritization of M1C in the preceding analyses, we next examined whether *CNNM2* expression showed a corresponding regional pattern. *CNNM2* expression was elevated in M1C across all cells, inhibitory neurons and PVALB⁺ inhibitory neurons (**Supplementary Fig. 37a-c**). The preservation of this M1C-enriched pattern within PVALB⁺ inhibitory neurons link the *CNNM2*⁺ PVALB⁺ state to the motor-cortical signal identified above.

Functional enrichment analysis comparing *CNNM2*⁺ and *CNNM2*⁻ PVALB⁺ neurons showed that *CNNM2*⁺ cells were enriched for synaptic organization, trans-synaptic signalling, dendrite development and membrane-potential regulation, with KEGG pathway enrichment for retrograde endocannabinoid signalling, glutamatergic synapse, axon guidance, Ras signalling and dopaminergic synapse (**Fig. 5d, e**). We next tested whether the *CNNM2*⁺ PVALB⁺ state was detectable in an independent human MDD case–control dataset^31^. After re-annotation of inhibitory subtypes, *CNNM2* was preferentially expressed in PVALB⁺ inhibitory neurons, in line with the discovery atlas (**Supplementary Fig. 38a–d**). *CNNM2*⁺ inhibitory neurons showed higher synaptic and nervous-system process signature scores than *CNNM2*⁻ inhibitory neurons in both cases and controls (**Supplementary Fig. 38e, f**), supporting a reproducible *CNNM2*⁺ PVALB⁺ program in human MDD case-control brain tissue.

To identify transcriptional regulatory programs associated with the *CNNM2*⁺ PVALB⁺ inhibitory-neuron state, we inferred transcription factor (TF) regulon activity using pySCENIC^78^ in the discovery atlas (**Fig. 5f**). Among 24 active TF regulons in PVALB⁺ neurons, CREB1 and THRB showed significantly higher regulon activity in *CNNM2*⁺ than *CNNM2*⁻ PVALB⁺ cells (**Fig. 5g–h**). CREB/CREB1 signalling has been linked to activity-dependent neuronal plasticity, depression-related neurobiology and antidepressant response^79^, whereas *THRB* encodes thyroid hormone receptor β, consistent with longstanding evidence connecting thyroid signalling and depressive symptoms^80^. In the independent replication dataset^31^, THRB regulon activity was also elevated in *CNNM2*⁺ PVALB⁺ neurons (**Supplementary Fig. 39a, b**).

We next asked whether the *CNNM2*⁺ PVALB⁺ state was associated with altered intercellular communication and co-expression network organization. Cell-to-cell communication analysis^81^ showed that *CNNM2*⁺ PVALB⁺ neurons had more predicted ligand–receptor interactions with other brain cell populations than *CNNM2*⁻ PVALB⁺ neurons (**Fig. 5i-j**), including increased WNT and FGF signalling pairs and enhanced *CX3CL1–CX3CR1* interactions with microglia (**Fig. 5k**). Similar increases in FGF and neuroimmune signalling were observed in the replication dataset (**Supplementary Fig. 40a-b**), paralleling altered trophic and neuroimmune communication programs in *CNNM2*⁺ PVALB⁺ neurons. Co-expression network analysis^82^ further placed *CNNM2* within a PVALB⁺ neuron module enriched for MAPK signalling and synaptic pathways (**Supplementary Fig. 40c**). Protein–protein interaction mapping^83^ of module hub genes linked *CNNM2* to MAPK-related network hubs, suggesting that the *CNNM2*⁺ PVALB⁺ state is embedded within a broader synaptic and kinase-signalling program (**Supplementary Fig. 40d**).

As a parallel excitatory-neuron analysis, the same prioritization framework nominated *NEGR1* as the top-ranked Ex-L2/4-linked candidate gene in both the discovery atlas and an independent replication dataset (**Supplementary Figs. 41–43**), consistent with prior functional and genetic evidence implicating *NEGR1* in depression-related biology^4,5,7,10,84,85^. In summary, these analyses identify a *CNNM2*-centered PVALB⁺ inhibitory-neuron program as the principal molecular signal emerging from subtype-level MDD genetic prioritization.

### Chronic stress attenuates Cnnm2 protein levels in motor-cortical PV^+^ neurons

To evaluate the *in vivo* relevance of the human genetically prioritized *CNNM2*-linked PVALB⁺ (PV⁺) inhibitory-neuron program, we exposed adult male C57BL/6J mice to a 21-day chronic unpredictable mild stress (CUMS) paradigm (**Fig. 6a, b**). CUMS mice exhibited increased immobility in both tail suspension and forced swim tests, together with reduced time in the centre of the open field, whereas total distance travelled, and movement speed were unchanged, supporting a depression-like rather than locomotor phenotype (**Fig. 6c-e**). Subsequently, we profiled Cnnm2 protein expression across coronal sections of the mouse brain. Cnnm2 showed marked regional specificity, with strong signal in the primary and secondary motor cortex (M1/M2) and sensory cortical regions, but minimal signal in the piriform cortex and caudate putamen (**Supplementary Fig. 44a**). This motor-cortical enrichment was consistent with the M1-prioritized anatomical context identified by the human single-cell genetic analyses.

**Figure 6.**
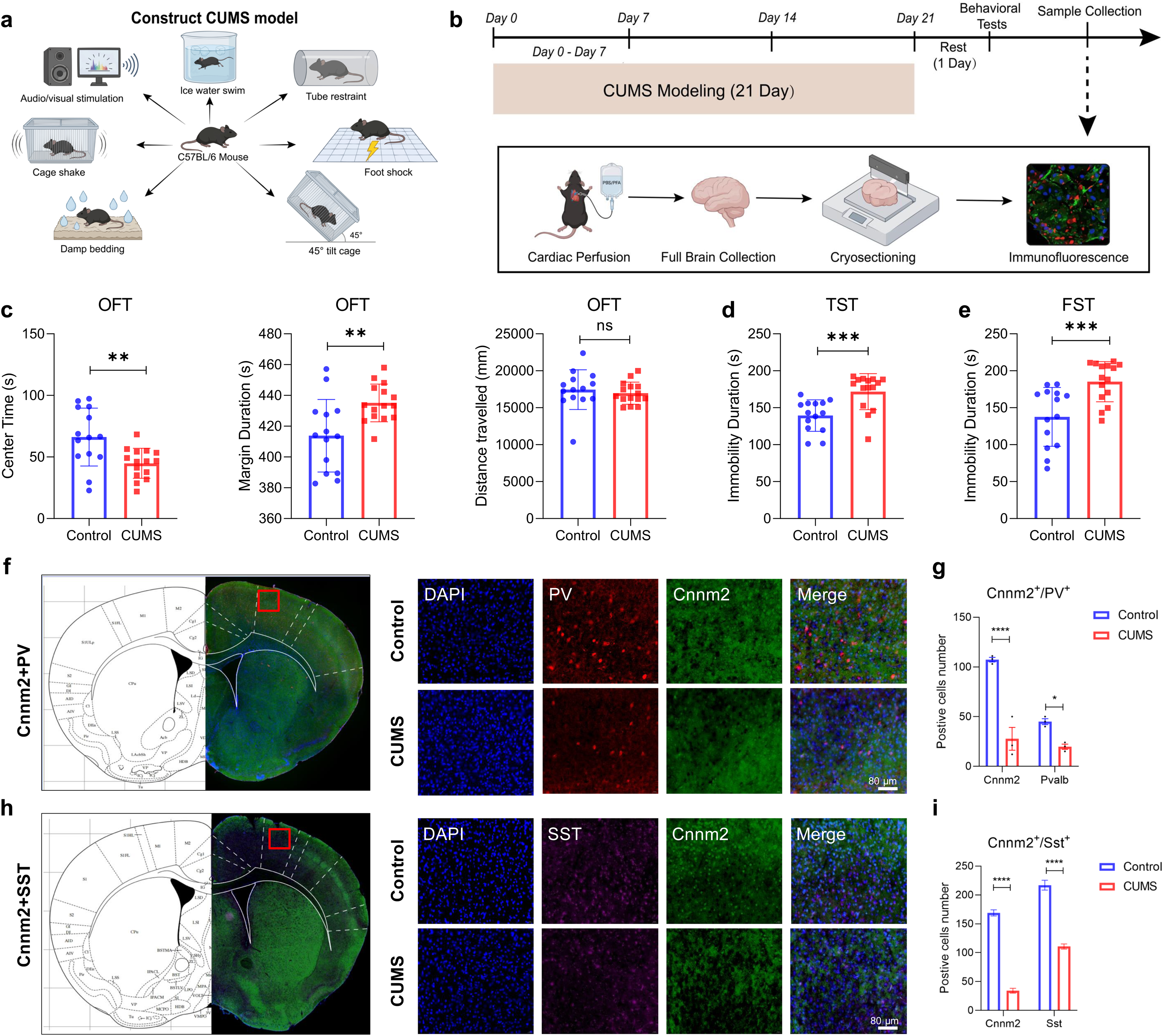
Verification of Cnnm2 expression in motor-cortical inhibitory neurons under chronic stress. **a.** Schematic illustration of the chronic unpredictable mild stress (CUMS) procedure in C57BL/6 mice. **b.** Schematic diagram of the experimental process. The timeline illustrates the 21-day CUMS modeling period, behavioral testing, and subsequent sample collection procedures, including cardiac perfusion, whole-brain collection, cryosectioning, and immunofluorescence. **c.** Anxiety-like behavioral tests following 21-day CUMS by open field test (OFT). **d,e.** Depressive-like behavioral tests following 21-day CUMS by forced swimming test (FST) and tail suspension test (TST). **f,g.** Expression of Cnnm2 in (**f**) PV^+^ and (**g**) SST^+^ inhibitory neurons. Left: Representative brain section indicating the immunofluorescent staining of Cnnm2 and PV^+^/SST^+^ neurons. Right: Summary data of PV^+^ neurons, Cnnm2/SST^+^ neurons, and their colocalization in M1 region in both Control and CUMS groups. Scale bar, 80 μm. *p < 0.05, **p < 0.01, ***p < 0.001, ****p < 0.0001, ns: not significant.

We next examined whether chronic stress altered Cnnm2 protein levels in motor-cortical inhibitory neurons. In M1, CUMS mice showed reduced Cnnm2 and PV fluorescence signals relative to controls (**Fig. 6f**). Quantitative analysis further confirmed a significant reduction in the number of *Cnnm2*⁺ and PV⁺ neurons in CUMS-treated mice (P < 0.05; **Fig. 6g**). Co-immunostaining of *Cnnm2* with somatostatin (SST) revealed a similar pattern, with weakened Cnnm2/SST co-staining intensity in M1 after CUMS (**Fig. 6h, i**). These results indicate that chronic stress attenuates Cnnm2 protein levels across prioritized inhibitory-neuron populations in the motor cortex. Parallel staining of *Arhgef7*, another PV⁺-linked MDD risk gene prioritized by our integrative analysis, with PV or SST showed comparable reductions in stressed mice (**Supplementary Fig. 44b**). Together, these results provide *in vivo* support for a motor-cortical *Cnnm2*⁺ inhibitory-neuron state that is sensitive to chronic stress, linking human single-cell genetic prioritization to stress-associated molecular changes in PV⁺/SST⁺ inhibitory neurons.

### Optogenetic activation of M1 PV^+^ neurons reverse stress-induced depression-like behaviours

Motivated by the motor-cortical enrichment of *CNNM2* and the prioritization of M1C in the human single-cell genetic analyses, we next asked whether M1 projections engage mood-regulatory brain regions and contribute to depression-like behaviour. Anterograde tracing with AAV2/9-hSyn-mCherry injected into M1 labelled M1-derived axonal fibres across multiple downstream targets, including M2, contralateral M1, caudate–putamen, thalamic nuclei, zona incerta, substantia nigra reticulata, superior colliculus, red nucleus and the lateral/ventrolateral periaqueductal gray (l/vlPAG) (**Fig. 7a, b**; **Supplementary Fig. 45a-l**). Among these targets, l/vlPAG was of particular interest because PAG subregions have established roles in anxiety, depression, defensive behaviour and pain modulation^86–88^. Retrograde tracing with AAV2-Retro-eGFP injected into l/vlPAG labelled neuronal somata in layer 5 of M1, confirming a direct layer-specific M1→l/vlPAG projection (**Fig. 7c, d**).

**Figure 7.**
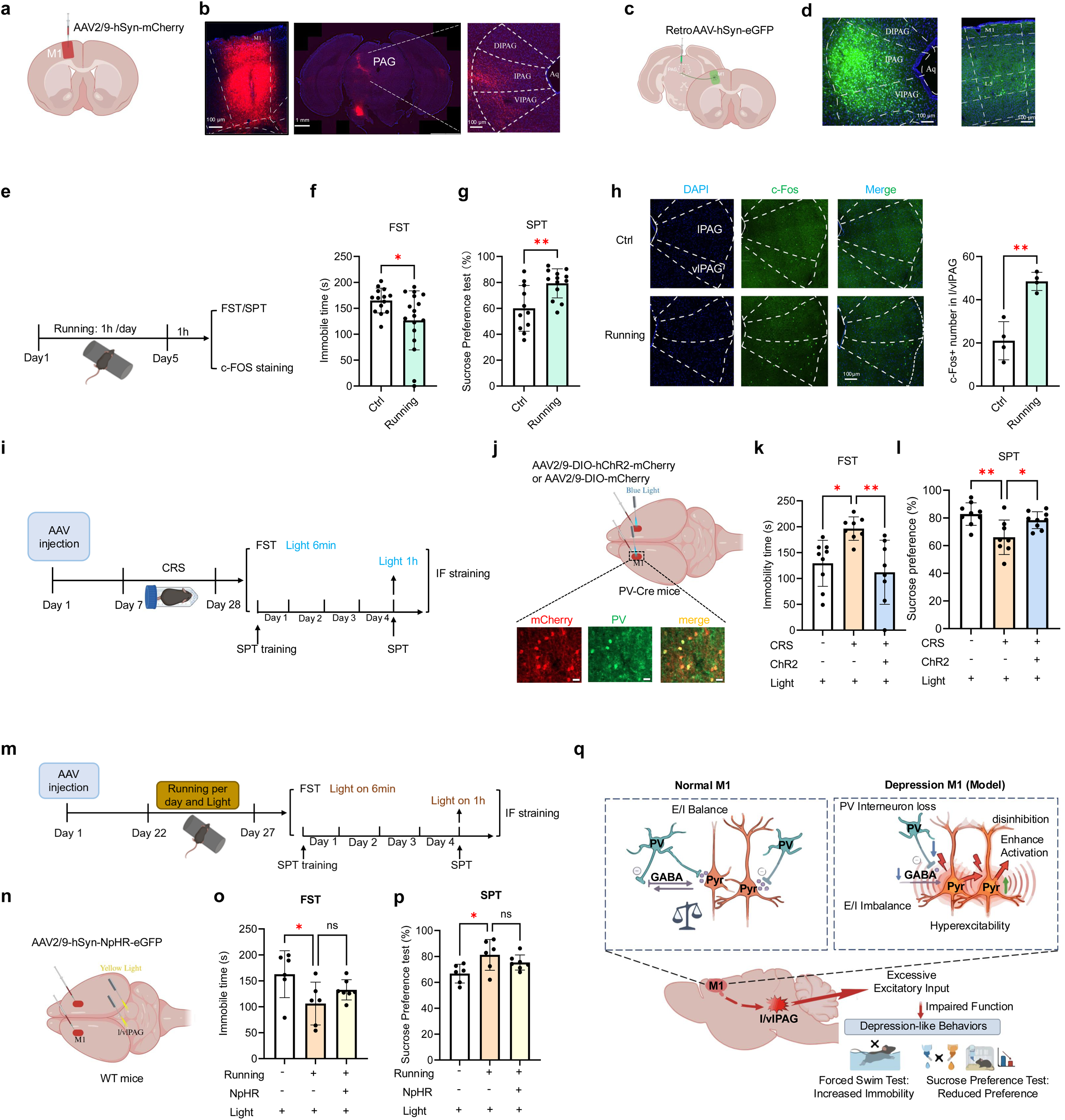
Exercise alleviates depressive-like behaviors caused by dysfunction of M1 PV^+^ neurons. **a,b**. Schematic showing the injection site of AAV2/9-hSyn-mCherry in the primary motor cortex (M1) (a). Three weeks after viral delivery, abundant mCherry-labeled fibers were detected in the cerebral peduncle (CP) and periaqueductal gray (PAG) (middle panels). Magnified views (right) show labeled fibers in the lateral (lPAG) and ventrolateral (vlPAG) subregions, but not the dorsolateral PAG (dlPAG). **c,d**. Schematic illustrating RetroAAV-hSyn-eGFP injection into the dlPAG. Three weeks post injection, eGFP-positive neurons were found in layer V of M1. **e.** Experimental timeline for rotarod training, subsequent behavioral assessments and c-Fos immunostaining. **f–h**. Rotarod training decreased immobility duration in the forced swim test (FST) (f), increased sucrose preference in the sucrose preference test (SPT) (g), and upregulated c-Fos expression in the lPAG and vlPAG (h). **i.** Schematic overview of the experimental design. **j.** Upper: Schematic of viral injection and optogenetic activation of PV^+^ neurons in M1. Lower: Representative immunofluorescence images showing mCherry, PV signals and their colocalization in M1. Scale bars: 20 μm. **k,l**. Optogenetic activation of M1 PV^+^ neurons rescued chronic restraint stress (CRS)-induced depressive-like behaviors in the FST (k) and SPT (l). **m.** Experimental design for experiments shown in (n) and (o). **n.** Schematic of viral injection and optogenetic inhibition targeting the M1–PAG pathway. **o,p**. Rotarod exercise exerted robust antidepressant-like effects in the FST (o) and SPT (p). Optogenetic suppression of the M1–PAG pathway produced no additional antidepressant benefits on top of running. **r.** Working model: Dysfunction of M1 PV^+^ neurons contribute to the development of depression-like behaviors. Rotarod training reverses these behavioral deficits by attenuating excitatory inputs from M1 to the PAG. Data are presented as mean ± SEM. *P < 0.05, **P < 0.01; ns, not significant (statistical analyses and n values).

We next examined whether l/vlPAG was recruited during an antidepressant behavioural intervention. Five consecutive days of rotarod running reduced forced-swim immobility and increased sucrose preference relative to non-running controls, consistent with an antidepressant-like effect of exercise (**Fig. 7e-g**). Rotarod running also increased c-Fos expression in both lPAG and vlPAG (**Fig. 7h**), suggesting that l/vlPAG is engaged during exercise-associated behavioural improvement.

To test the functional relevance of M1 PV+ neurons and the M1–l/vlPAG pathway, we selectively activated M1 PV+ neurons optogenetically in mice exposed to chronic restraint stress (CRS). This manipulation rescued CRS-induced depressive-like behaviors in the forced-swim and sucrose-preference tests (**Fig. 7i–l**). In parallel, rotarod exercise fully reversed CRS-associated depression-like phenotypes, whereas optogenetic suppression of the M1–l/vlPAG pathway produced no additional behavioural improvement beyond exercise alone (**Fig. 7m–p**). This occlusion effect suggests that exercise-associated antidepressant-like effects may converge on the M1–l/vlPAG pathway. Together, these anatomical tracing, activity-mapping and optogenetic experiments identify M1 PV⁺ neurons and the M1→l/vlPAG projection as a motor-cortical circuit axis linking the cellular program prioritized by single-cell genetic analyses to depression-like behaviour and exercise-associated antidepressant effects (**Fig. 7q**).

### scDepBrain provides an interactive resource for MDD-linked brain cell types and states

To facilitate exploration and reuse of the integrated single-cell atlas and MDD genetic-prioritization results, we developed the scDepBrain, a web-accessible resource for querying MDD-associated cellular programs across brain contexts. scDepBrain organizes more than five million cells and nuclei from 34 published brain single-cell and single-nucleus studies into searchable dataset entries with metadata including species, brain region, developmental stage, disease status, data source, accession number, cell count and omics type (**Supplementary Fig. 46**; **Supplementary Tables 1–4**). The platform provides dataset-level visualization and analysis modules, including UMAP exploration, selectable gene-expression display, marker-gene inspection, cellular-composition comparison, differential-expression analysis and GO/KEGG pathway enrichment (**Fig. 1f**; **Supplementary Fig. 47a,b**). scDepBrain also enables interactive exploration of genetic-prioritization results, including single-cell TRSs and cell-type-level MDD association results (**Supplementary Fig. 47c**). Collectively, scDepBrain provides a public platform for cross-dataset comparison, hypothesis generation and future investigation of the cellular and molecular architecture of MDD.

## DISCUSSION

In this study, we integrated human brain single-cell atlases, large-scale MDD GWAS resources and experimental circuit analyses to map MDD genetic liability to cellular populations, candidate genes and *in vivo* circuitry. Across complementary polygenic-enrichment frameworks^39–41,44^, MDD common-variant risk was preferentially enriched in neuronal populations of the adult brain, particularly excitatory and inhibitory neurons. This result is consistent with previous genetic and transcriptomic studies implicating neuronal rather than broadly glial programs in depression-related phenotypes^12,18,45,89^, and extends them by resolving trait-relevant cellular and anatomical contexts at single-cell resolution. Our analyses further nominated M1C as a motor-cortical context for MDD risk and identified PVALB⁺ inhibitory neurons and layer-specific excitatory neurons as major neuronal substrates. Thus, these findings move beyond cell-type nomination by linking MDD polygenic risk to regional, subtype-specific and molecularly defined neuronal states.

A central finding is the prioritization of a *CNNM2*-centered PVALB⁺ inhibitory-neuron state. *CNNM2* emerged as the top PVALB⁺-linked candidate through integration of gene-level association, transcriptome-wide association, and cell-state-specific expression evidence. Its expression was coupled to disease-relevance scores across different cellular contexts, and *CNNM2*⁺ PVALB⁺ neurons were enriched for synaptic organization, trans-synaptic signalling, dendrite development, membrane-potential regulation, and activity-related pathways. This aligns with broader evidence that cortical GABAergic dysfunction, including PVALB⁺ and SST⁺ interneuron abnormalities, contributes to stress and depression biology^70^. The identification of CREB1 and THRB regulon activity, MAPK-linked co-expression modules and altered trophic/neuroimmune communication suggests that the *CNNM2*⁺ PVALB⁺ state reflects a coordinated molecular program rather than a single-gene signal. Prior proteome-wide and genetic studies^71–75^ have implicated *CNNM2* or *CNNM2*-related biology in depression and neuropsychiatric traits, providing external support for its prioritization here. In parallel, the prioritization of *NEGR1* in Ex-L2/4 neurons suggests that MDD genetic risk also converges on layer-specific excitatory-neuron states, complementing the *CNNM2*-centered inhibitory-neuron program.

The prioritization of M1C is unexpected considering the historical emphasis on prefrontal, limbic and reward-related regions in depression. However, motor cortical regions are increasingly recognized as participating in sensorimotor integration, action selection, motivational control and psychomotor features of depression^52,55,57–59^. Our data place this biology in a genetic and cellular framework: M1C showed higher single-cell TRS values, *CNNM2* expression was elevated in M1C within PVALB⁺ inhibitory neurons, Cnnm2 protein was enriched in mouse M1/M2, and chronic stress attenuated Cnnm2 expression in motor-cortical PVALB⁺/SST⁺ inhibitory neurons. Rather than replacing prefrontal or limbic models of MDD, these results expand them by identifying the motor cortex as an underexplored anatomical context in which inherited risk may converge on inhibitory-neuron dysfunction.

The circuit experiments provide functional support for this model by linking the M1 PV⁺ inhibitory-neuron signal to PAG-associated behavioural regulation. Anatomical tracing identified a direct M1 projection to l/vlPAG, a midbrain region involved in defensive behaviour, pain modulation, anxiety and depression-like states^61–63,86^. Rotarod exercise recruited l/vlPAG activity and produced antidepressant-like behavioural effects, in line with clinical and preclinical evidence that exercise can reduce depressive symptoms^60^. Cell-type-specific optogenetic activation of M1 PV⁺ neurons alleviated stress-induced depression-like behaviours, whereas the interaction between exercise and M1–l/vlPAG suppression suggests that exercise-associated behavioural improvement may involve overlapping motor-cortical–PAG circuitry. Although these experiments do not establish *CNNM2* as a causal mediator of circuit function, they provide an experimentally tractable link between the human single-cell genetic signal and motor-cortical inhibitory control of depression-like behaviour.

More broadly, our study illustrates how single-cell genetic mapping can bridge human polygenic association signals with experimentally tractable cellular and circuit hypotheses. By combining an adult human brain discovery atlas with replication atlases spanning adult human brain, developing human brain, mouse brain, cerebral organoids and human MDD case–control brain tissue, we assessed regional specificity, developmental timing, cross-species relevance, and disease-state expression of MDD-associated cellular programs. The developing human brain showed relatively stronger glial-lineage signals than adult brain datasets, suggesting that MDD genetic liability may intersect with distinct cellular programs across developmental stages. This pattern is consistent with epidemiological evidence that MDD and other mental disorders frequently emerge during adolescence and early adulthood^67,68^, as well as transcriptomic studies showing extensive developmental remodeling of human brain cell states and gene-expression programs^66,90–92^. Thus, adult neuronal vulnerability and developmental maturation programs may represent complementary dimensions of MDD biology.

To facilitate reuse and community exploration, we developed scDepBrain, a web-accessible interactive resource that organizes the integrated atlas and genetic-prioritization results into searchable, visualization and downloadable modules. scDepBrain enables users to query MDD-associated cell types, genes, pathways and trait-relevance patterns across discovery and replication datasets and will be updated as additional brain single-cell, genetic and disease datasets become available. This resource provides a practical platform for hypothesis generation, cross-dataset comparison, and future experimental follow-up.

Several limitations should be considered. First, although the atlas integrates many public datasets, differences in sequencing platform, brain-region sampling, donor composition and annotation depth may influence cell-type representation and regional comparisons. Second, our genetic analyses rely primarily on common-variant GWAS summary statistics and do not capture rare variants, structural variants, or environmental exposures. Statistical associations between MDD liability and individual cells or cell types should also not be interpreted as direct evidence of causality, as they may reflect causal cellular contexts, correlated transcriptional states or downstream consequences of genetically influenced programs. Nevertheless, convergence across complementary polygenic frameworks supports the biological relevance of the prioritized cells. Third, our primary SNP-to-gene assignment used a proximal window, following previous studies^23,39^; future work incorporating fine-mapping, chromatin accessibility, enhancer–gene links, chromatin conformation and cell-type-specific eQTLs may improve variant-to-gene resolution^17–19^. Fourth, the mouse experiments support motor-cortical inhibitory circuitry but do not establish that *CNNM2* causally mediates behavioural phenotypes; cell-type-specific *CNNM2* perturbation will be needed to test this mechanism directly. Finally, because exercise engages multiple peripherals, cortical, subcortical and neuromodulatory pathways, the M1→l/vlPAG projection is likely one component of a broader circuit network.

### Conclusion

In summary, our study maps MDD polygenic risk from human single-cell brain atlases to neuronal populations, candidate genes, and experimentally tractable circuits. By converging on a *CNNM2*-centered PVALB⁺ inhibitory-neuron state and an M1 → l/vlPAG circuit axis, our findings nominate motor-cortical inhibitory biology as an underexplored dimension of MDD risk and exercise-associated antidepressant-like effects. This work provides a framework for translating human genetic associations into cell-state- and circuit-level hypotheses for neuropsychiatric diseases, supported by scDepBrain as a public resource for future discovery.

## METHODS

### Ethical statement

This study complied with all relevant ethical regulations and the principles of the Declaration of Helsinki. All human genomic analyses were performed exclusively using publicly available single-cell and single-nucleus brain datasets and GWAS summary statistics. No new human participants were recruited, and no identifiable private information was accessed. Ethics approval and written informed consent for the human datasets were obtained by the original studies. All mouse experiments were approved by the Animal Care and Use Committee of Hangzhou City University and were conducted in accordance with institutional guidelines for animal care and use.

### Single-cell and single-nucleus brain datasets

We curated publicly available human and mouse brain single-cell and single-nucleus transcriptomic datasets from the Gene Expression Omnibus, Single Cell Portal, and other public repositories. Searches were performed using combinations of terms related to single-cell profiling, including “scRNA-seq”, “single-cell RNA-seq”, “single-cell transcriptome”, “snRNA-seq”, “single-nucleus RNA-seq” and “single-cell sequencing”, together with “brain”, “nervous system” and names of specific brain regions. Duplicate datasets, cell lines and non-brain samples were excluded after manual inspection.

For the discovery atlas, we retained adult human brain single-cell/nucleus datasets generated from non-disease samples and spanning 14 anatomical regions: anterior cingulate cortex (ACC), caudate nucleus (CN), cortex (CTX), frontal cortex (FC), substantia nigra (SN), cerebellar cortex (CER), primary somatosensory cortex (S1C), primary motor cortex (M1C), dorsolateral prefrontal cortex (DFC), prefrontal cortex (PFC), middle temporal gyrus (MTG), primary auditory cortex (A1C), primary visual cortex (V1C), and lateral amygdala (LA). Samples were further assigned to postnatal age periods and anatomical regions according to the original metadata and published annotations. Independent replication atlases were assembled to evaluate cross-dataset robustness, developmental timing, cross-species relevance, and disease-state expression, including adult human brain, developing human brain, mouse brain, cerebral organoid and human MDD case–control brain datasets (Supplementary Fig. 1 and Supplementary Tables 1-4).

### Single-cell data preprocessing and integration

Single-cell and single-nucleus transcriptomic datasets were processed using Seurat v4.1.0^93^. For each dataset, cells were first filtered using dataset-specific quality-control metrics provided by the original studies. Empty droplets, likely doublets or multiplets and low-quality cells were removed before integration. To further exclude outlier cells, we applied median absolute deviation (MAD)-based filtering using the *isOutlier* function from the scater package^94^. Cell-cycle scores were calculated using Seurat’s CellCycleScoring function. Gene expression counts were normalized and variance-stabilized using Seurat’s *SCTransform* function. Briefly, raw counts for each gene were normalized by total cellular counts, scaled to 10,000 transcripts per cell and transformed using a regularized negative-binomial regression model to reduce technical variation across cells. Cell-cycle-related variation and other dataset-specific technical effects were regressed when applicable. Highly variable genes were identified for each dataset, and the top 3,000 highly variable genes were used for dimensionality reduction.

Principal component analysis (PCA) was performed on the scaled expression matrix, and the leading principal components (PCs) were used for integration and clustering. To minimize batch effects across studies, donors and technical platforms, we applied Harmony^95^ using Seurat PCA embeddings as input. The top 50 Harmony-corrected principal components were retained for downstream analyses. The integrated dataset was visualized in two dimensions using uniform manifold approximation and projection (UMAP) based on these 50 corrected PCs. For clustering, we constructed a shared-nearest-neighbour (SNN) graph using Seurat’s *FindNeighbors* function with the top 50 Harmony-corrected PCs. Graph-based clustering was then performed using Seurat’s modularity optimization algorithm with a resolution of 0.2. The cells in the integrated dataset were clustered into 29 clusters.

### Cell-type and neuronal-subtype annotation

Major cell classes were annotated using canonical marker genes together with published adult human brain single-cell atlases^96,97^. After initial clustering of the integrated discovery atlas, each cluster was assigned to a major lineage according to the enrichment of well-established marker genes. Clusters without clear lineage-specific marker expression, or clusters showing mixed expression of markers from multiple major lineages, were annotated as ambiguous and excluded from downstream cell-type-level genetic analyses. This strategy assigned cells to eight major brain cell classes: excitatory neurons, inhibitory neurons, Purkinje neurons, astrocytes, oligodendrocytes, OPCs, microglia and endothelial cells.

To resolve neuronal heterogeneity, excitatory and inhibitory neurons were extracted separately and subjected to a second round of clustering using the same normalization, dimensionality-reduction and graph-based clustering framework described above. In the discovery atlas, excitatory neurons were resolved into layer-enriched subtypes, including Ex-L2/3, Ex-L2/4, Ex-L4/6, Ex-L5, Ex-L5/6, Ex-L6, Ex-NRGN and Ex-mix populations, whereas inhibitory neurons were resolved into six major interneuron subtypes, including PVALB⁺, SST⁺, VIP⁺, LAMP5⁺, SHANK2⁺ and CALM1⁺ populations. Subtype identities were assigned by combining canonical marker-gene expression, differentially expressed genes and published cortical neuron taxonomies^96,98^. Marker genes for each major cell type and neuronal subtype were identified using Seurat’s *FindAllMarkers* function with the Wilcoxon rank-sum test. Genes with Benjamini–Hochberg-adjusted P < 0.05 and at least 1.5-fold higher expression in one cluster relative to the average expression in the remaining clusters were considered marker genes.

### Cell-composition and stratified analyses

Cellular composition was quantified by calculating the proportion of each annotated cell type within each sample, anatomical region or age group. Similarity between brain regions was assessed using Pearson correlations of cell-type proportions, followed by hierarchical clustering. For age-stratified analyses, samples were grouped into postnatal developmental periods, including childhood, adolescence, young adulthood, middle adulthood and late adulthood, following published human brain developmental frameworks. For stratified genetic analyses, the discovery atlas was subset by sex, age period and brain region. Cell-type-level and single-cell-level prioritization analyses were then performed within each stratum to assess whether MDD genetic signals were sex-dependent, developmentally regulated or anatomically heterogeneous.

### GWAS summary statistics

We used the MDD GWAS summary statistics from Howard et al. as the primary genetic dataset^5^. This dataset was derived from UK Biobank and Psychiatric Genomics Consortium data, excluding 23andMe samples, and included 170,756 cases and 329,443 controls of European ancestry. Summary statistics were provided in HG19/GRCh37 coordinates and included SNP-level association statistics, effect and non-effect alleles, effect-size estimates and association P values.

To assess the robustness of MDD-associated cellular prioritization across updated genetic resources^12^, we further analyzed two PGC 2025 depression GWAS datasets excluding 23andMe samples: a European-ancestry GWAS comprising 412,305 cases and 1,588,397 controls, and a trans-ancestry GWAS comprising 537,363 cases and 2,061,567 controls. Both datasets were provided in GRCh37 coordinates and were used as complementary resources for subtype-level and regional validation analyses. As a negative-control trait to assess specificity, we also analyzed a European-ancestry standing-height GWAS comprising 461,950 individuals (**Supplementary Table 5**).

Before downstream analyses, we retained autosomal variants with minor allele frequency > 0.01 and excluded the extended major histocompatibility complex region (chr6: 25–35 Mb) because of its complex linkage disequilibrium (LD) structure^34,99–101^. Unless otherwise specified, LD was estimated using the 1000 Genomes Project Phase 3 European reference panel^102^ for analyses based on European-ancestry GWAS datasets. For trans-ancestry GWAS analyses, the same variant filtering and SNP-to-gene annotation framework was applied, and results were interpreted as complementary robustness analyses rather than ancestry-specific fine-mapping.

### Gene-wise association analysis

Gene-wise association statistics were computed using MAGMA v1.10^64^. SNPs were assigned to protein-coding genes if they mapped to the gene body or within 50 kb upstream or downstream of the gene boundary (--annotate window=50,50), using NCBI build 37 gene coordinates. Gene-based tests were performed using MAGMA’s default SNP-wise mean model, with the 1000 Genomes Project Phase 3 European reference panel used for LD estimation. For each GWAS, SNP-level P values and corresponding effective sample sizes were provided as input. Multiple testing across genes was controlled separately within each GWAS using the Benjamini-Hochberg false-discovery rate procedure, and genes with FDR ≤ 0.05 were considered significant.

The resulting MAGMA gene-level statistics were used for three downstream analyses. First, FDR-significant genes were used for risk-gene prioritization. Second, for scDRS (v1.0.3b)^44^ gene-set construction, the top 1,000 MAGMA-ranked genes per trait were converted from P values to z scores using an inverse-normal transformation and truncated to ±10. Third, for MAGMA-CellTyping^40^, MAGMA gene-level results were tested against cell-type expression-specificity scores as continuous gene-level covariates.

### Transcriptome-wide association analysis

Transcriptome-wide association analysis (TWAS) was performed using FUSION^65^ to identify genes whose genetically predicted expression was associated with MDD. FUSION integrates GWAS summary statistics with precomputed *cis*-expression prediction weights from reference transcriptomic panels and an LD reference panel to test associations between genetically regulated gene expression and trait risk. We applied FUSION to the processed MDD GWAS summary statistics using GTEx brain-tissue expression weight models. For each gene, FUSION generated a TWAS association statistic and corresponding P value. Genes with nominal TWAS P < 0.05 were retained as TWAS-supported candidates for downstream integration with MAGMA gene-wise association and cell-state-specific expression analyses. Ensembl gene identifiers from FUSION outputs were converted to gene symbols using clusterProfiler^103^, and duplicate or unmapped genes were removed before downstream analyses.

### Integration of MDD GWAS with single-cell transcriptomes using scPagwas

We used scPagwas v1.0.0^39^ to integrate processed MDD GWAS summary statistics with single-cell transcriptomic profiles and to prioritize MDD-relevant cells, cell types and genes. Analyses were performed independently for each of the three MDD GWAS resources: the primary Howard et al. European-ancestry GWAS^5^, the updated PGC European-ancestry GWAS and the updated PGC trans-ancestry GWAS^12^. The normalized single-cell gene-expression matrix was used as input, and KEGG pathways^104^ were used as the default pathway collection.

Briefly, scPagwas first projected pathway-specific expression programs into low-dimensional pathway activity scores using singular value decomposition. GWAS variants were mapped to genes using SNP-to-gene annotations described above, and pathway-level SNP annotations were generated according to pathway gene membership and genomic block annotation. For each pathway, scPagwas linked pathway activity patterns in single cells with GWAS-derived genetic signals while accounting for LD structure. This procedure generated genetically associated pathway activity scores for individual cells, which are aggregated across pathways to derive cell-level genetic relevance scores.

MDD-prioritized genes were identified by calculating the Pearson correlation between cell-level genetic relevance scores and gene expression across cells. Top positively correlated genes were then used to calculate trait-relevance scores (TRSs) for individual cells using matched background gene sets. Empirical cell-level significance was estimated by comparing observed TRSs with background-corrected random gene-set scores. To assess cell-type-level significance, cells were grouped according to their annotated cell populations, and corrected single-cell p values were combined within each cell type using the *Merge_celltype_p* function in scPagwas. Briefly, single-cell p values were converted to z scores, averaged within each cell type and transformed back to cell-type-level p values. Cell-type-level p values were then adjusted for multiple testing across annotated cell populations within each GWAS analysis. This framework enabled both cell-type-level prioritization and single-cell-level identification of MDD-relevant cellular states for downstream stratified, subtype-level and cross-GWAS validation analyses.

### Complementary cell-type and single-cell genetic prioritization

To assess the robustness of scPagwas-based cellular prioritization, we applied three complementary frameworks: scDRS^44^, partitioned LDSC-SEG^41^ and MAGMA-CellTyping^40^. Unless otherwise specified, analyses were performed using the MDD GWAS resources described above, with the updated PGC 2025 datasets used as cross-cohort and cross-ancestry robustness analyses. Multiple testing was controlled within each GWAS and analysis level using the Benjamini–Hochberg false-discovery rate procedure.

For single-cell-level validation, scDRS^44^ was used to compute per-cell disease-relevance scores. For each GWAS, MDD gene sets were constructed from MAGMA gene-wise association statistics by retaining the top 1,000 genes per trait, weighted by z scores converted from MAGMA P values and truncated to ±10. scDRS scored each cell for the weighted MDD gene set relative to 1,000 matched control gene sets with similar mean expression and expression variance, yielding a normalized disease-relevance score for each cell. Cell-type-level and subtype-level associations were assessed using the scDRS group-association framework, which aggregates per-cell scores within annotated populations and estimates Monte Carlo-based association P values. Concordance with scPagwas trait-relevance scores was evaluated at the single-cell and subtype levels using Pearson correlation.

For heritability-based validation, partitioned LDSC-SEG^41^ was used to test whether MDD SNP heritability was enriched in annotations linked to cell-type- or subtype-specific genes. Cell-type specificity was calculated using the Bryois specificity score, defined as the mean log-normalized expression of a gene in a given cell type divided by the summed mean expression of that gene across all cell types. For each annotated population, the top 10% specifically expressed genes were selected, and binary genomic annotations were generated for SNPs within 100 kb of these genes. LD scores were calculated for chromosomes 1–22 using the 1000 Genomes Project Phase 3 European reference panel^102^, HapMap3 SNPs and a 1-cM LD window. GWAS summary statistics were processed with munge_sumstats.py^105^ and restricted to HapMap3 SNPs. Stratified LD score regression was performed using the baseline-LD v2.2 model, an all-gene control annotation and regression weights excluding the MHC region. LDSC-SEG was applied to the European-ancestry GWAS datasets and was not applied to the trans-ancestry GWAS because of a mismatch between trans-ancestry LD structure and the European LD reference panel.

For gene-association-based validation, MAGMA-CellTyping^40^ was used to test whether cell-type expression specificity was associated with MDD gene-level association strength. SNP-level GWAS statistics were converted to gene-level association statistics using MAGMA v1.10^64^ with the gene-mapping strategy described above, including a ±50-kb gene window, exclusion of the extended MHC region, the 1000 Genomes Phase 3 European LD reference panel and MAGMA’s SNP-wise mean model. Cell-type specificity scores were then tested as continuous gene-level covariates against MAGMA gene-level association statistics. Analyses were performed independently at the broad cell-class level and neuronal subtype level within each GWAS.

### Cell-state-specific candidate-gene prioritization

To prioritize candidate MDD risk genes within genetically implicated neuronal populations, we integrated gene-level association, transcriptome-wide association and rank-based single-cell expression evidence. Genetic candidate genes were defined as genes supported by MAGMA gene-wise association and overlapping with FUSION TWAS-supported genes. For visualization, genetic association strength was represented using MAGMA-derived FDR values.

For each prioritized neuronal population, we evaluated these genetically supported genes using three expression-ranking criteria. First, cells within the focal population were ranked by their scPagwas trait-relevance score (TRS), and genes were ranked according to their relative expression between cells in the top and bottom 10% of the TRS distribution. Second, genes were ranked by their enrichment in the focal neuronal population relative to other major brain cell classes. Third, genes were ranked by their specificity in the focal population relative to closely related neuronal subtypes. These rank-based expression lists were then intersected with the genetically supported candidate genes to identify candidates with convergent genetic and cell-state-specific expression evidence.

For each gene, a convergent evidence score was calculated by summing support across the genetic evidence layer and the three rank-based expression layers, yielding scores from 1 to 4. This framework was applied to PVALB⁺ inhibitory neurons to prioritize CNNM2-linked inhibitory-neuron programs and to Ex-L2/4 excitatory neurons to evaluate NEGR1-linked excitatory-neuron programs.

### pySCENIC regulon analysis

To infer transcriptional regulatory programs in prioritized neuronal states, we used pySCENIC v0.12.1^78^. Analyses were performed on the single-cell expression matrices of selected neuronal populations, including *CNNM2⁺* versus *CNNM2⁻* PVALB⁺ inhibitory neurons and *NEGR1⁺* versus *NEGR1⁻* Ex-L2/4 excitatory neurons. The pySCENIC workflow consisted of three main steps. First, transcription factor (TF)– target gene co-expression modules were inferred from the gene-by-cell expression matrix. Second, candidate modules were refined by motif-enrichment analysis using *cis*-regulatory motif rankings, retaining only TF–target relationships supported by enriched motifs. The resulting regulons consisted of transcription factors and their predicted direct target genes. Third, AUCell was used to calculate regulon activity scores for each cell by estimating the enrichment of regulon target genes among the highly expressed genes in that cell. For each prioritized neuronal state, regulon activity scores were compared between marker-positive and marker-negative cells within the same neuronal subtype.

### CellChat ligand–receptor analysis

To assess intercellular communication involving prioritized neuronal states, we performed ligand–receptor analysis using CellChat v1.6.1^81^. Normalized single-cell expression matrices and cell-type annotations were used as input. Analyses focused on *CNNM2⁺* versus *CNNM2⁻* PVALB⁺ inhibitory neurons and *NEGR1⁺* versus *NEGR1⁻* Ex-L2/4 excitatory neurons in relation to other annotated brain cell populations. We used the Secreted Signaling category of CellChat resource, which includes secreted, endocrine, and paracrine signalling interactions. CellChat inferred communication probabilities for ligand–receptor pairs based on ligand, receptor and cofactor expression, and pathway-level communication strength was obtained by aggregating significant ligand–receptor interactions within each signalling pathway. We compared the number, strength and pathway composition of predicted interactions between marker-positive and marker-negative neuronal states.

### hdWGCNA co-expression network analysis

To identify co-expression networks associated with prioritized neuronal states, we performed high-dimensional weighted gene co-expression network analysis using hdWGCNA v0.2.24^82^. Analyses were conducted separately for *CNNM2⁺* PVALB⁺ inhibitory neurons, together with their corresponding comparison groups. Because single-cell expression matrices are sparse, we first constructed metacells by aggregating transcriptionally similar cells within the same sample and cell population. Metacell expression matrices were then used to build weighted gene co-expression networks. For each analysis, an appropriate soft-thresholding power was selected to approximate scale-free network topology, and gene modules were identified based on topological overlap of co-expression patterns. Module eigengenes were calculated to summarize the expression pattern of each module across metacells and to evaluate module activity in marker-positive and marker-negative neuronal states.

Modules containing *CNNM2* were selected for downstream characterization. Within each selected module, hub genes were defined according to module connectivity and module membership. Genes from grey or unassigned modules, or genes not associated with the prioritized cell state, were excluded from downstream interpretation. Hub genes from the *CNNM2*-associated modules were subjected to pathway-enrichment analysis to identify biological processes and signalling pathways represented by each co-expression program. Moreover, to examine protein-level interactions among module genes, hub genes were mapped to the human protein– protein interaction network from the STRING (v11.0b) database^83^. Interactions among *CNNM2* and module hub genes were visualized as weighted co-expression and protein-interaction networks.

### Functional enrichment analysis

To characterize biological pathways associated with prioritized neuronal states, we performed functional enrichment analysis using clusterProfiler v4.8.2^103^. Differentially expressed genes were identified between *CNNM2⁺* and *CNNM2⁻* PVALB⁺ inhibitory neurons, and between *NEGR1⁺* and *NEGR1⁻* Ex-L2/4 excitatory neurons. Gene Ontology biological process (GO-BP)^106^ and Kyoto Encyclopedia of Genes and Genomes (KEGG)^104^ enrichment analyses were then performed using the corresponding differentially expressed genes as input. Expressed genes detected in the relevant neuronal population were used as the background gene set where applicable. Enrichment significance was assessed using the hypergeometric test implemented in clusterProfiler, and multiple testing was controlled using the Benjamini–Hochberg false-discovery rate procedure. Pathways with FDR < 0.05 were considered significant and were used for downstream interpretation and visualization.

### scDepBrain web-resource construction

The back end of scDepBrain (https://scdepbrain.su-lab.org/) was implemented in Python using the Django web framework and deployed with Nginx for web serving and request handling. The front-end interface was built using JSP, CSS3, jQuery, Bootstrap and ECharts, enabling interactive visualization of UMAP embeddings, gene-expression profiles, cell-type composition, differential-expression results, pathway enrichment and MDD genetic-prioritization outputs. All processed datasets, metadata tables and precomputed analysis results were stored in SQLite, a lightweight relational database, to support efficient search, visualization and download functions.

### Animals

All experimental animals were adult C57BL/6J mice (6-8 weeks old, weighing 20-25 g) purchased from GemPharmatech Co., Ltd. Prior to experiments, all mice were acclimatized for one week under standard housing conditions with a 12-hour light/dark cycle, temperature maintained at 22 ± 2°C, and relative humidity at 50 ± 10%. All experimental procedures were approved by the Institutional Animal Care and Use Committee (IACUC) of Hangzhou City University and conducted in compliance with national and regional guidelines for animal research. At the start of the experiment, eight mice were randomly divided into two groups: Experimental group (CUMS group, n = 4): Subjected to chronic unpredictable mild stress (CUMS) modeling. Control group (untreated group, n = 4): Received no treatment but underwent the same behavioral assessments.

### Chronic Unpredictable Mild Stress (CUMS) Model

To establish a chronic depression model, this study utilized chronic unpredictable mild stress (CUMS) to induce depressive-like behaviors in mice. During the experiment, the mice in the CUMS group were subjected to a series of unpredictable stressors for 21 days. The specific stressors were included: Cage shaking: Mice from the same group were placed in the same cage, and the shaker speed was set to 100 rpm for 1 hour; Colds water swimming: Mice were placed in a beaker for 5 minutes with water at 4 ± 1°C, 200–300 g of ice was added, and the water volume was 500 ml, ensuring that the mice’s toes could touch the bottom. Restraint: Mice were placed in a transparent restrainer tube for 3 hours, with the opening sealed by cotton to immobilize the mice while allowing normal breathing and preventing asphyxiation. Noise: Mice were placed in an audiovisual stimulation box set to a frequency of 5000 Hz and a volume of 80 dB for 5 minutes. Cage tilt: The cage was tilted at a 45° angle and maintained in this position for 24 hours. Wet bedding: The bedding in the mouse cages was moistened with water, ensuring the water level did not exceed the surface of the bedding, creating a humid environment. Footshocks: Mice were placed in a shock box, and a current of 0.3 mA was applied for 5 seconds for habituation. The electric shock lasted 10 seconds, followed by a 5-second interval, repeated 10 times.

### Behavioral testing

The Open Field Test (OFT) is used to assess the activity levels and anxiety responses in mice. The experiment was conducted in a 50 × 50 × 40 cm open field, which was divided into central and peripheral zones. Each mouse was placed individually in the center of the field, and the test lasted for 10 minutes. The time spent in the central and peripheral zones, as well as the total distance traveled, were recorded. Mice with higher anxiety levels tend to remain in the peripheral zone, while those with lower anxiety levels spend more time in the central zone.

The Tail Suspension Test (TST) is used to assess depressive-like behaviors in mice. Mice were suspended by their tails from a metal frame, maintaining an elevated position for the duration of the test, which lasted 6 minutes. The ratio of immobility time to activity time during the final 4 minutes of the test was recorded, with increased immobility correlating with depressive-like behaviors.

The Forced Swimming Test (FST) is used to assess depressive-like behaviors in mice. Mice were placed in a transparent cylindrical tank filled with water to a depth of approximately 25 cm, with the water temperature maintained at 24 ± 1°C. The test lasted for 6 minutes. The floating time during the last 4 minutes of the test was recorded. An increase in floating time is typically associated with depressive-like behaviors, indicating a state of learned helplessness.

### Immunocytochemistry

Mice were anesthetized with 3% isoflurane, fixed on a dissection board, and a thoracotomy was performed to expose the heart. A 21G needle was inserted into the left ventricle, and pre-chilled PBS was slowly perfused until the effluent became clear. The perfusion was then switched to 4% PFA for continued tissue fixation. After perfusion, the scalp was incised, the skull was removed, and the brain was carefully extracted and fixed overnight in 4% PFA at 4°C. The next day, the brain was transferred to a 15% sucrose solution and incubated overnight. It was then transferred to a 30% sucrose solution, allowing the tissue to fully dehydrate and settle.

The tissue was extracted, embedded in OCT, and rapidly frozen at -80°C. Using an Epredia cryostat, the tissue was sectioned into 30 μm coronal brain slices and transferred to pre-chilled cryoprotectant (PBS, ethylene glycol, and glycerol mixture) at -20°C for storage. Prior to immunofluorescence staining, brain slices were removed from the cryoprotectant and washed three times with PBS (10 minutes per wash). The slices were then blocked at room temperature for 12 minutes using the Beyotime Quick Block (Beyotime, P0260), which both permeabilizes and blocks the tissue. The brain slices were incubated overnight at 4 ° C in a solution of specific primary antibodies (Cnnm2, Thermofisher; Arhgef7, Proteintech; PV, Beyotime; Somatostatin, Thermofisher). The following day, the slices were washed three times with PBS and then incubated for 1 hour at room temperature with secondary antibodies conjugated to Alexa Fluor 488 or Alexa Fluor 594, protected from light. After washing, the slices were mounted and coverslipped using a Beyotime anti-fade mounting medium with DAPI (Beyotime, P0131) for fluorescence preservation. Imaging and photography of tissue sections were performed using the 3DHISTECH Pannoramic MIDI scanner.

### Stereotaxic Surgery

Male mice were anesthetized in an induction chamber with isoflurane and then head-fixed in a stereotaxic frame (RWD Instruments). During the surgery, the isoflurane concentration was maintained at 1.5%, and the mice were kept warm on heating pads. A small hole was drilled in the skull at the target brain region (anterior spot coordinates (mm): primary motor cortex (M1, AP: 0.25 mm anterior from the bregma, ML: ±1.22 mm from the midline, DV: -1.4 mm from the dura); lateral/ventrolateral periaqueductal gray matter in the midbrain (lPAG/vlPAG, AP: -4.6 mm from the bregma, ML: ±0.6 mm from the midline, DV: -2.6 mm from the dura). For mice receiving viral microinjection, a glass pipette connected to a pressure microinjector (RWD) was used to inject the virus at a rate of 70 nL/min. For mice receiving an intracranial implant (optogenetic fiber), the implant was slowly lowered to the target site and fixed to the skull using bone screws and dental adhesive (New century dental).

For the inhibition of the M1 downstream pathway, 200 nL of AAV2/9-hSyn-eNpHR3.0-EYFP (titer: 1.76 × 10^12^ v.g./ml) or the control virus AAV2/9-hSyn-ZsGreen was bilaterally injected into M1 (AP: 0.25 mm anterior from the bregma, ML: ±1.22 mm from the midline, DV: -1.4 mm from the dura), and a double fiber cannula (NA = 0.37, Inper) was implanted in its downstream brain region, lPAG/vlPAG (AP: -4.6 mm anterior from the bregma, ML: ±0.6 mm from the midline, DV: -2.5 mm from the dura).

For the activation of the M1 downstream pathway, 200 nL of rAAV2/9-EFIα-DIO-hChR2(HI34R)-mCherry-WPRE-hCH polyA (titer: 5.08 × 10^12^ vg/mL) or the control virus rAAV2/9-EFIα-DIO-mCherry-WPRE-hCH polyA (titer: 5.18 × 10^12^ vg/ ml) was bilaterally injected into the M1 of PV-Cre mice; 200 nL of rAAV2/9-CaMKIIα-hChR2(HI34R)-mCherry-WPRE-hCH polyA (titer: 5.15 × 10^12^ vg/mL) or the control virus PT-0108 rAAV2/9-CaMKIIα-mCherry-WPRE-hCH polyA (titer: 5.29 × 10^12^ vg/ mL) was bilaterally injected into the M1 of wild-type (WT) mice (AP: 0.25 mm anterior from the bregma, ML: ±1.22 mm from the midline, DV: -1.4 mm from the dura), and a double fiber cannula (NA = 0.37, Inper) was implanted into their downstream brain region, lPAG/vlPAG (AP: -4.6 mm anterior from the bregma, ML: ±0.6 mm from the midline, DV: -2.5 mm from the dura).

For anterograde tracking experiments, 200 nL of rAAV2/9-hSyn-mCherry-WPRE-hCH polyA (titer: 5.15 × 10^12^ vg/mL) was injected into the right M1 (AP: 0.25 mm anterior from the bregma, ML: -1.22 mm from the midline, DV: -1.4 mm from the dura). For retrograde tracing experiments, 50 nL of AAV2-Retro-hsyn-EGFP (titer: 1 × 10^13^ vg/mL) was unilaterally injected into the right lPAG/vlPAG (AP: -4.6 mm anterior from the bregma, ML: -0.6 mm from the midline, DV: -2.6 mm from the dura).

### Anterograde and Retrograde Tracing

For anterograde tracking experiments, 3 weeks after injecting AAV2/9-hSyn-mCherry into M1, the mice were perfused. The brains were placed in 4% paraformaldehyde (PFA) overnight and then incubated in 20% and 30% sucrose (dissolved in PBS) sequentially until the samples sank. Coronal brain sections with a thickness of 30 μm were cut using a Cryosectioner (NX50, Thermo). The sections were mounted on slides and sealed using an anti-fluorescence quenching sealer containing DAPI (Beyotime Biotechnology). Imaging was performed using a SpinSR turntable confocal microscope (Evident/SpinSR).

For retrograde tracking experiments, 3 weeks after injecting AAV2-Retro-hsyn-EGFP into lPAG/vlPAG, the mice were perfused. The samples were placed in 4% PFA overnight and then incubated in 20% and 30% sucrose (dissolved in PBS) sequentially until the samples sank. Coronal brain sections (of lPAG/vlPAG and M1 brain regions) with a thickness of 30 μm were cut using a Cryosectioner (NX50, Thermo). The sections were mounted on slides and sealed using an anti-fluorescence quenching sealer containing DAPI (Beyotime Biotechnology). Imaging was performed using a SpinSR turntable confocal microscope (Evident/SpinSR).

### Chronic Restraint Stress (CRS)

For optogenetic circuit experiments, mice were subjected to chronic restraint stress (CRS) by placement in 50-ml conical tubes for 6-7 h per day for 21 days. After CRS, mice underwent optogenetic manipulation during behavioural testing as described below.

### Optogenetic Manipulations

The light output was measured with an optical power meter and adjusted to 5 mW for 473 nm light or 10 mW for 590 nm light. For mice expressing eNpHR3.0 and eGFP, a 589 nm yellow light stimulus was continuously delivered throughout the test period with a light intensity of 10 mW. For mice expressing ChR2 and mCherry, a 473 nm blue light laser was used, with a light intensity of 5 mW, 5 ms pulses of light stimulation, and a frequency of 20 Hz, following a pattern of 10 s on and 2 s off.

(1) Rotarod Running: To assess the effects of general and challenging exercise on depression, a commercially available rotarod (Ugo Basile) was used, with the rotarod set at a constant speed of 10 rpm. The animals were subjected to six constant rotarod running trials per day, with a time limit of 240 s per trial and a rest interval of 240 s to avoid stress and fatigue. For the control group, mice were placed in a separate compartment of the rotarod, which remained stationary. For optogenetic manipulations, the laser stimulus was delivered as soon as the mice were placed on the rotarod to start locomotion and turned off when locomotion stopped.
(2) Forced Swim Test (FST): Mice were acclimatized in a test room for 1 h before the experiment. They were individually placed in transparent glass cylinders (45 cm high and 19 cm in diameter) containing water at a height of 23 cm (temperature 23-25°C). The mice swam for 6 min under normal light conditions. The water depth was set to prevent the animal’s tail and hind limbs from touching the bottom. Animal behavior was recorded from the front. The immobility time during the last 4 min of the trial was measured using a tracking system (Ethovision XT, Noldus Information Technology, Netherlands). Increased immobility in mice was used to measure behavioral despair, whereas decreased immobility was considered to indicate an antidepressant effect. For optogenetic manipulations, laser stimuli were delivered immediately after placing the mice in water and continued for 6 min. To minimize the impact of the optogenetics cable on swimming behavior, the cable length was adjusted so that the cable just touched the water surface.
(3) Sucrose Preference Test (SPT): The animals were housed individually and habituated to two bottles of water for 2 days, followed by one bottle of water and one bottle of 1% sucrose solution for 2 days, with the positions of the two bottles exchanged every 24 h. At the end of the acclimatization period, the animals were water-fasted for 12 h. For the formal test, the mice were given one bottle of water and one bottle of 1% sucrose solution for 1 day, with the positions of the two bottles exchanged at 12 h. Each bottle was weighed before and after the test, and the weight difference was taken as the amount ingested by the mice from each bottle. The preference for the sucrose solution was expressed as the percentage of sucrose intake relative to the total intake (sum of water and sucrose intake). For optogenetic manipulations, the mice were placed in cages with normal bedding and allowed to move freely, and photostimulation was provided during the first hour of the 24-hour test.

### Immunofluorescence

Mice were deeply anesthetized with isoflurane and then perfused with ice-cold PBS and 4% PFA. The brains were removed, placed in 4% PFA overnight, and then incubated in 20% and 30% sucrose (dissolved in PBS) sequentially until the samples sank. Coronal brain sections with a thickness of 30 μm were cut using a Cryosectioner (NX50, Thermo). The sections were washed 3 times in PBS for 5 min each; permeabilized with 0.5% Triton X-100 for 15 min; washed 3 times with PBS; and then incubated with 5% normal goat serum at room temperature for 1 h. They were then incubated with primary antibodies diluted in PBS, including anti-CaMKII (1:200, rabbit, ThermoFisher), anti-c-Fos (1:1000, rabbit, Cell signaling technology), anti-PV (1:800, mouse, Merck), and anti-mCherry (1:800, rabbit, Proteintech) at 4°C overnight. The next day, the sections were removed and rewarmed at room temperature for 30 min, then washed three times with PBS and incubated with appropriate secondary antibodies (1:1000, Thermo Fisher Scientific) for 1 h at room temperature in the dark. After that, the sections were sealed using an anti-fluorescence quenching sealer containing DAPI (Beyotime Biotechnology). Images were captured using a SpinSR turntable confocal microscope (Evident/SpinSR). Regions of interest in the brain slices were selected by manually drawing contours based on brain mapping, and the upper and lower thresholds were adjusted to match the fluorescence.

### Statistical Analysis

Statistical analyses were performed using R, Python and GraphPad Prism 9.5. For single-cell differential-expression analyses, Wilcoxon rank-sum tests were performed using Seurat, with Benjamini–Hochberg FDR correction. GO-BP and KEGG enrichment analyses were performed using hypergeometric tests in clusterProfiler, and pathways with FDR < 0.05 were considered significant. Unless otherwise specified, multiple testing across genes, cell types, pathways, regulons or interaction pairs was controlled using Benjamini–Hochberg FDR correction. For cell-level comparisons of trait-relevance scores, regulon activities, gene-expression levels, pathway scores and predicted ligand–receptor interactions, two-sided Wilcoxon rank-sum tests were used unless otherwise stated. For *in vivo* behavioural, immunofluorescence and optogenetic experiments, data were analyzed using GraphPad Prism 9.5 and are presented as mean ± s.e.m. Two-group comparisons were performed using unpaired two-tailed Student’s t tests or Mann–Whitney tests, as appropriate; comparisons involving more than two groups or multiple conditions were analyzed using one-way or two-way ANOVA followed by post hoc multiple-comparison tests. Exact tests, sample sizes and significance thresholds are reported in the figure legends. P < 0.05 was considered statistically significant.

## Funding

This study was funded by the National Natural Science Foundation of China (32200535 to Y.M; 82401786 to H.H; 61871294 and 82172882 to J.S), the Scientific Research Foundation for Talents of Wenzhou Medical University (KYQD20201001 to Y.M.), Natural Science Foundation of Zhejiang Province (LQ24C090002 to H.H), and the Natural Science Foundation of Zhejiang Province (LR19C060001 to J.S).

## Author Contributions

Y.M. and J.S. conceived and designed the study. Y.M., C.C., W.D., F.Q., J.L., Y.Z., D.J., G.Z., Z.Z., C.D., Y.H., C.J., J.,E., D.V., and J.Z. contributed to management of data collection. X.H., C.G., H.H., and X. C. contributed to mice model construction and *in vivo* experiments. Y.M., Y.J.Z., W.D., and J.L. conducted bioinformatics analysis and data interpretation. C.C., and W.D. wrote the code of the website and database of scDepBrain. Y.M., M.G. and J.S. wrote and revised the manuscripts. All authors reviewed and approved the final manuscript.

## Data availability

GWAS summary statistics used in this study are publicly available from the MRC IEU OpenGWAS database (https://opengwas.io/) and the PGC MDD 2025 data release (https://doi.org/10.6084/m9.figshare.27061255). The OpenGWAS datasets included the major depression GWAS (ID: ieu-b-102) and the standing-height negative-control GWAS (ID: ukb-b-10787). Updated PGC MDD 2025 European-ancestry and trans-ancestry summary statistics were obtained from the PGC MDD 2025 release. Public human and mice brain single-cell and single-nucleus transcriptomic datasets were collected from GEO, Allen Brain Map, PsychENCODE, UCSC Cell Browser, NeMO Archive, Figshare, GitHub repositories, and other public resources. All human cerebral organoid single-cell datasets were downloaded from scHBO database (https://schob.su-lab.org/). These datasets comprise the adult human brain discovery atlas and five independent replication atlases spanning adult human brain, developing human brain, mouse brain, cerebral organoids and human MDD case–control brain tissue, with detailed accessions and source links provided in Supplementary Tables 1–5. Processed MDD-associated cell-type prioritization results, trait-relevance scores, gene-expression patterns and pathway-enrichment results are available through scDepBrain (https://scdepbrain.su-lab.org/). Source data underlying the figures are provided with this paper.

## Code availability

The code to reproduce the results is accessible on the Github repository (https://github.com/Ma-brain-group/scDepBrain).

## Competing Interests

The authors declare no competing interests.

## Notes

### Competing Interest Statement

The authors have declared no competing interest.

