## Supplementary Figures for "Human genetic risk for major depressive disorder implicates motor cortex PVALB⁺ inhibitory neurons"

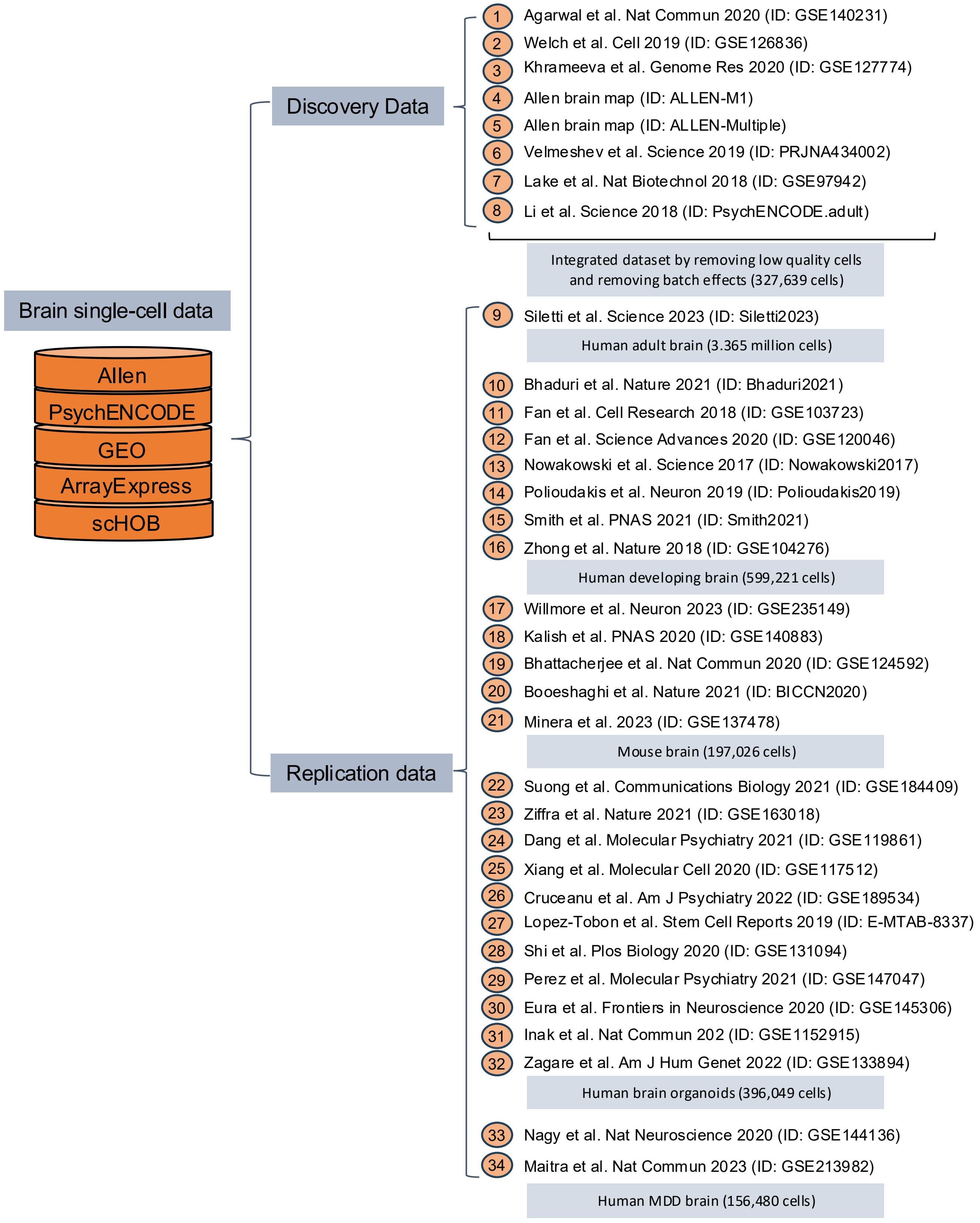


**Supplementary Figure 1 | Overview of data collection and integration for the brain cell atlas, related to Figure 1.**
In total, we collected 34 independent brain single-cell/nuclei datasets from public databases, including Allen, PsychENCODE, GEO, ArrayExpress, and scHOB. These datasets were categorized into four groups based on species origin (human, mouse, or organoid), brain developmental stage (adult or developing), and health status (disease or healthy). The datasets were further classified into one discovery dataset and five replication datasets. The publication years of these datasets range from 2017 to 2023. For more detailed information on these datasets, please refer to the Supplementary Tables 1-3.

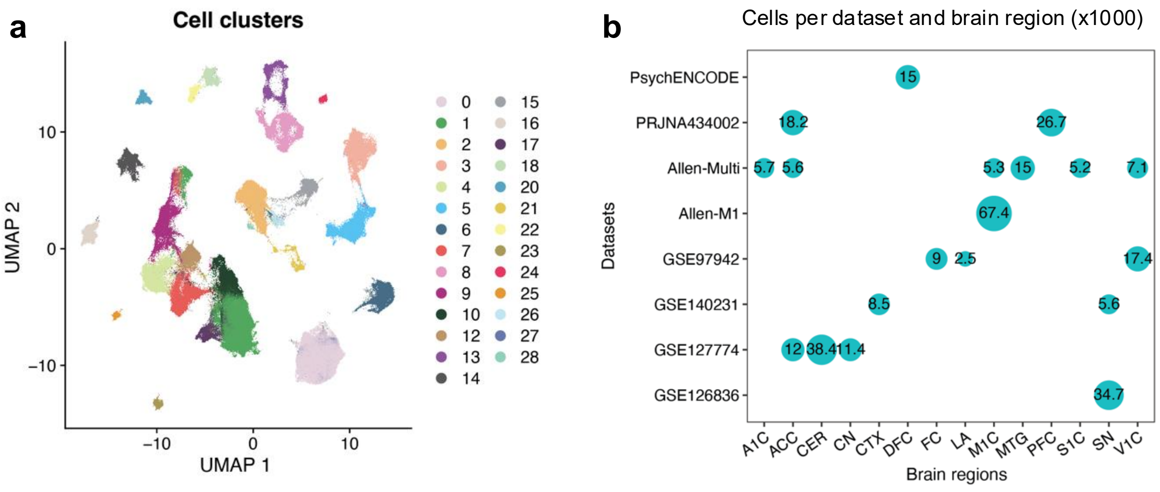


**Supplementary Figure 2 | Distribution and clustering of discovery-set single-cell/nucleus profiles, related to Figure 2.**

a. UMAP embedding of 327,639 high-quality single-cell/nucleus profiles from the adult-human discovery atlas, colored by the 29 clusters identified after integration of eight published datasets.

b. Distribution of cells across source datasets and anatomical regions. Dot size denotes the number of cells for each dataset–region combination, scaled by 1,000 cells.

Note: the discovery atlas comprised cells or nuclei from 14 anatomical regions: anterior cingulate cortex (ACC), caudate nucleus (CN), cortex (CTX), frontal cortex (FC), cerebellar cortex (CER), substantia nigra (SN), primary somatosensory cortex (S1C), primary motor cortex (M1C), dorsolateral prefrontal cortex (DFC), prefrontal cortex (PFC), middle temporal gyrus (MTG), primary auditory cortex (A1C), primary visual cortex (V1C) and lateral amygdala (LA). After UMAP embedding and graph-based clustering using highly variable genes, 29 clusters were identified. Two low-quality clusters were removed, and the remaining 27 clusters comprising 310,833 cells were used for downstream cell-type annotation. Dataset details are provided in Supplementary Table 1.

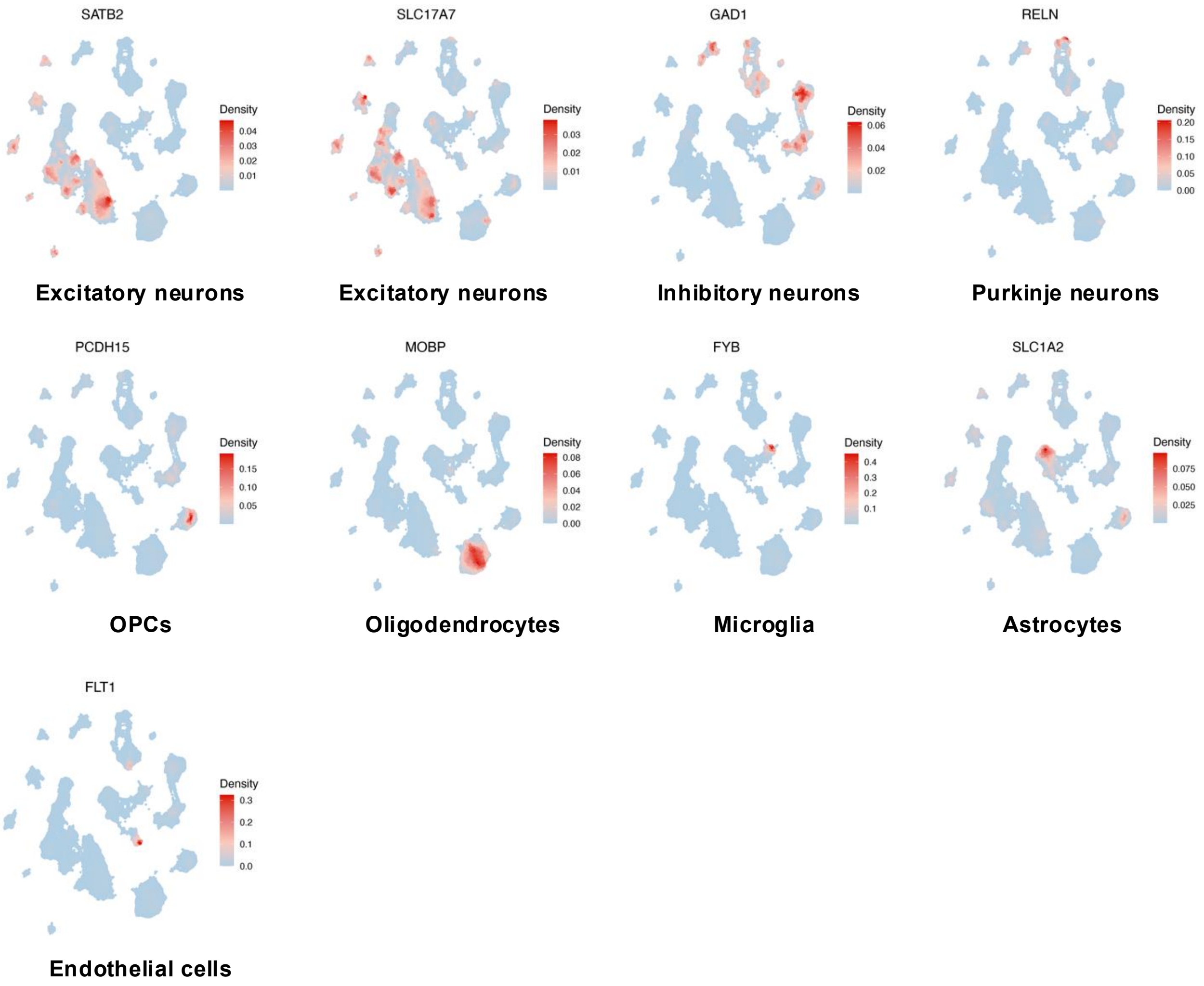


**Supplementary Figure 3 | Scaled expression of representative marker gene for all brain cells in the discovery dataset, related to Figure 2**.

Based on brain cells from eight independent studies, we integrated and yielded 327,639 high-qualified cells and subjected them to a UMAP embedding plot based on highly variable genes using the Seurat.

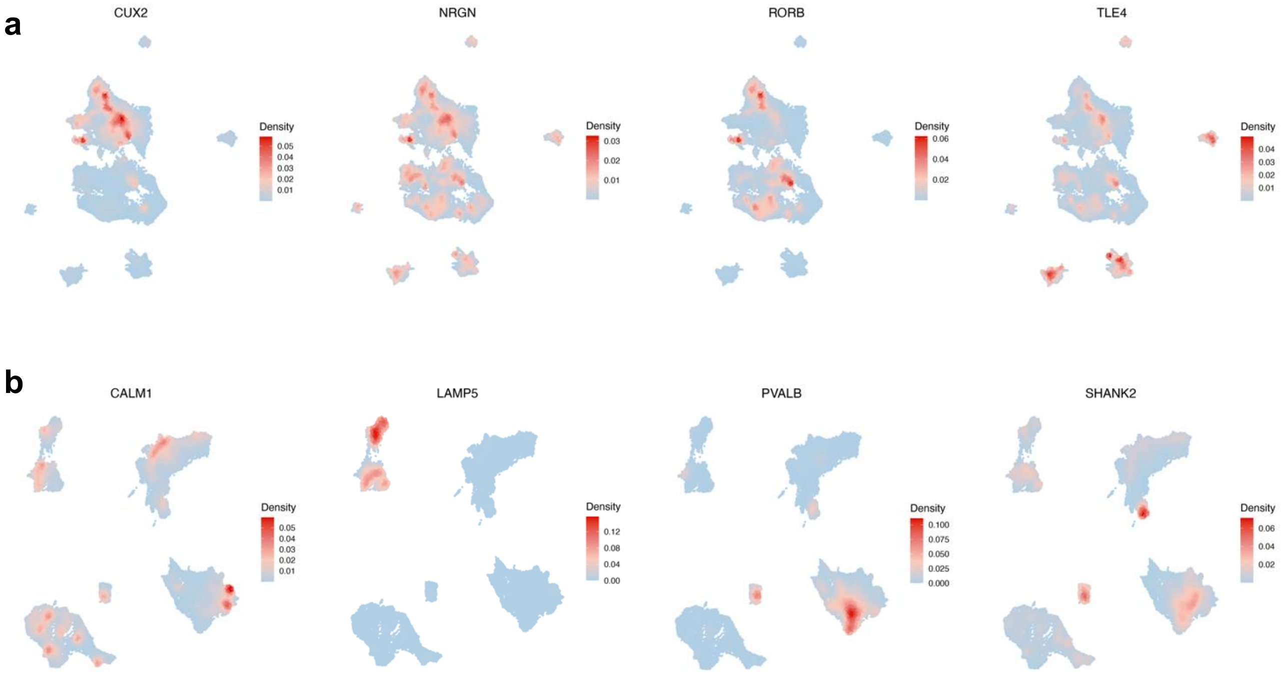


**Supplementary Figure 4 | Marker-gene expression validates neuronal subtype annotations in the discovery atlas, related to Figure 2**.

a,b. UMAP density plots showing scaled expression of representative marker genes for excitatory neuronal subtypes (a) and inhibitory neuronal subtypes (b) in the adult human brain discovery atlas. Excitatory-neuron markers include *CUX2*, *NRGN*, *RORB* and *TLE4*. Inhibitory-neuron markers include *CALM1*, *LAMP5*, *PVALB* and *SHANK2*. Colour indicates scaled expression density across the UMAP embedding.

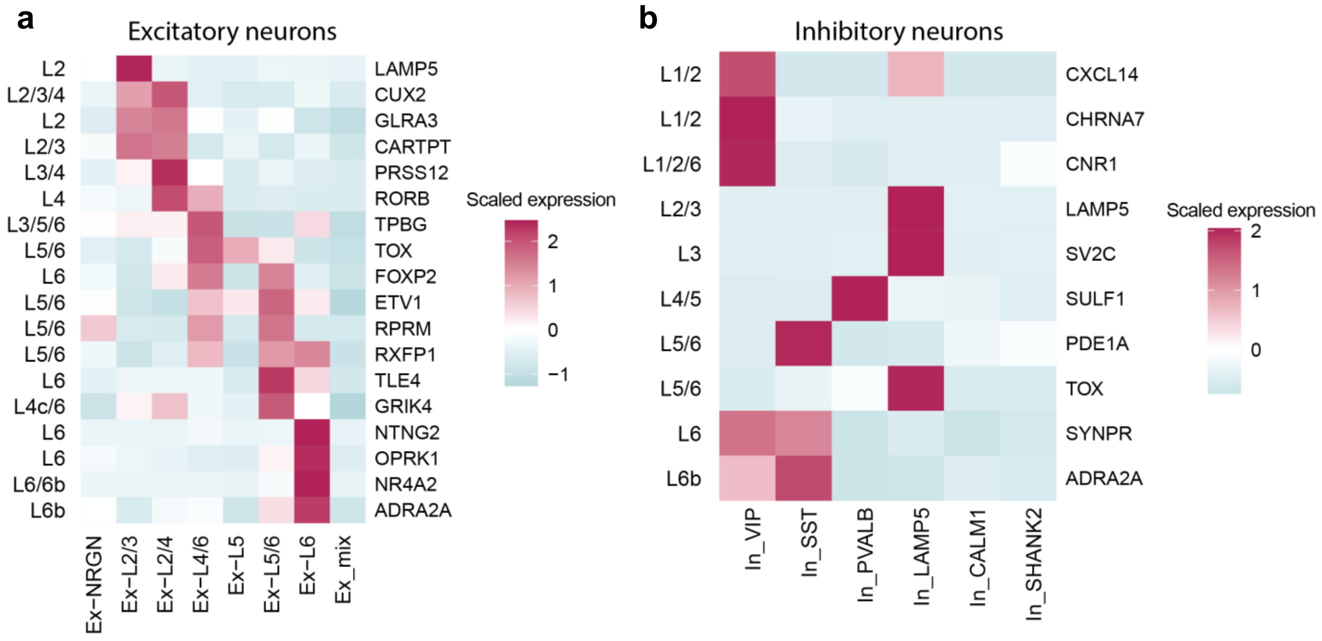


**Supplementary Figure 5 | Layer-enriched marker expression supports excitatory and inhibitory neuronal subtype annotations, related to Figure 2.**

a,b. Heatmaps showing scaled expression of layer-enriched and subtype-enriched marker genes across excitatory neuronal subtypes (a) and inhibitory neuronal subtypes (b) in the adult human brain discovery atlas. Rows indicate marker genes with reported layer or subtype enrichment, and columns indicate annotated neuronal subtypes. Colour represents scaled average expression.

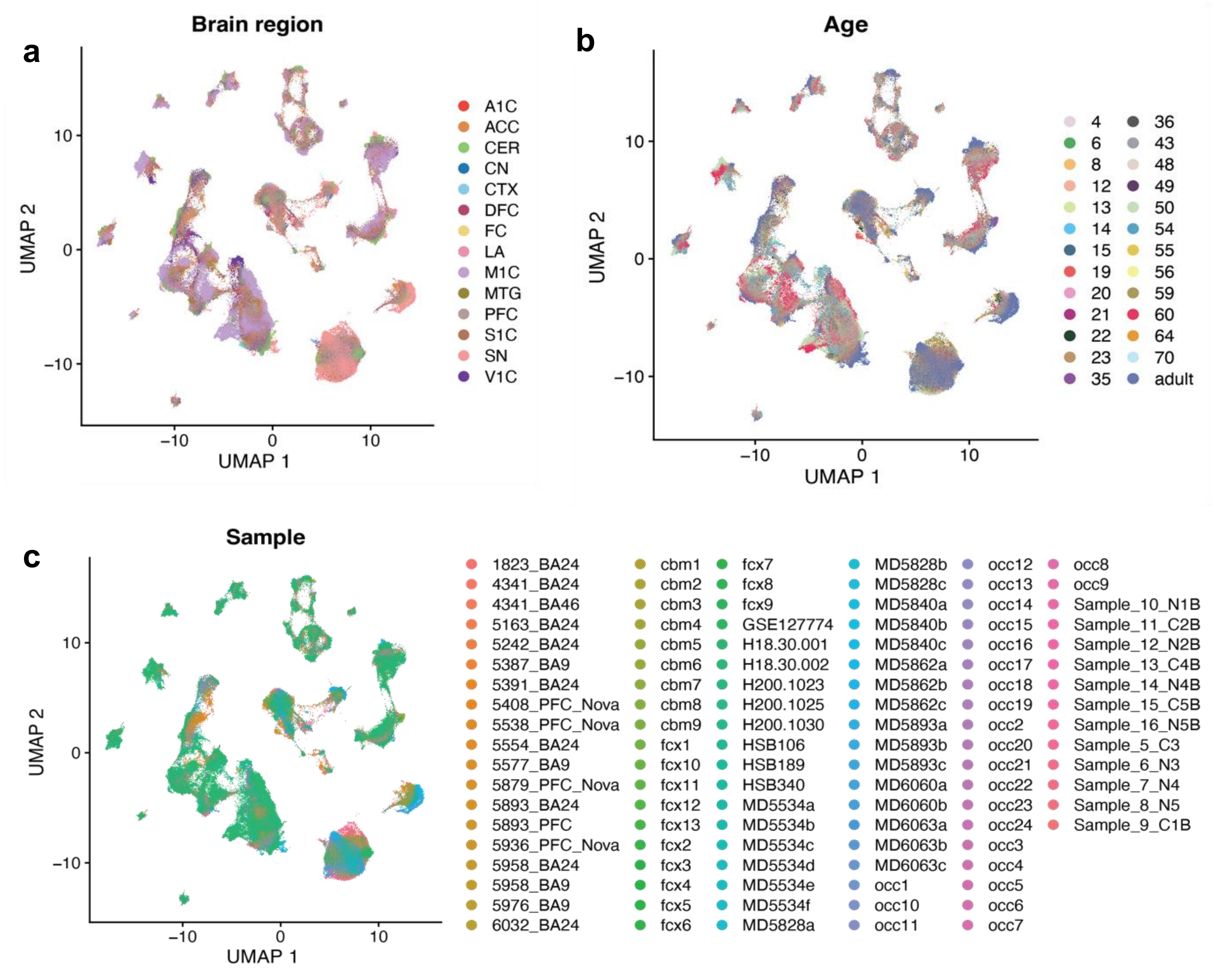


**Supplementary Figure 6 | Integrated UMAP visualization of the adult human brain discovery atlas, related to Figure 2.**

a–c, UMAP embeddings of cells and nuclei in the integrated adult human brain discovery atlas, coloured by anatomical brain region (a), donor age (b) and sample identity (c). Cells from multiple brain regions, ages and samples were jointly embedded after preprocessing and integration. Abbreviations: ACC, anterior cingulate cortex; A1C, primary auditory cortex; CER, cerebellar cortex; CN, caudate nucleus; CTX, cortex; DFC, dorsolateral prefrontal cortex; FC, frontal cortex; LA, lateral amygdala; M1C, primary motor cortex; MTG, middle temporal gyrus; PFC, prefrontal cortex; S1C, primary somatosensory cortex; SN, substantia nigra; V1C, primary visual cortex. Detailed dataset and sample information is provided in Supplementary Table 1.

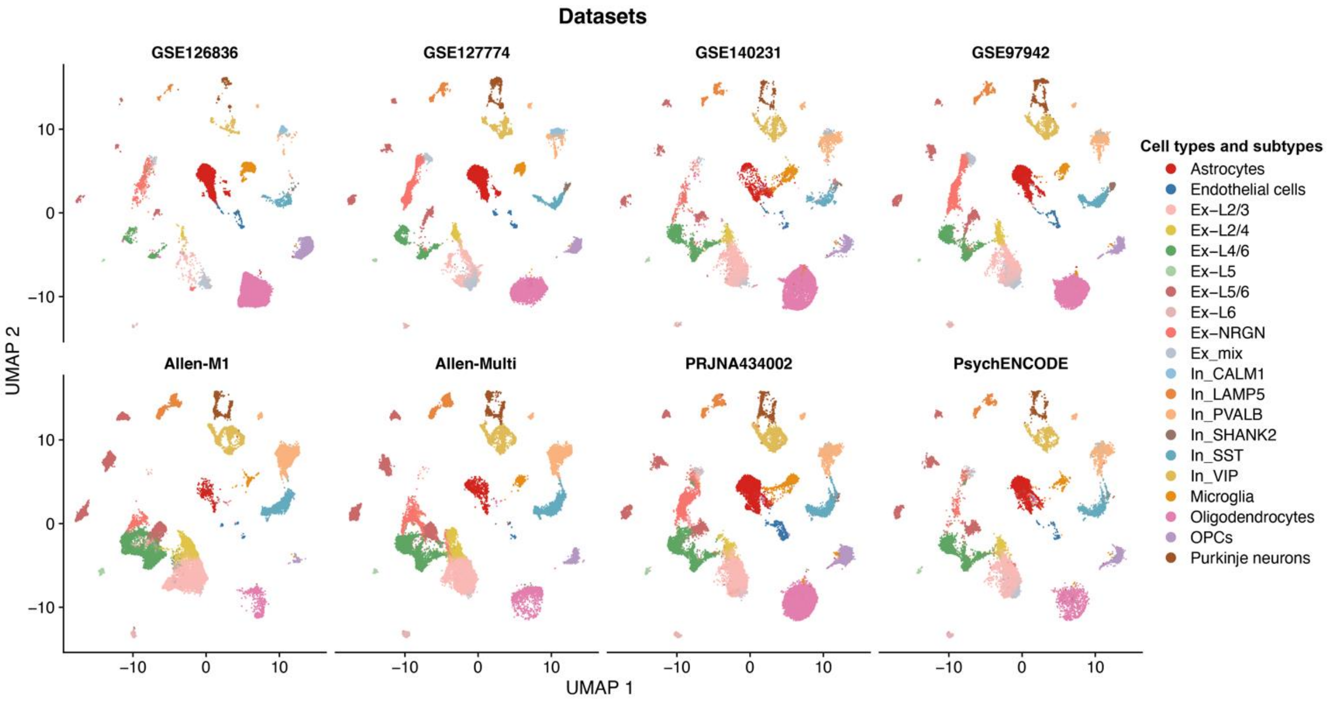


**Supplementary Figure 7 | Cell-type and subtype composition of individual datasets in the adult human brain discovery atlas, related to Figure 2.**
UMAP embeddings of cells and nuclei from each of the eight datasets included in the adult human brain discovery atlas, coloured by annotated major cell type or neuronal subtype. Each panel shows cells from one source dataset projected onto the integrated UMAP space. Colours denote annotated cell classes and neuronal subtypes. Detailed dataset information, including accession numbers, brain regions, sample ages and sequencing platforms, is provided in Supplementary Table 1.

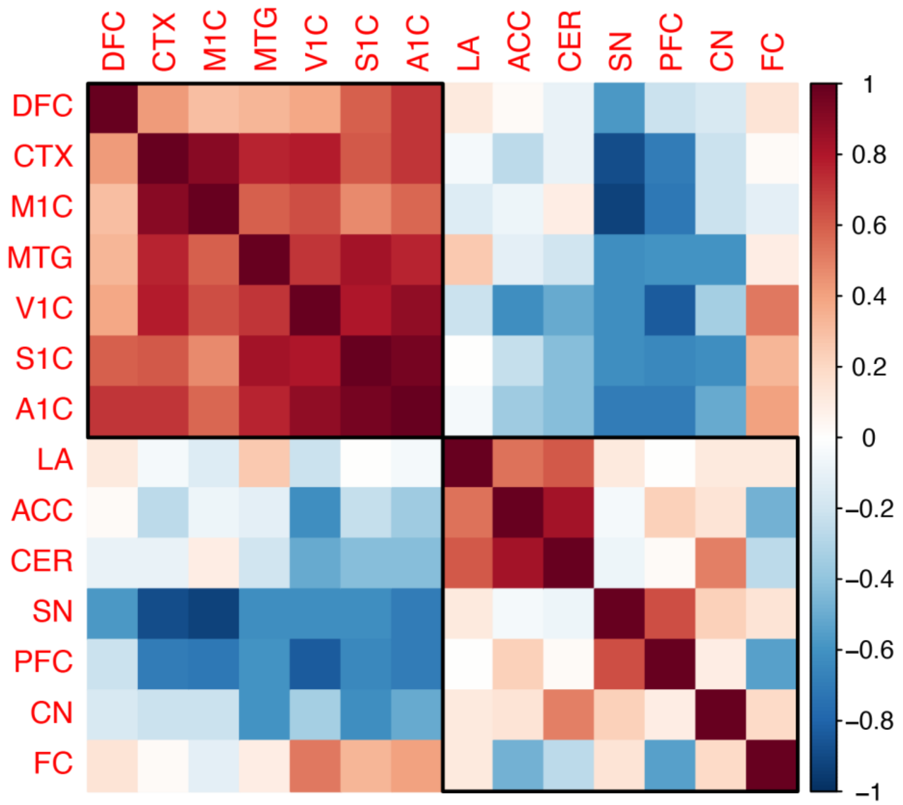


**Supplementary Figure 8 | Regional similarity based on cell-type composition in the adult human brain discovery atlas, related to Figure 2.**
Heatmap showing pairwise Pearson correlation coefficients between anatomical brain regions based on the proportions of annotated major cell types and neuronal subtypes. Hierarchical clustering groups regions with similar cellular composition. Warmer colours indicate higher positive correlations, whereas cooler colours indicate lower or negative correlations. Abbreviations: ACC, anterior cingulate cortex; A1C, primary auditory cortex; CER, cerebellar cortex; CN, caudate nucleus; CTX, cortex; DFC, dorsolateral prefrontal cortex; FC, frontal cortex; LA, lateral amygdala; M1C, primary motor cortex; MTG, middle temporal gyrus; PFC, prefrontal cortex; S1C, primary somatosensory cortex; SN, substantia nigra; V1C, primary visual cortex. Detailed dataset information is provided in Supplementary Table 1.

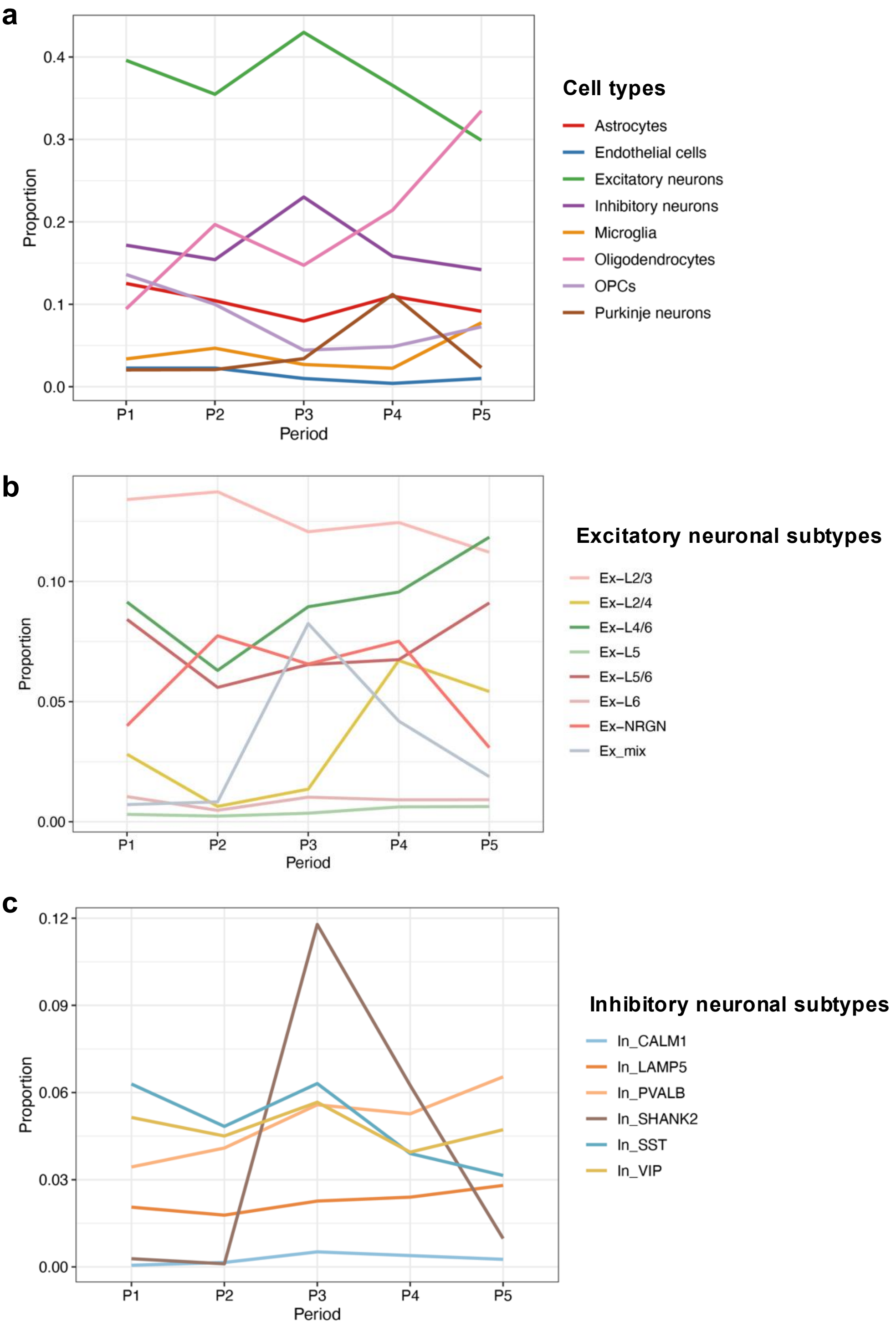


**Supplementary Figure 9 | Postnatal changes in cell-type and neuronal-subtype composition in the adult human brain discovery atlas, related to Figure 2.**
a–c. Line plots showing the proportions of major brain cell types (a), excitatory neuronal subtypes (b) and inhibitory neuronal subtypes (c) across five postnatal age periods in the discovery atlas. Age periods were defined as P1, 4–8 years; P2, 12–19 years; P3, 20–39 years; P4, 40–59 years; and P5, 60 years and older. Proportions were calculated as the fraction of cells assigned to each annotated cell type or subtype within each age period.

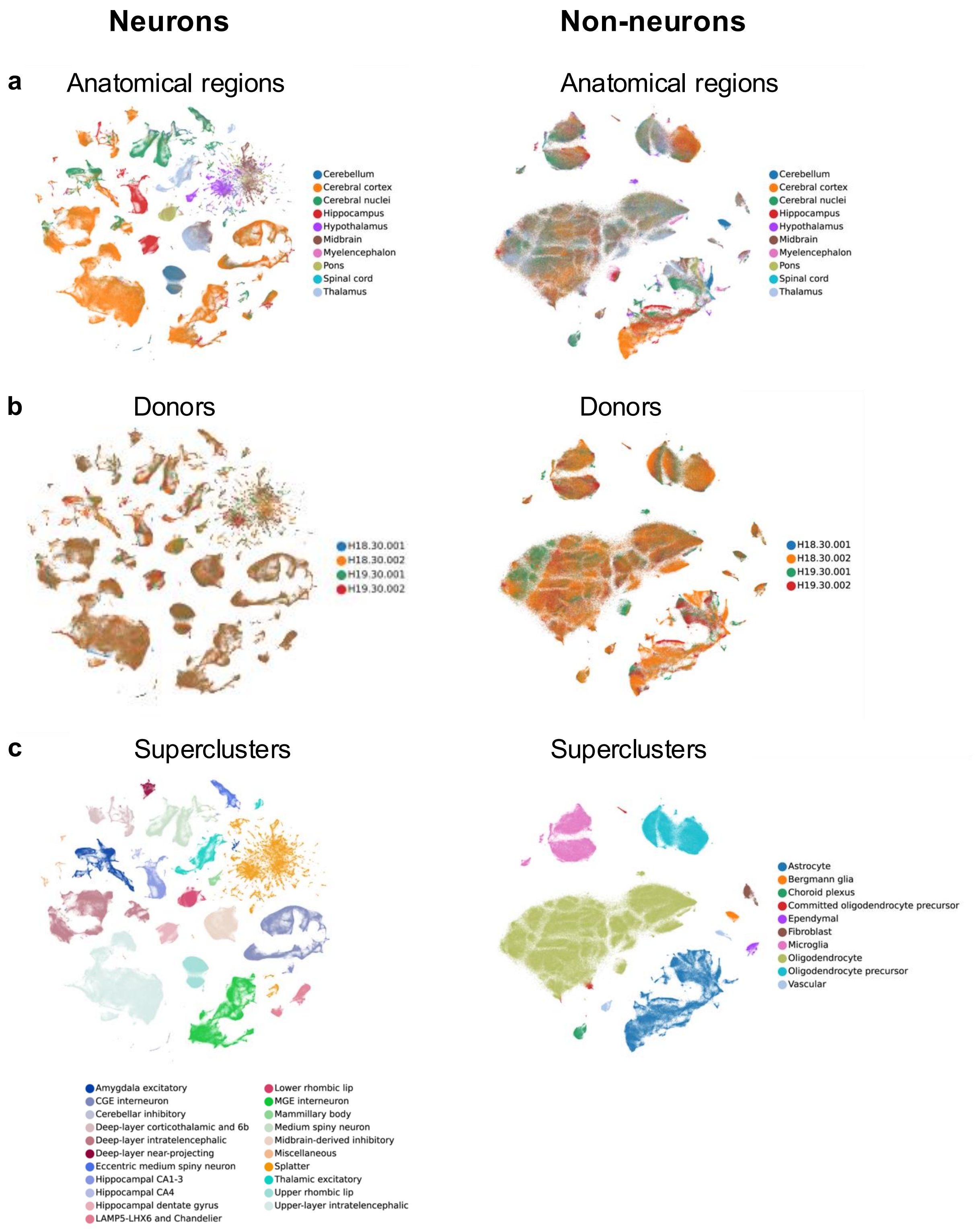


**Supplementary Figure 10 | Cell-type structure of the independent adult human brain replication atlas, related to Figure 2.**

a–c, t-SNE embeddings of neuronal cells and non-neuronal cells in the adult human brain replication atlas (n = 3,369,219 cells), coloured by anatomical region (a), donor (b) and annotated supercluster (c). Left panels show neuronal populations and right panels show non-neuronal populations. Detailed dataset and sample information is provided in Supplementary Table 2.

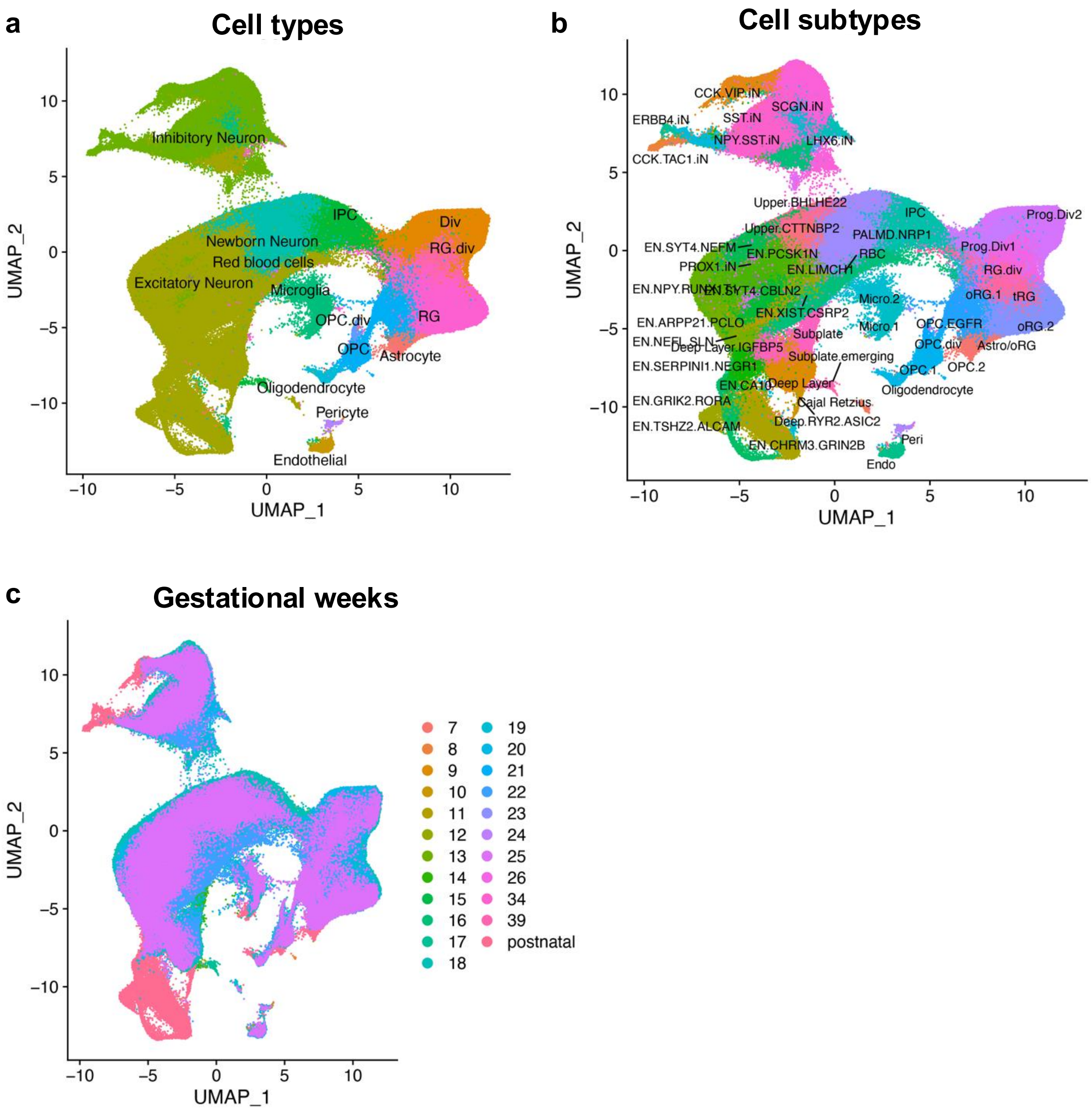


**Supplementary Figure 11 | Cell-type structure and developmental coverage of the human developing brain replication atlas, related to Figure 2.**
a–c, UMAP embeddings of cells and nuclei in the human developing brain replication atlas, coloured by major cell type (a), annotated cell subtype (b) and gestational or postnatal age (c). This replication atlas comprised 599,221 high-quality cells and nuclei spanning fetal developmental stages and postnatal samples. Detailed dataset and sample information is provided in Supplementary Table 2.

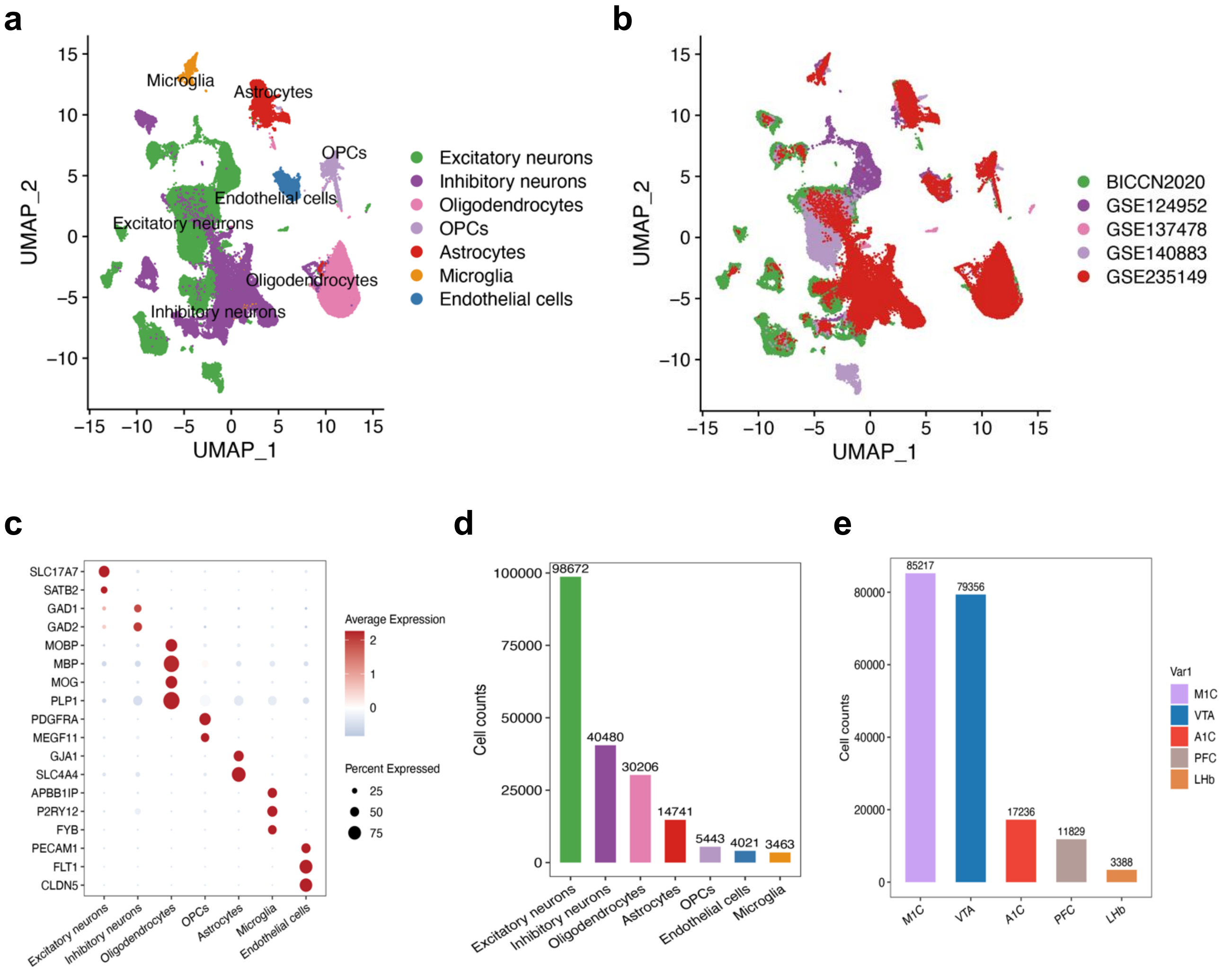


**Supplementary Figure 12 | Overview of the integrated mouse brain replication atlas, related to Figure 2.**

a,b. UMAP embeddings of 197,026 high-quality cells from the integrated mouse brain replication atlas, coloured by major cell type (a) and source dataset (b).

c. Dot plot showing representative marker-gene expression across annotated major cell types. Dot size denotes the percentage of cells expressing each gene, and colour indicates scaled average expression.

d. Bar plot showing the number of cells assigned to each major cell type in the integrated mouse brain atlas.

e, Bar plot showing the distribution of cells across annotated brain regions in the mouse brain atlas. Detailed dataset and sample information is provided in Supplementary Table 2.

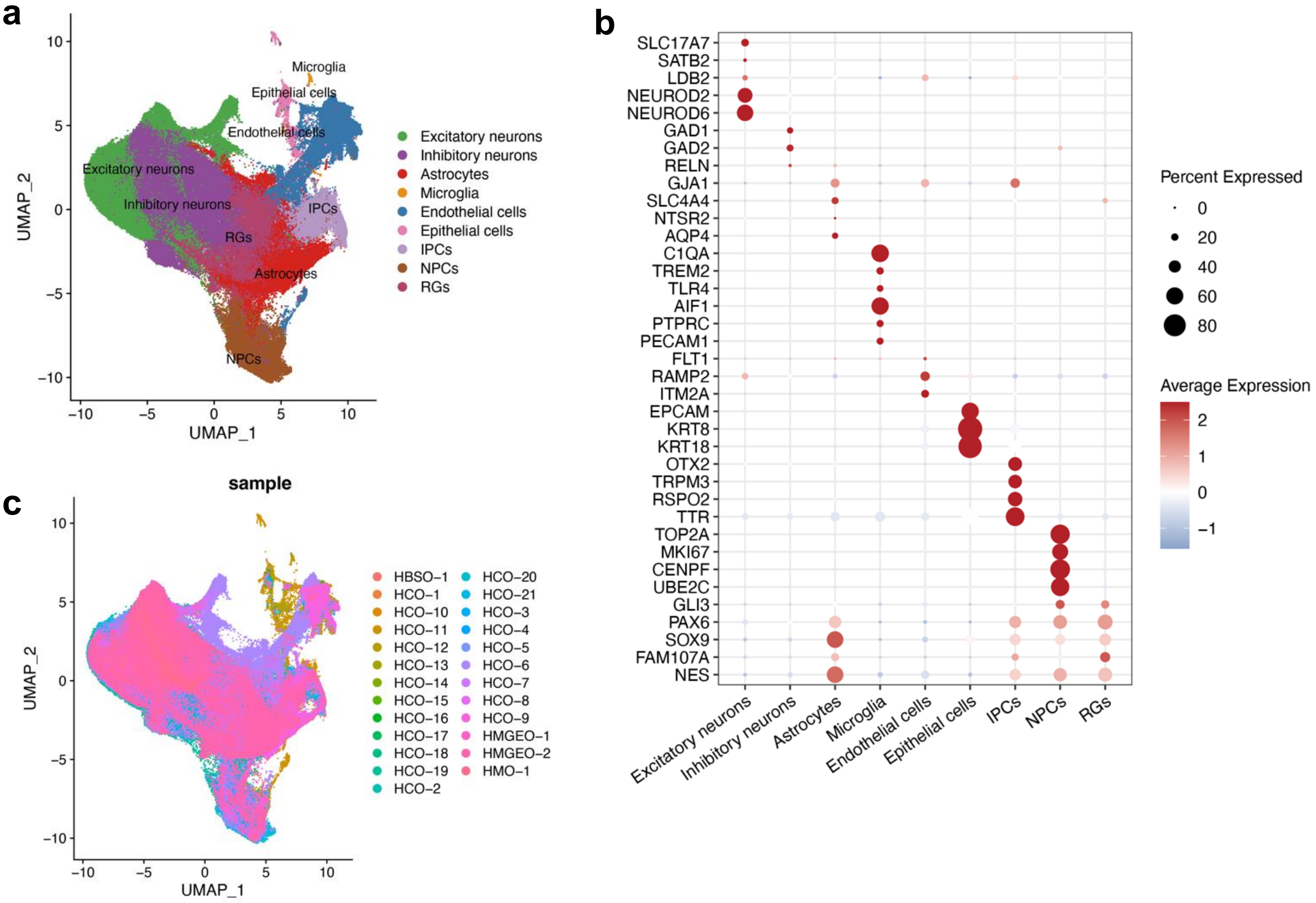


**Supplementary Figure 13 | Overview of the integrated human brain organoid replication atlas, related to Figure 2.**

a,c. UMAP embeddings of 396,049 high-quality cells from the integrated human brain organoid replication atlas, coloured by annotated major cell type (a) and sample identity (c).

b. Dot plot showing representative marker-gene expression across annotated cell types in the integrated brain organoid atlas. Dot size denotes the percentage of cells expressing each gene, and colour indicates scaled average expression. Detailed dataset and sample information is provided in Supplementary Table 3.

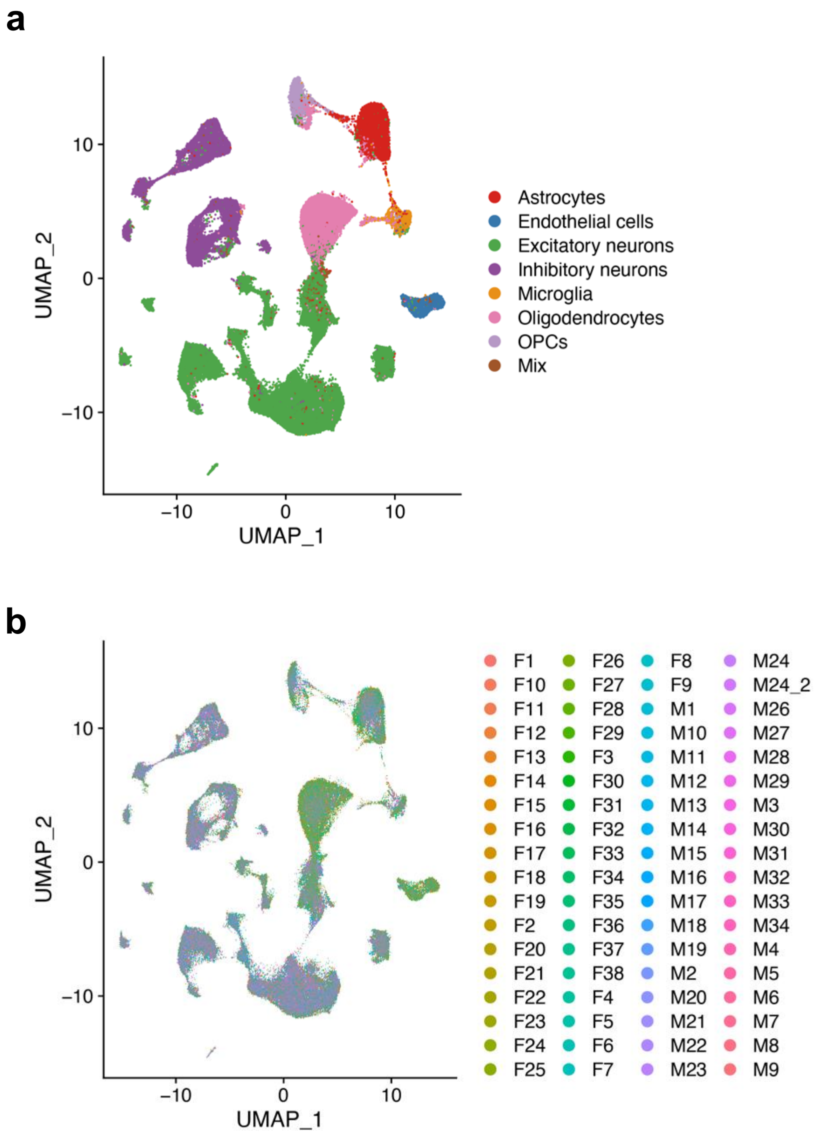


**Supplementary Figure 14 | UMAP visualization of cell types and samples in the replication dataset 5, related to Figure 2.**

a. UMAP exhibiting 160,222 high-quality cells colored by seven cell types.

b. UMAP showing the distribution of cells colored by different samples.

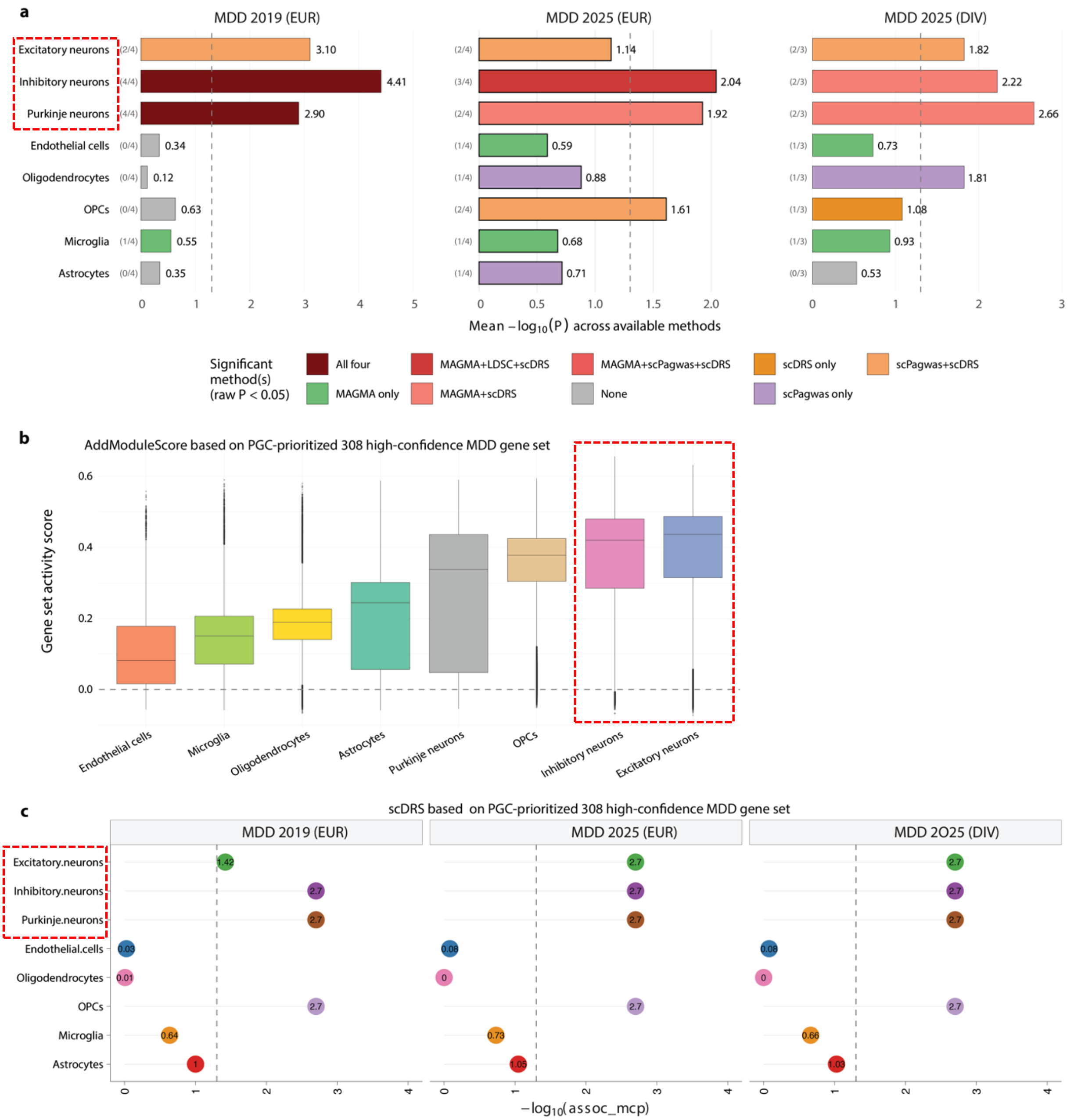


**Supplementary Figure 15 | Cross-method and cross-GWAS analyses support neuronal enrichment of MDD genetic risk, related to Figure 3.**

a. Integrated cell-type enrichment of MDD genetic risk across four complementary approaches—scPagwas, MAGMA-CellTyping, LDSC-SEG and scDRS—applied to three MDD GWAS resources: Howard et al. 2019 European-ancestry GWAS, PGC MDD 2025 European-ancestry GWAS and PGC MDD 2025 trans-ancestry GWAS. LDSC-SEG was not applied to the trans-ancestry GWAS because of ancestry–LD-reference mismatch. For each broad brain cell type, horizontal bars show the mean $-\log_{10}(P)$ across available methods, with values indicated at the bar ends. Bar colours denote the combination of methods reaching nominal significance ($P < 0.05$; colour key). The dashed vertical line indicates $P = 0.05$. Across GWAS datasets and analytical frameworks, neuronal populations showed the strongest enrichment, with recurrent signals in inhibitory, excitatory and Purkinje neurons.

b. Per-cell module activity scores for 308 high-confidence MDD-prioritized genes from the PGC 2025 study across broad cell types in the adult human brain discovery atlas. Gene-set/module scores were calculated using Seurat *AddModuleScore*. Box plots show the median, interquartile range and 1.5 × interquartile range; outliers are omitted for visualization. Cell types are ordered by median module score.

c. scDRS cell-type association analysis using the same 308 PGC-prioritized MDD genes across the three MDD GWAS resources. Each dot represents one cell type within one GWAS dataset. The x axis shows $-\log_{10}$ Monte Carlo association P values; the dashed vertical line indicates $P = 0.05$. Dot colours match the cell-type palette in b. Cell types with dots to the right of the threshold line show nominally significant association.

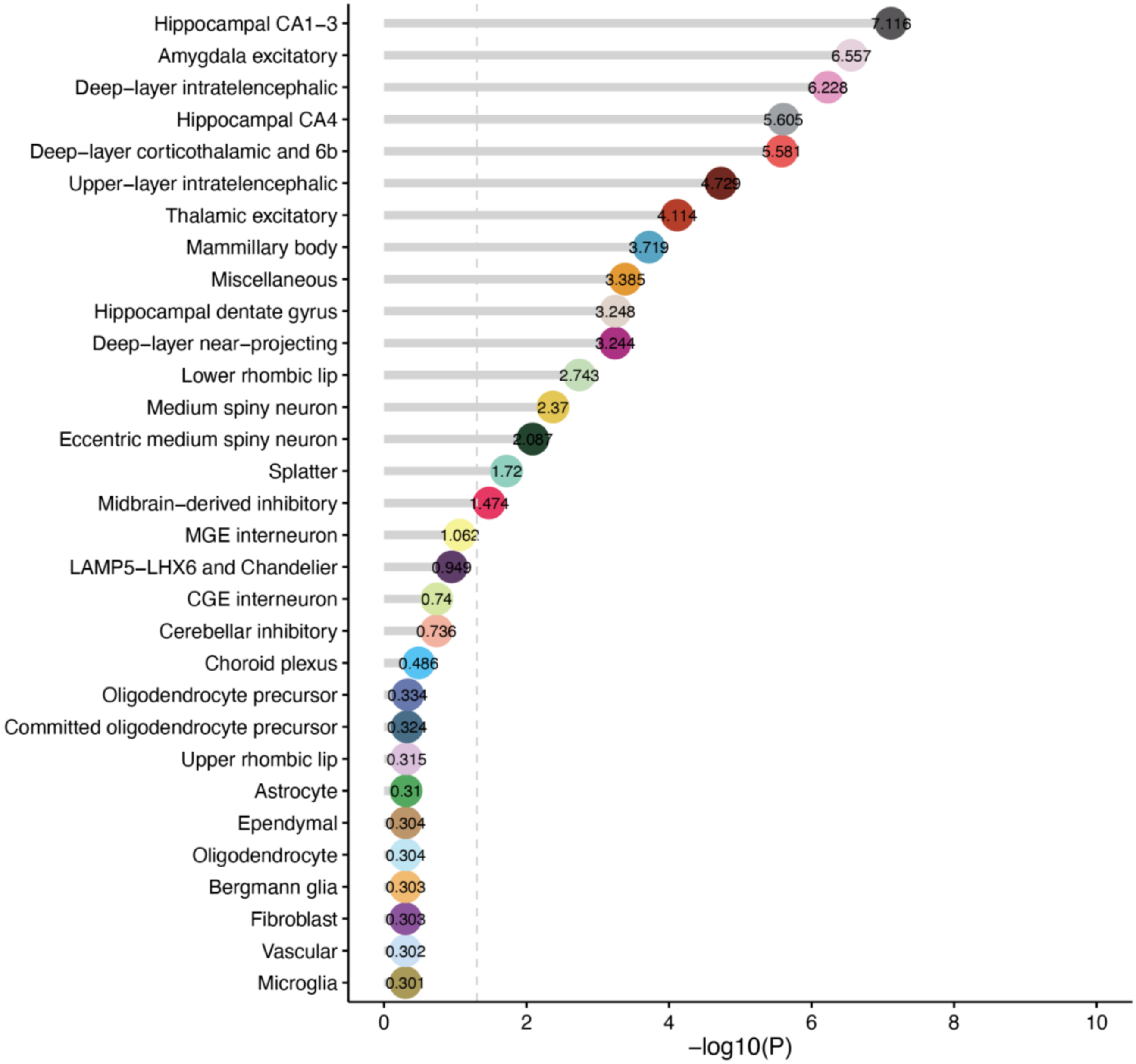


**Supplementary Figure 16 | Bar plot showing the results of scPagwas-based cell type-level inference analysis for MDD in the human adult brain dataset, related to Figure 3.**

This cell type-level inference analysis using scPagwas to infer MDD-relevant cell types among the human adult brain dataset (n = 3,369,219 cells; replication dataset 1). The dash vertical line indicates the significance threshold (FDR < 0.05). The x axis represents negative log2-transformed P-values.

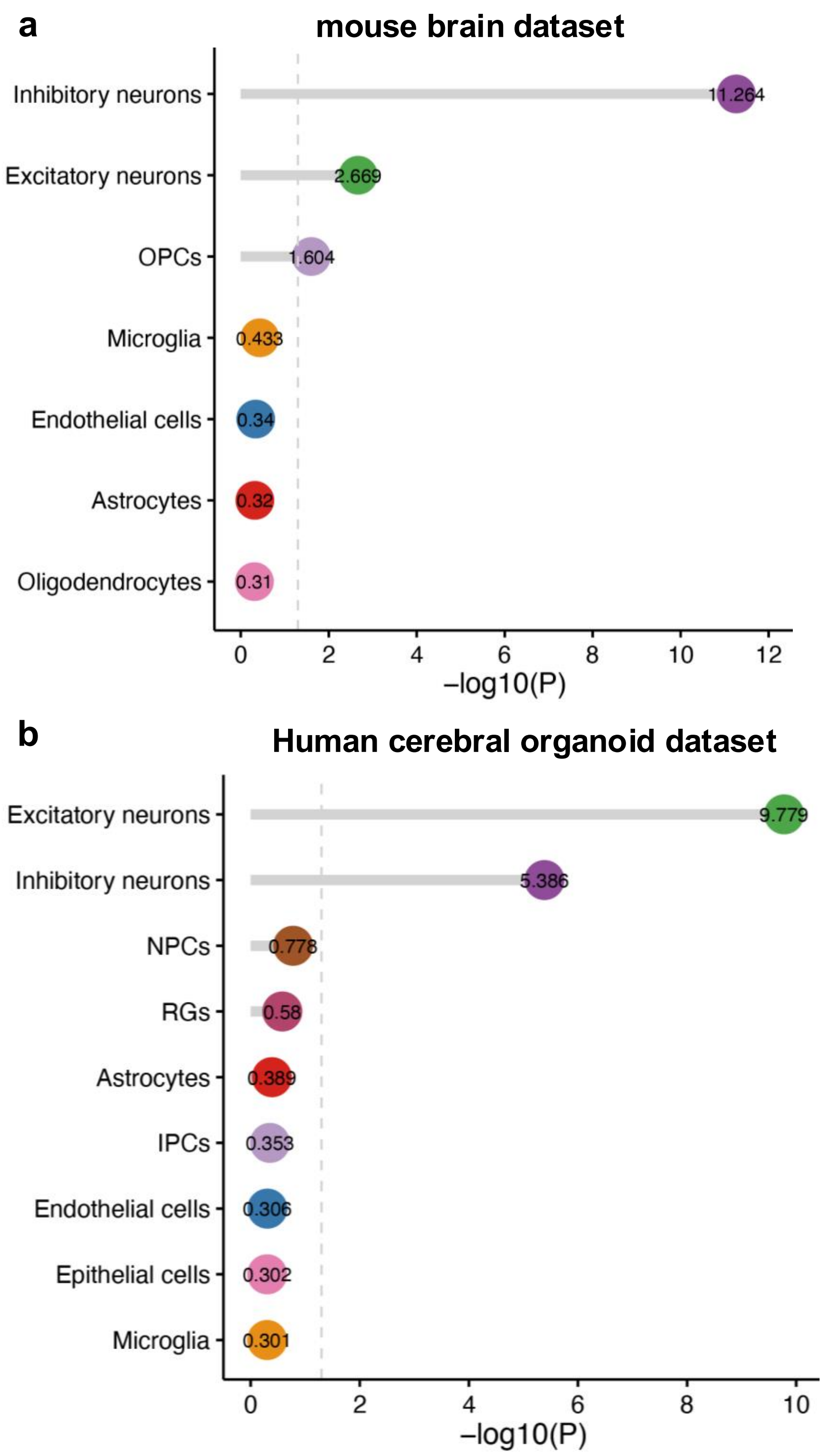


**Supplementary Figure 17 | Dot plots showing the results of cell type-level inference analysis for MDD in the mouse brain and human cerebral organoid using scPagwas, related to Figure 3.**

a. Cell-type-level inference analysis in the mouse brain dataset (n = 197,026 cells; replication dataset 2);

b. Cell type-level inference analysis in the human cerebral organoid dataset (n = 396,049 cells; replication dataset 3).

Note: Cell type-level inference analyses using scPagwas to infer MDD-relevant cell types among the both mouse and human cerebral organoid datasets (replication datasets 3 and 4). The dash vertical line indicates the significance threshold ($P < 0.05$). The x axis represents negative log2-transformed P-values. Box color indicates the cell types.

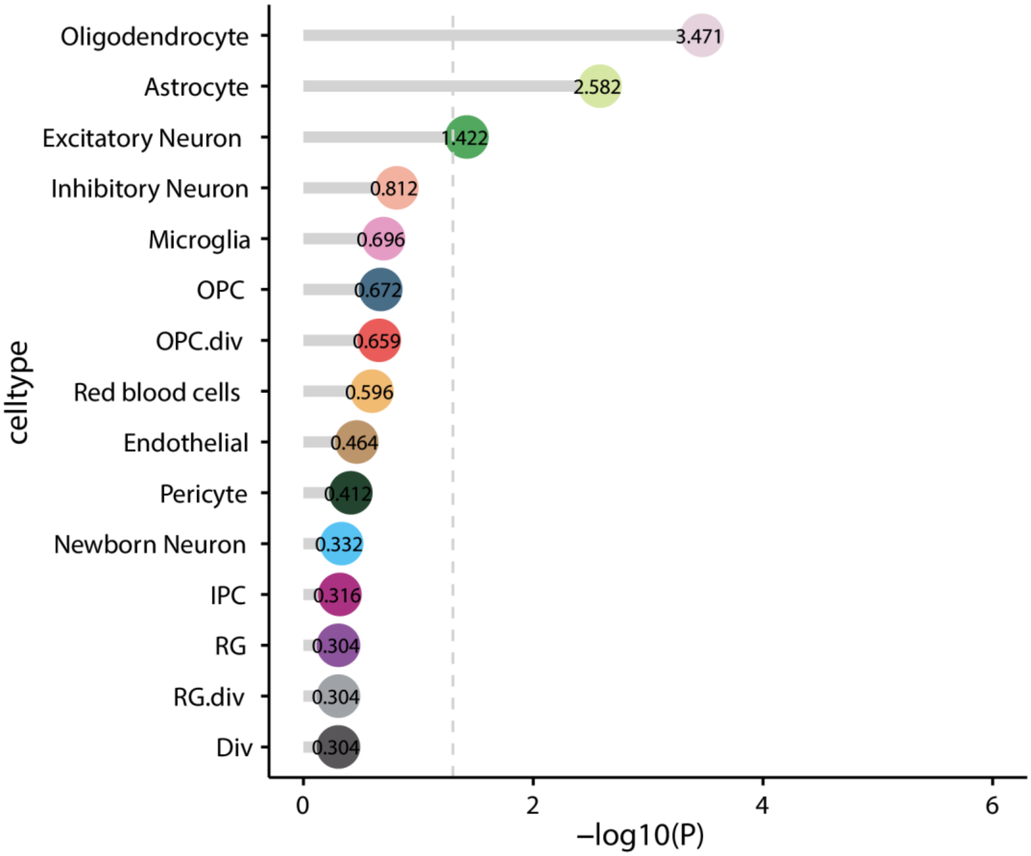


**Supplementary Figure 18 | Bar plot showing the results of scPagwas-based cell type-level inference analysis for MDD in the human developing brain dataset, related to Figure 3.**

This cell type-level inference analysis using scPagwas to infer MDD-relevant cell types among the human developing brain dataset (n = 599,221 cells; replication dataset 4). The dash vertical line indicates the significance threshold (FDR < 0.05). The x axis represents negative log2-transformed P-values.

a

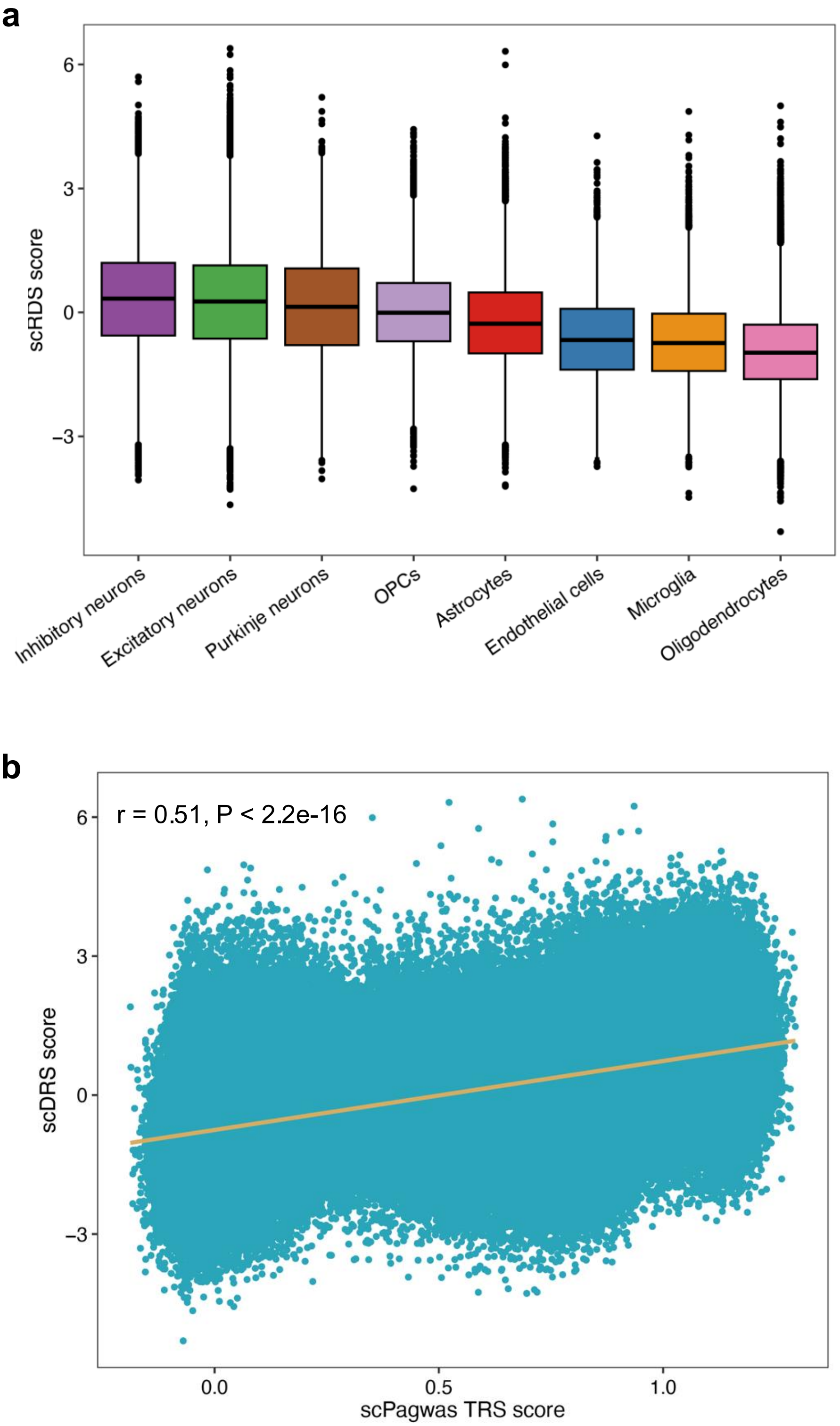


**Supplementary Figure 19 | Results of the single cell-level inference in the discovery dataset using an independent method scDRS, related to Figure 3.**
a. Boxplot showing the single cell-level inference results in the discovery dataset using scDRS. Box plots show median and interquartile range; whiskers indicate 1.5× interquartile range.
b. Correlation analysis of the TRS score of all brain cells between scPagwas and scDRS.
Note: The scDRS TRS score for each cell in corresponding cell type was applied to generate the boxplot. Pearson correlation method was used to calculate the correlation coefficients and statistical significance. Color indicates the cell type.

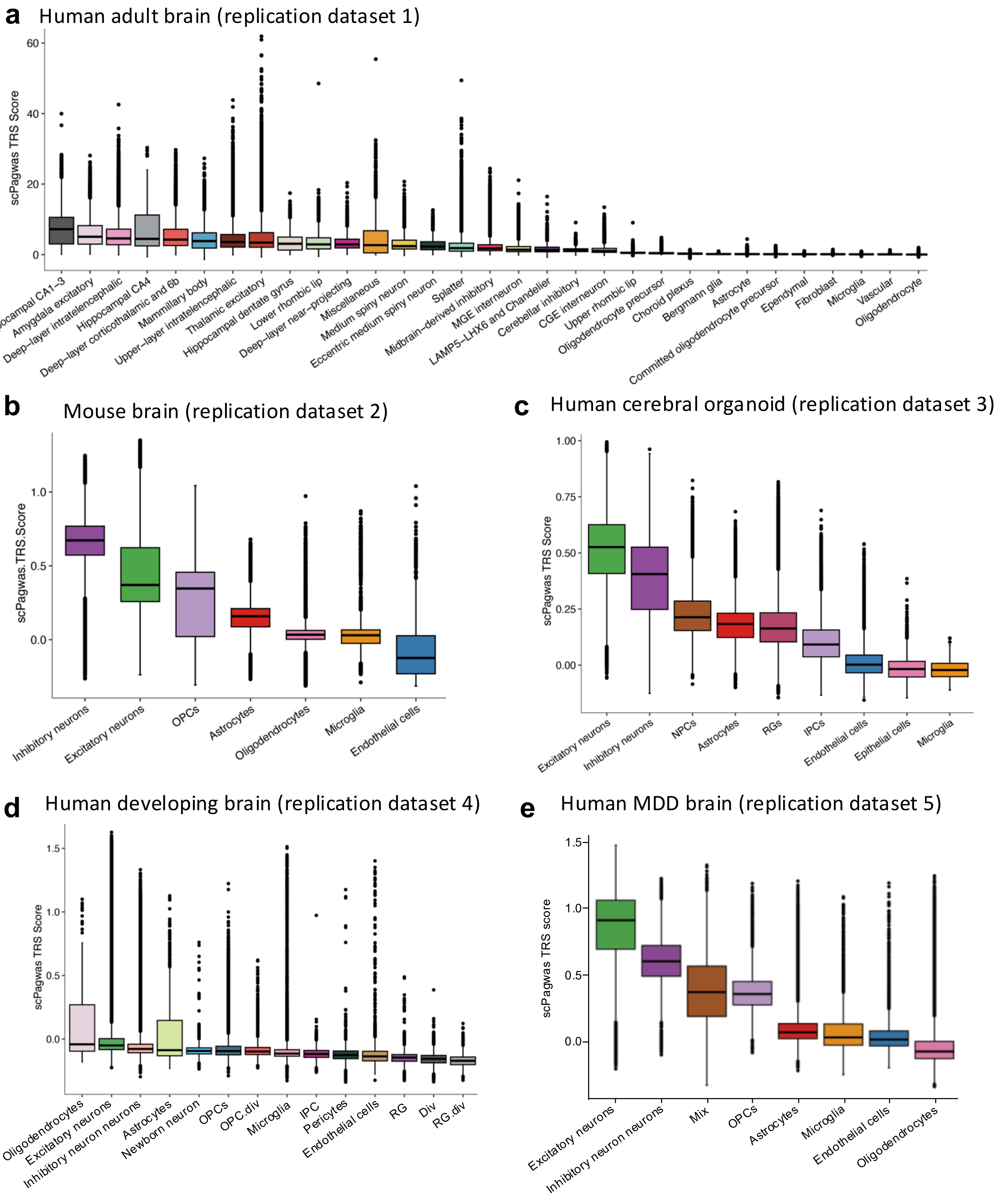


**Supplementary Figure 20 | Single-cell-level scPagwas inference across independent replication datasets, related to Figure 3.**

a–e. Box plots showing cell-type-resolved distributions of scPagwas trait-relevance scores (TRS) across five independent replication datasets: adult human brain (a), mouse brain (b), human cerebral organoids (c), developing human brain (d), and human MDD brain (e). For each dataset, TRS was calculated at single-cell resolution and summarized by annotated cell type. Box plots show median and interquartile range; whiskers indicate 1.5× interquartile range.

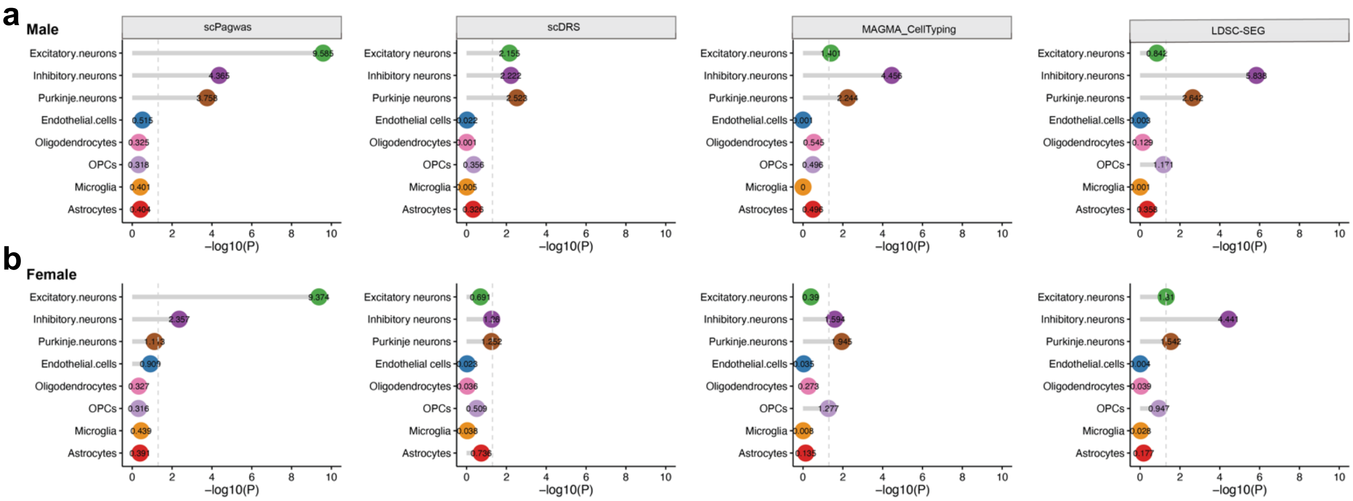


**Supplementary Figure 21 | Sex-stratified cell-type-level inference in the discovery dataset, related to Figure 3.**

a-b. Dot plots showing sex-stratified cell-type-level inference results in male (a) and female (b) samples from the discovery dataset. Analyses were performed using four complementary methods: scPagwas, scDRS, MAGMA_CellTyping, and LDSC-SEG. The dashed vertical line indicates the nominal significance threshold (*P* < 0.05). For more details, please refer to Supplementary Table 12.

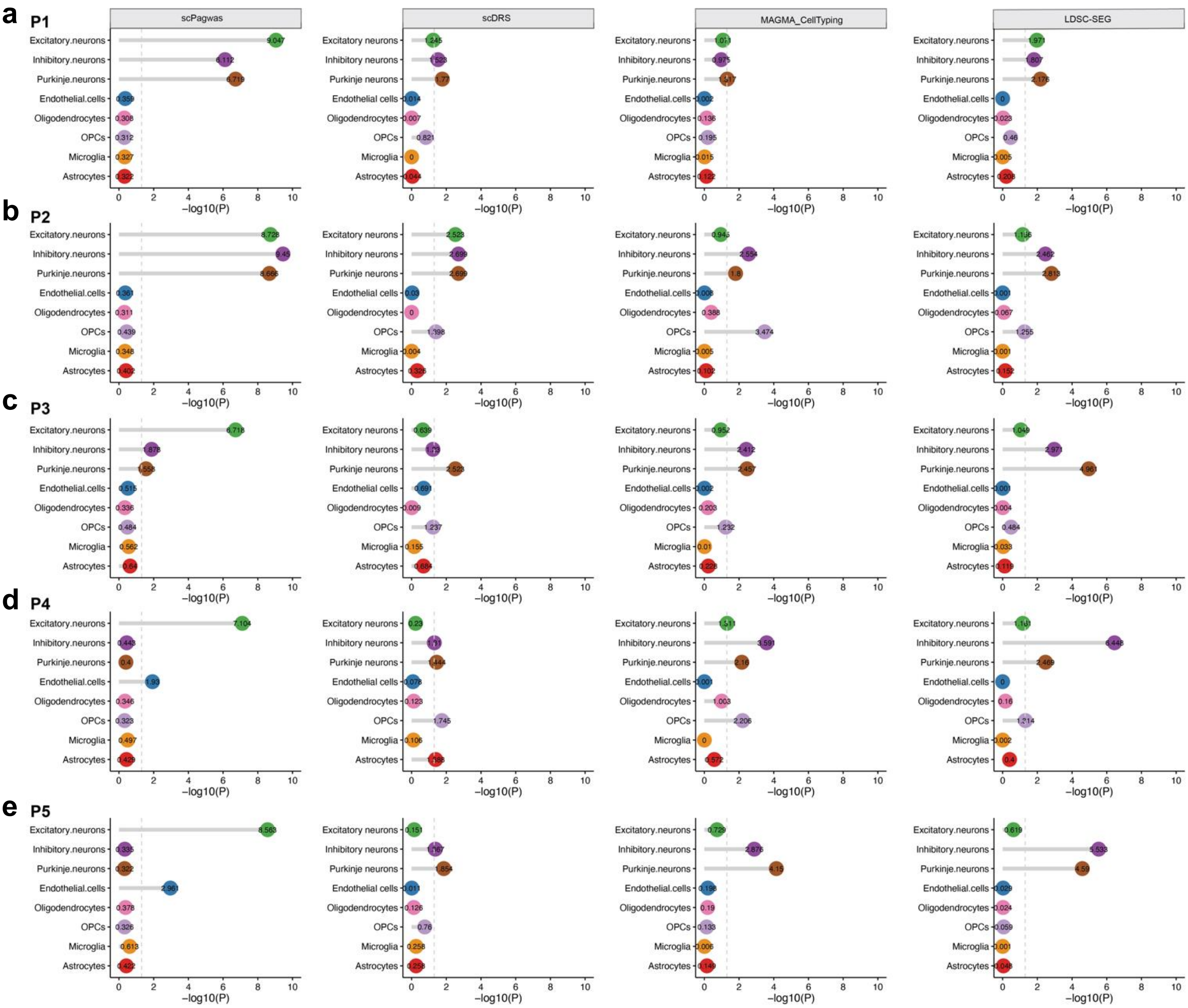


**Supplementary Figure 22 | Age-stratified cell-type-level inference in the discovery dataset, related to Figure 3.**

a–e. Dot plots showing age-stratified cell-type-level inference results across five postnatal periods in the discovery dataset: P1, childhood, 4–8 years (a); P2, adolescence, 12–19 years (b); P3, young adulthood, 20–39 years (c); P4, middle adulthood, 40–59 years (d); and P5, late adulthood, ≥60 years (e). Analyses were performed using four complementary methods: scPagwas, scDRS, MAGMA_CellTyping, and LDSC-SEG. The dashed vertical line indicates the nominal significance threshold (*P* < 0.05). For more details, please refer to Supplementary Table 13.

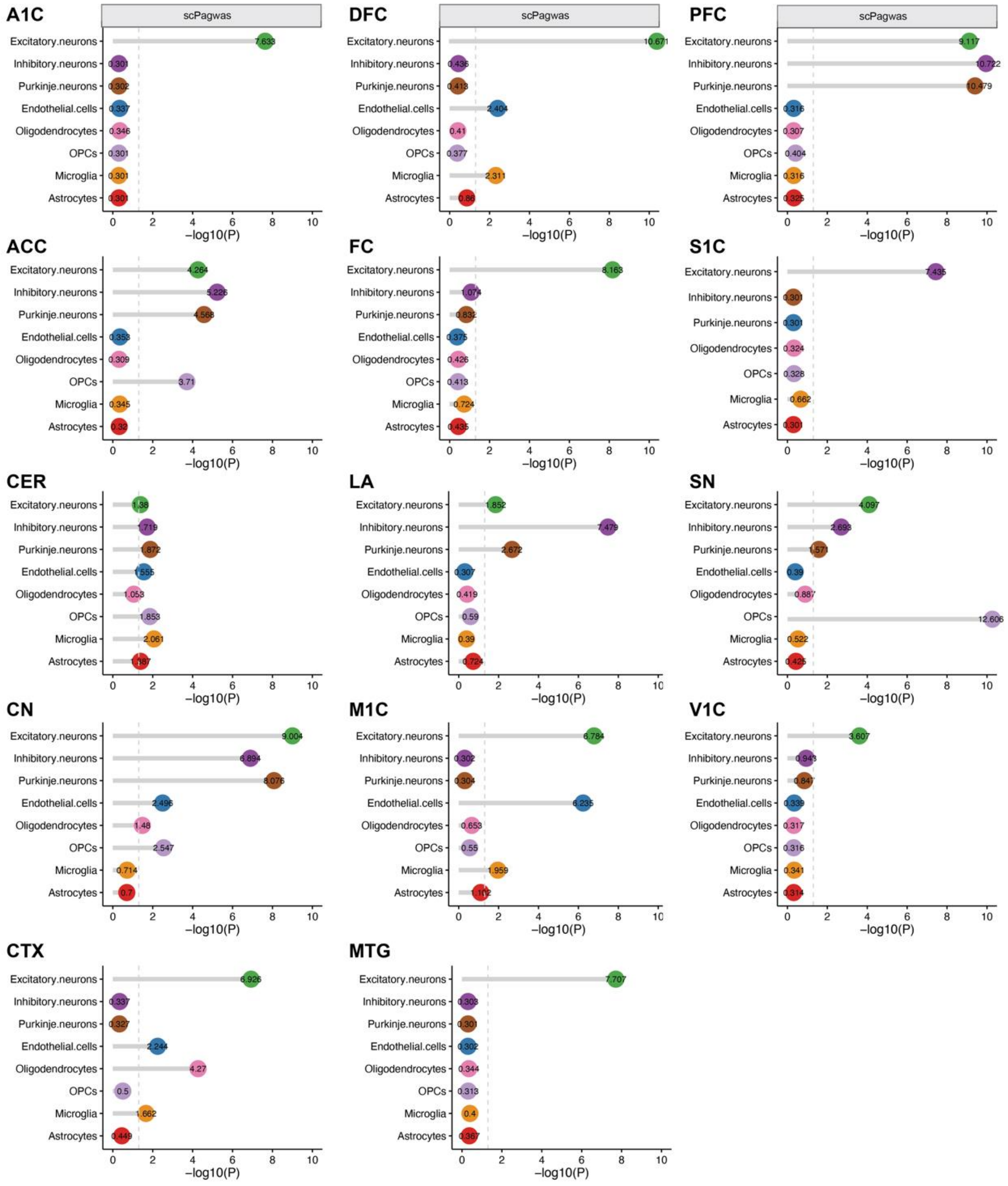


**Supplementary Figure 23 | Dot plots displaying the results of brain region-stratified cell type-level inference analysis in the discovery dataset using scPagwas, related to Figure 3.**

All these cell type-level inference analyses were performed based on the discovery dataset using scPagwas. Dash vertical line indicates the significant threshold (P < 0.05). Abbreviations: ACC - anterior cingulate cortex, CN - caudate nucleus, CTX - cortex, FC - frontal cortex, SN - substantia nigra, CER - cerebellum cortex, S1C - primary somatosensory cortex, M1C - primary motor cortex, DFC - dorsolateral prefrontal cortex, PFC - prefrontal cortex, MTG - middle temporal gyrus, A1C - primary auditory cortex, V1C - primary visual cortex, and LA - lateral. For more details, please refer to Supplementary Table 14.

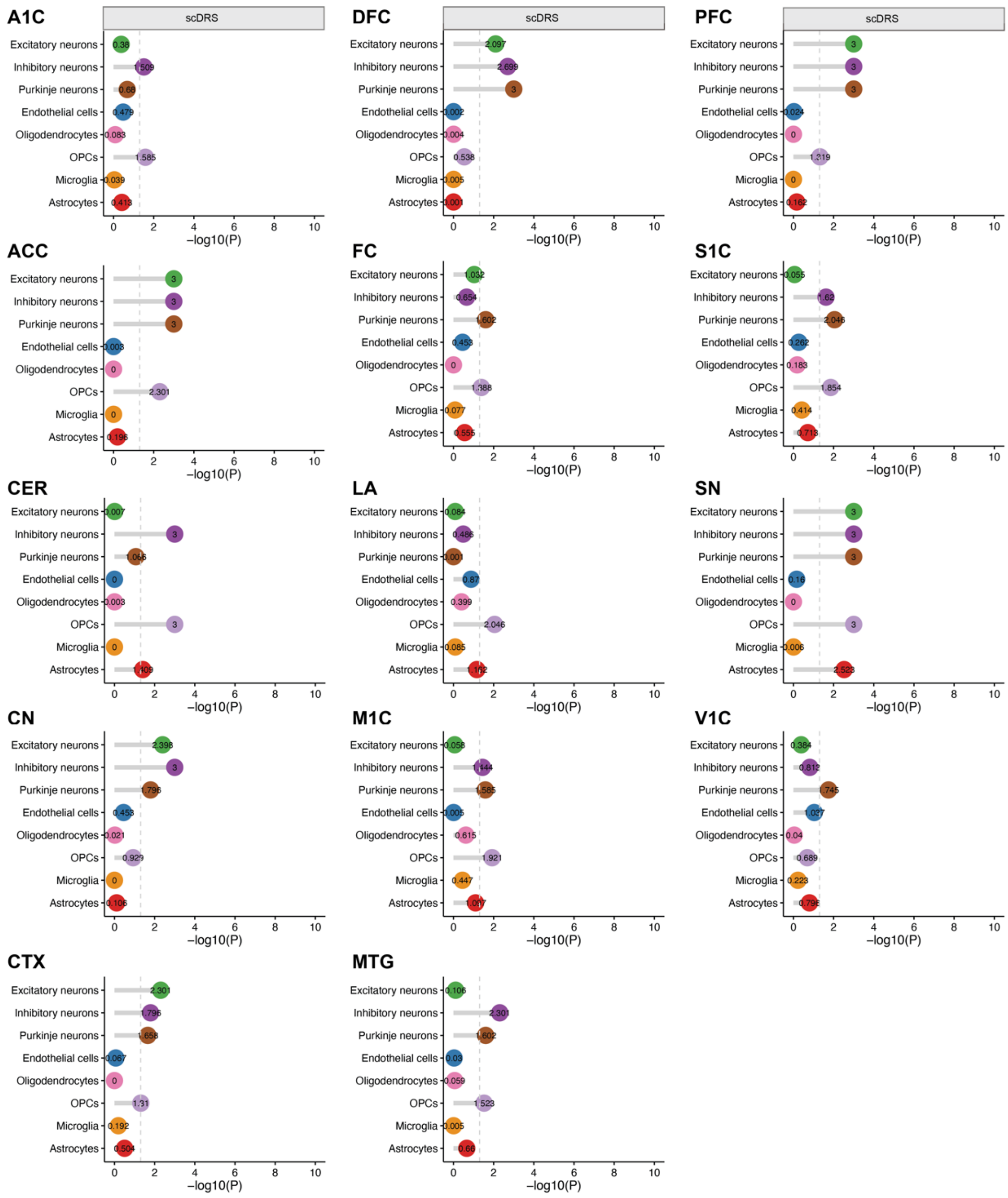

**Supplementary Figure 24 | Dot plots displaying the results of brain region-stratified cell type-level inference analysis in the discovery dataset using scDRS, related to Figure 3.**

All these cell type-level inference analyses were performed based on the discovery dataset using scDRS. Dash vertical line indicates the significant threshold ($P < 0.05$). For more details, please refer to Supplementary Table 15.

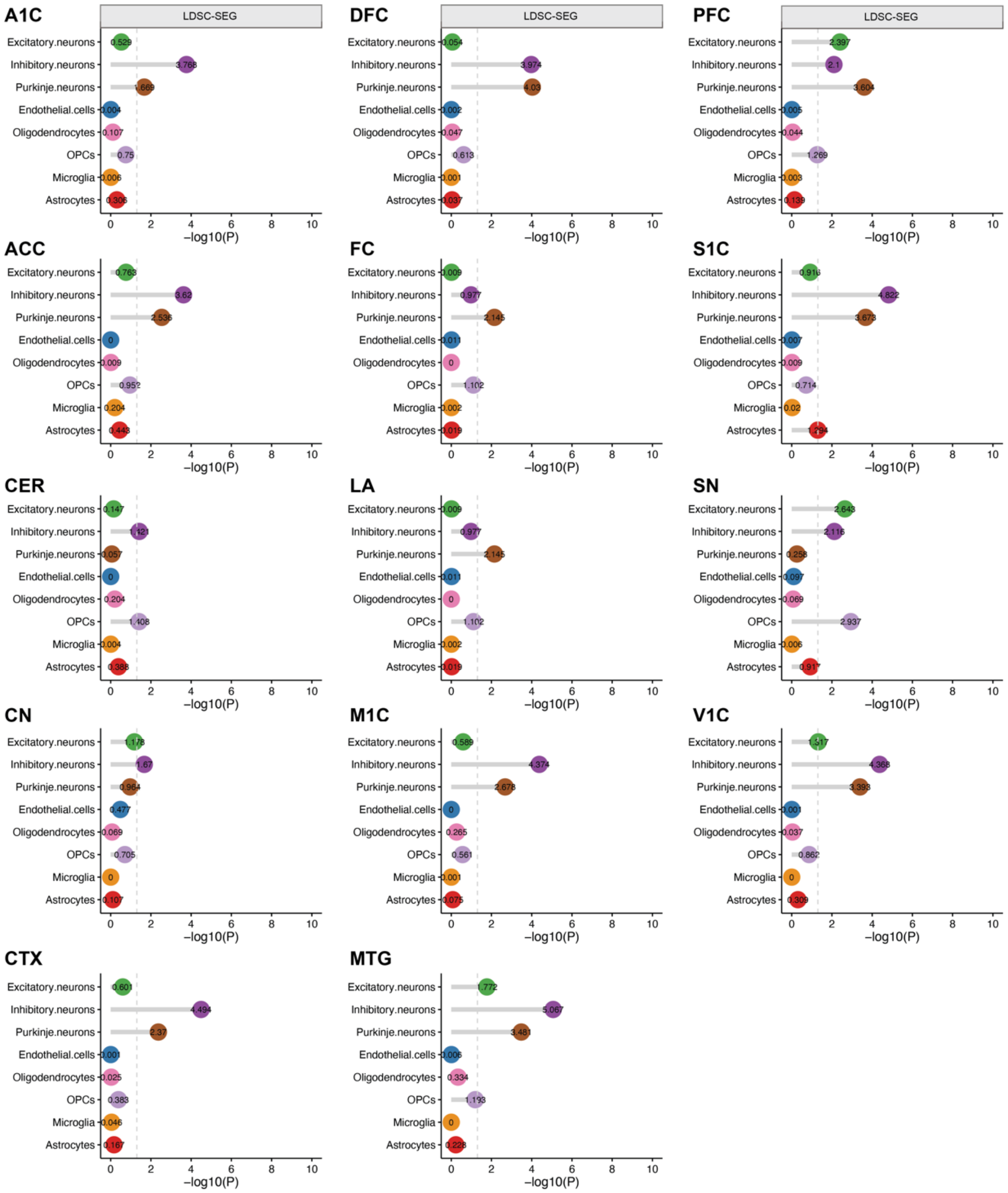


**Supplementary Figure 25 | Dot plots displaying the results of brain region-stratified cell type-level inference analysis in the discovery dataset using LDSC-SEG, related to Figure 3.**

All these cell type-level inference analyses were performed based on the discovery dataset using LDSC-SEG. Dash vertical line indicates the significant threshold (P < 0.05). For more details, please refer to Supplementary Table 16.

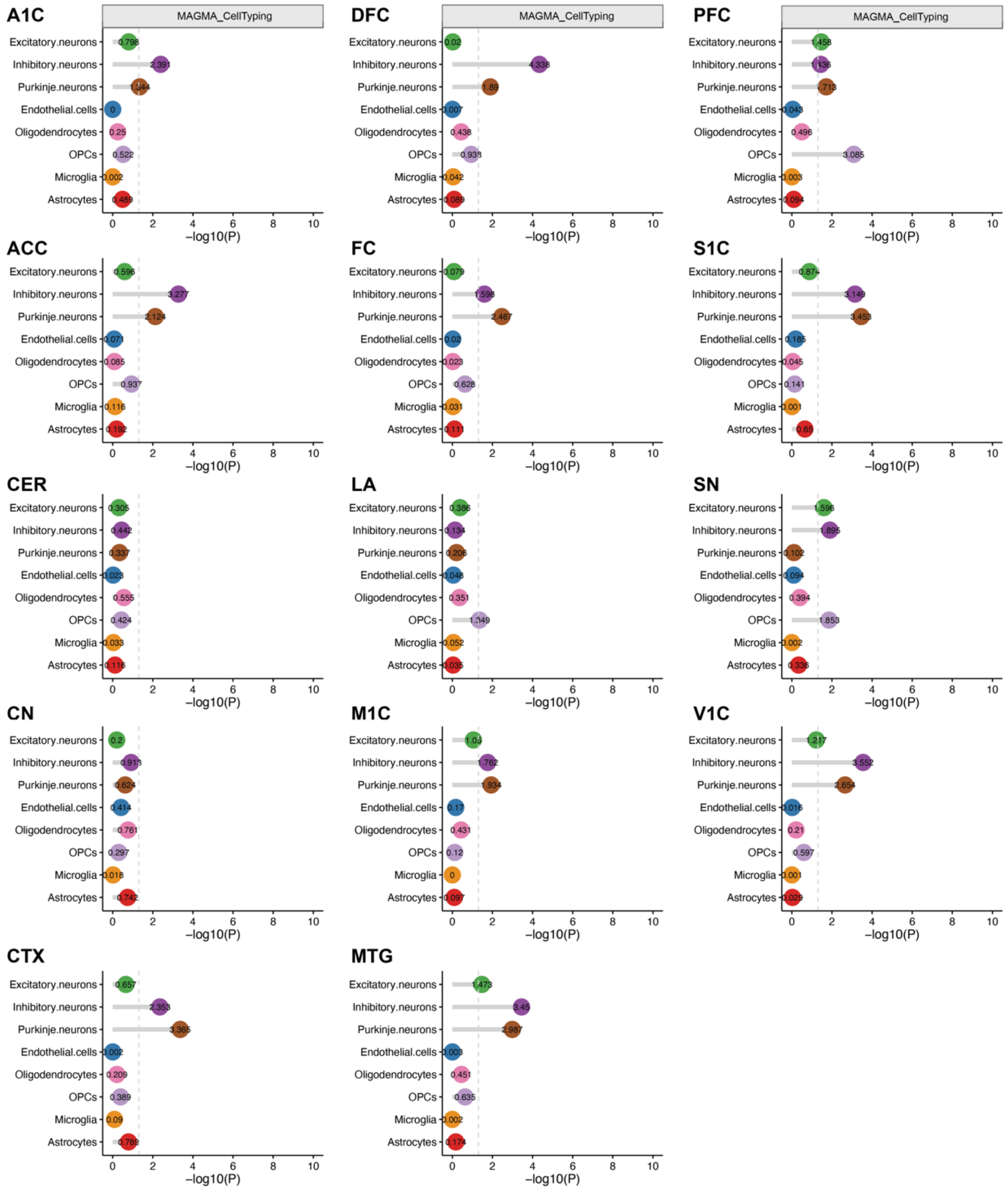


**Supplementary Figure 26 | Dot plots displaying the results of brain region-stratified cell type-level inference analysis in the discovery dataset using MAGMA_CellTyping, related to Figure 3.**

All these cell type-level inference analyses were performed based on the discovery dataset using MAGMA_CellTyping. Dash vertical line indicates the significant threshold ($P < 0.05$). For more details, please refer to Supplementary Table 17.

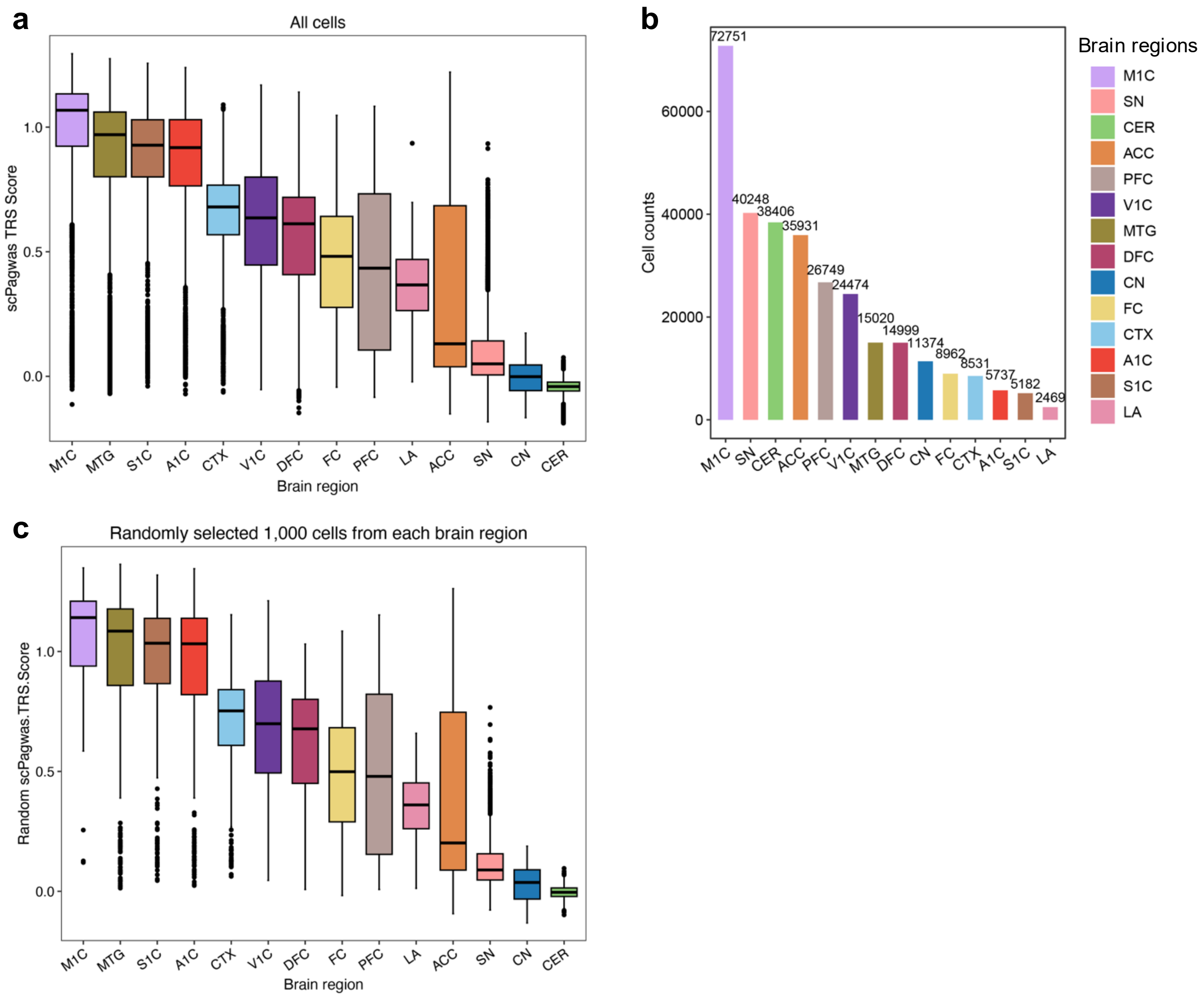


**Supplementary Figure 27 | Brain region-stratified single-cell level inference results in the discovery dataset using scPagwas, related to Figure 3.**

a. Boxplot showing the single cell-level inference results in the discovery dataset based on all cells;

b. Summary of the cell numbers in each brain region across 14 regions in the discovery dataset;

c. Boxplot showing the single cell-level inference results in the discovery dataset based on sub-selection of 1,000 cells per region;

Note: All these single cell level-inference analyses were performed based on the discovery dataset using scPagwas. Dash vertical line indicates the significant threshold ($P < 0.05$). Abbreviations: ACC - anterior cingulate cortex, CN - caudate nucleus, CTX - cortex, FC - frontal cortex, SN - substantia nigra, CER - cerebellum cortex, S1C - primary somatosensory cortex, M1C - primary motor cortex, DFC - dorsolateral prefrontal cortex, PFC - prefrontal cortex, MTG - middle temporal gyrus, A1C - primary auditory cortex, V1C - primary visual cortex, and LA - lateral. Given the varied number of cells in each region, to avoid this bias, we randomly selected 1,000 cells from each brain region to perform single-cell level inference analysis in the discovery dataset using scPagwas. Box plots show median and interquartile range; whiskers indicate 1.5× interquartile range.

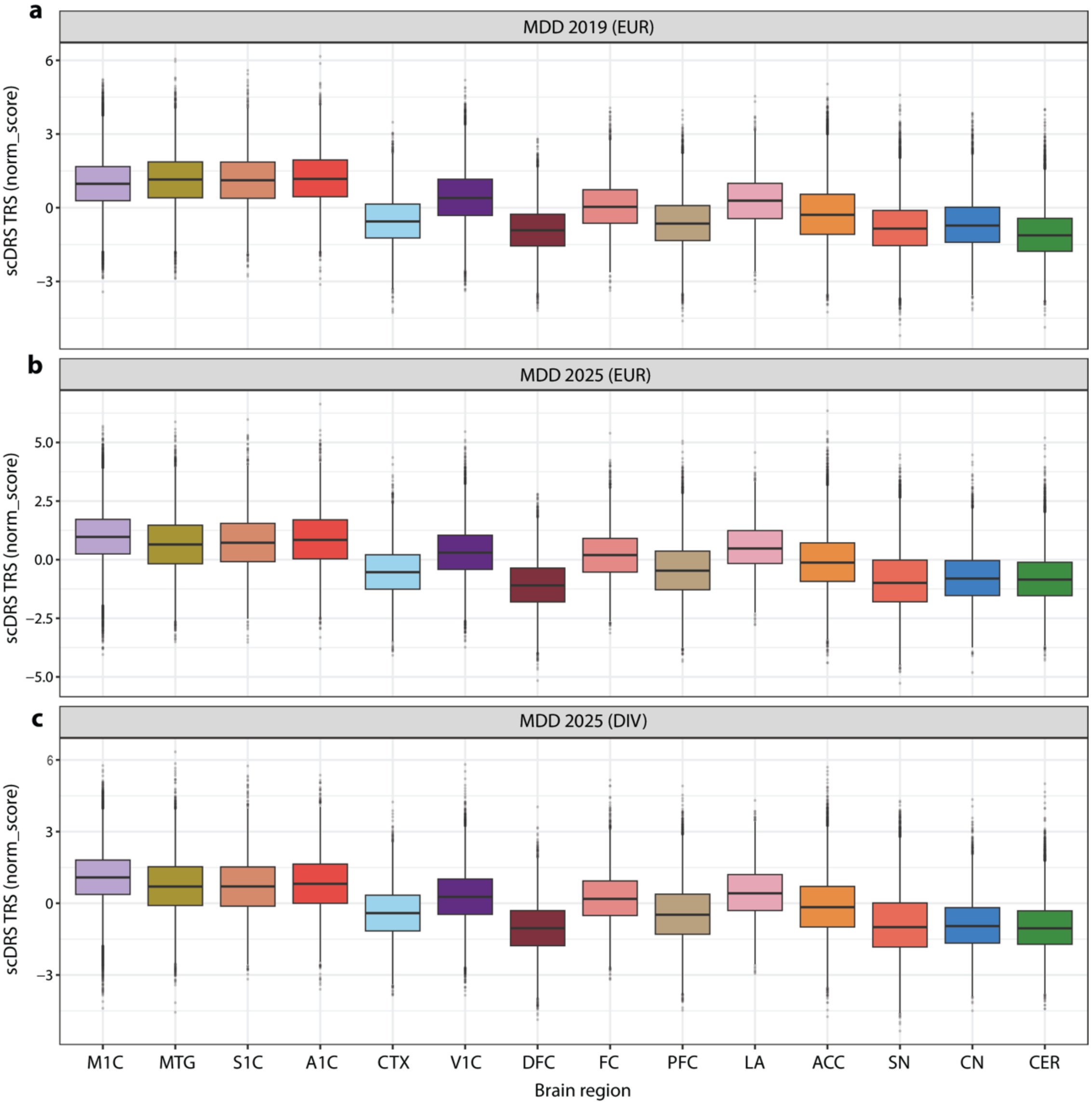


**Supplementary Figure 28 | Regional single-cell MDD trait-relevance scores prioritize cortical regions across GWAS datasets using scDRS, related to Figure 3.**

a–c, Distribution of single-cell MDD trait-relevance scores across 14 anatomical brain regions in the adult human brain discovery atlas, estimated using scDRS with GWAS-specific MAGMA-derived gene weights. Analyses were performed using three MDD GWAS resources: a, Howard et al. 2019 European-ancestry GWAS (MDD 2019 EUR); b, PGC 2025 MDD European-ancestry GWAS (MDD 2025 EUR); and c, PGC MDD 2025 trans-ancestry GWAS (MDD 2025 DIV). Box plots show the median, interquartile range and 1.5 × interquartile range; individual outlier cells are shown as black points. The dashed horizontal line indicates a normalized score of zero. Across all three GWAS datasets, cortical regions showed higher MDD trait-relevance scores than subcortical and cerebellar regions, with cortical regions (including M1C, A1C, MTG and S1C) consistently among the highest-scoring regions across all three GWAS resources. These results support the robustness of the cortical and motor-cortical enrichment pattern across GWAS resources.

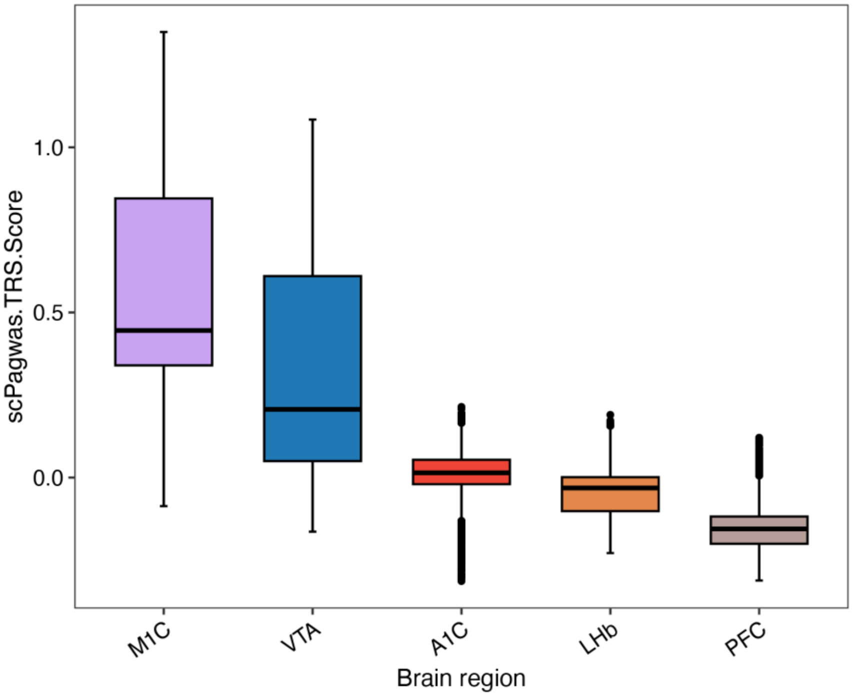


**Supplementary Figure 29 | M1C shows elevated MDD trait-relevance scores among selected depression-relevant brain regions in an independent mouse brain atlas, related to Figure 3.**

Box plots showing single-cell scPagwas trait-relevance scores (TRSs) in the integrated mouse brain single-cell replication dataset (replication dataset 2, n = 197,026 cells) across five selected brain regions: primary motor cortex (M1C), ventral tegmental area (VTA), primary auditory cortex (A1C), lateral habenula (LHb) and prefrontal cortex (PFC). These regions were selected to represent motor-cortical, reward-related, sensory, habenular and prefrontal contexts relevant to depression-related biology. Consistent with the brain-wide regional analysis and cell-number-matched subsampling results, M1C showed the highest TRS distribution among these selected regions in the independent mouse atlas. Box plots show median and interquartile range; whiskers indicate 1.5× interquartile range.

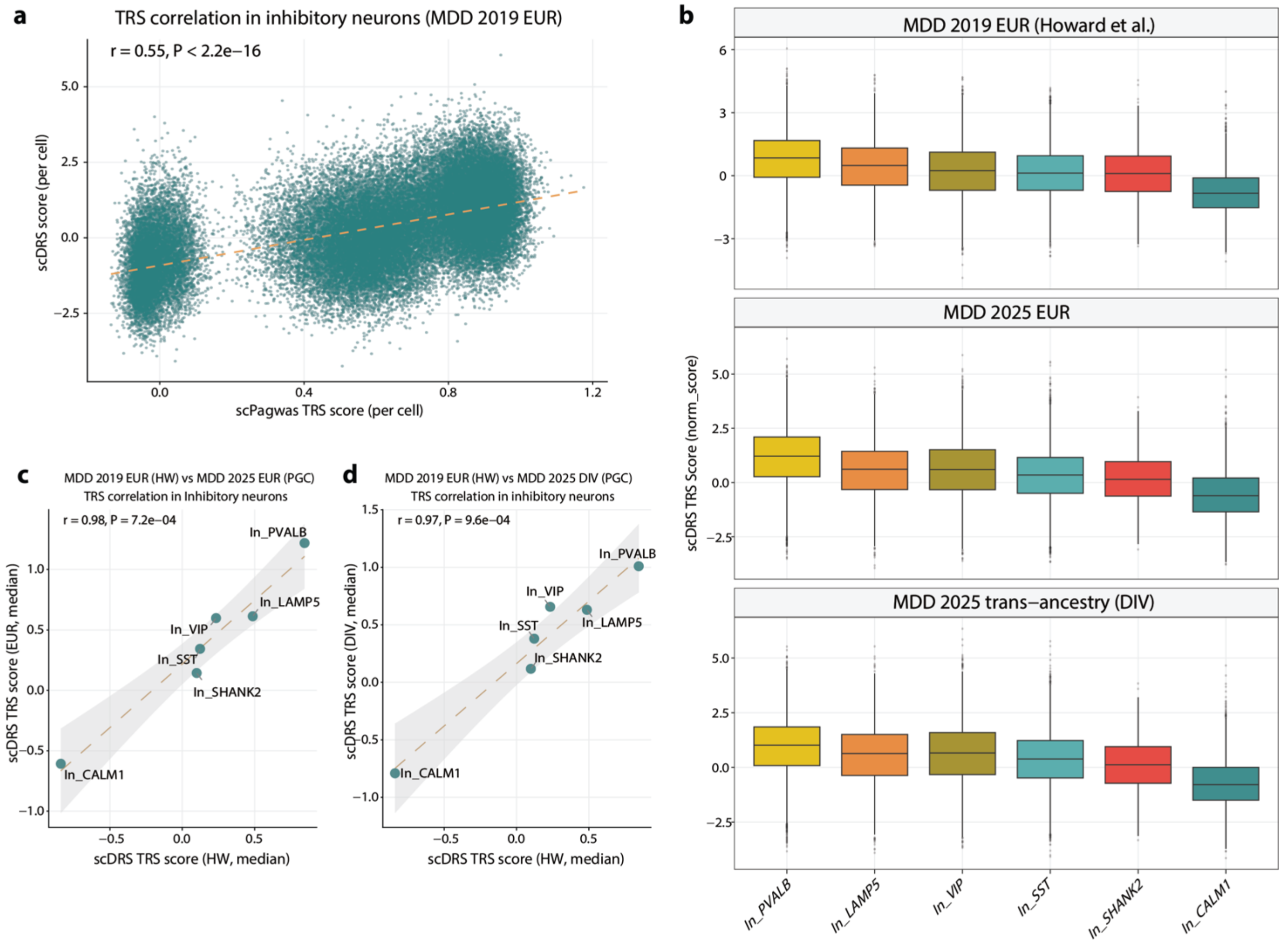


**Supplementary Figure 30 | scDRS analysis supports reproducible PVALB⁺ inhibitory-neuron MDD signals across GWAS datasets, related to Figure 4.**

a. Single-cell-level correlation between scPagwas trait-relevance scores (TRSs) and scDRS normalized scores across inhibitory neurons in the adult human brain discovery atlas, using the Howard et al. 2019 European-ancestry MDD GWAS (EUR). Each point represents one cell. The dashed line indicates the linear regression fit. scPagwas and scDRS scores showed significant single-cell-level concordance.

b. Distribution of per-cell scDRS normalized scores across inhibitory neuron subtypes for three MDD GWAS resources: Howard et al. 2019 European-ancestry GWAS (EUR), PGC MDD 2025 European-ancestry GWAS (EUR) and PGC MDD 2025 trans-ancestry GWAS (DIV). Box plots show the median, interquartile range and 1.5 × interquartile range; individual outlier cells are shown as points. $PVALB^{+}$ inhibitory neurons consistently showed the highest median scDRS scores across all three GWAS datasets.

c. Subtype-level correlation of median scDRS scores between the Howard et al. 2019 GWAS and the PGC MDD 2025 European-ancestry GWAS across inhibitory neuron subtypes. Each point represents one subtype; the dashed line indicates the linear regression fit and shading indicates the 95% confidence interval. Subtype rankings were highly concordant between the two European-ancestry GWAS datasets.

d. Subtype-level correlation of median scDRS scores between the Howard et al. 2019 GWAS and the PGC MDD 2025 trans-ancestry GWAS across inhibitory neuron subtypes. Plot conventions are as in c. The strong concordance supports the robustness of the $PVALB^{+}$ inhibitory-neuron signal across GWAS resources and ancestry composition.

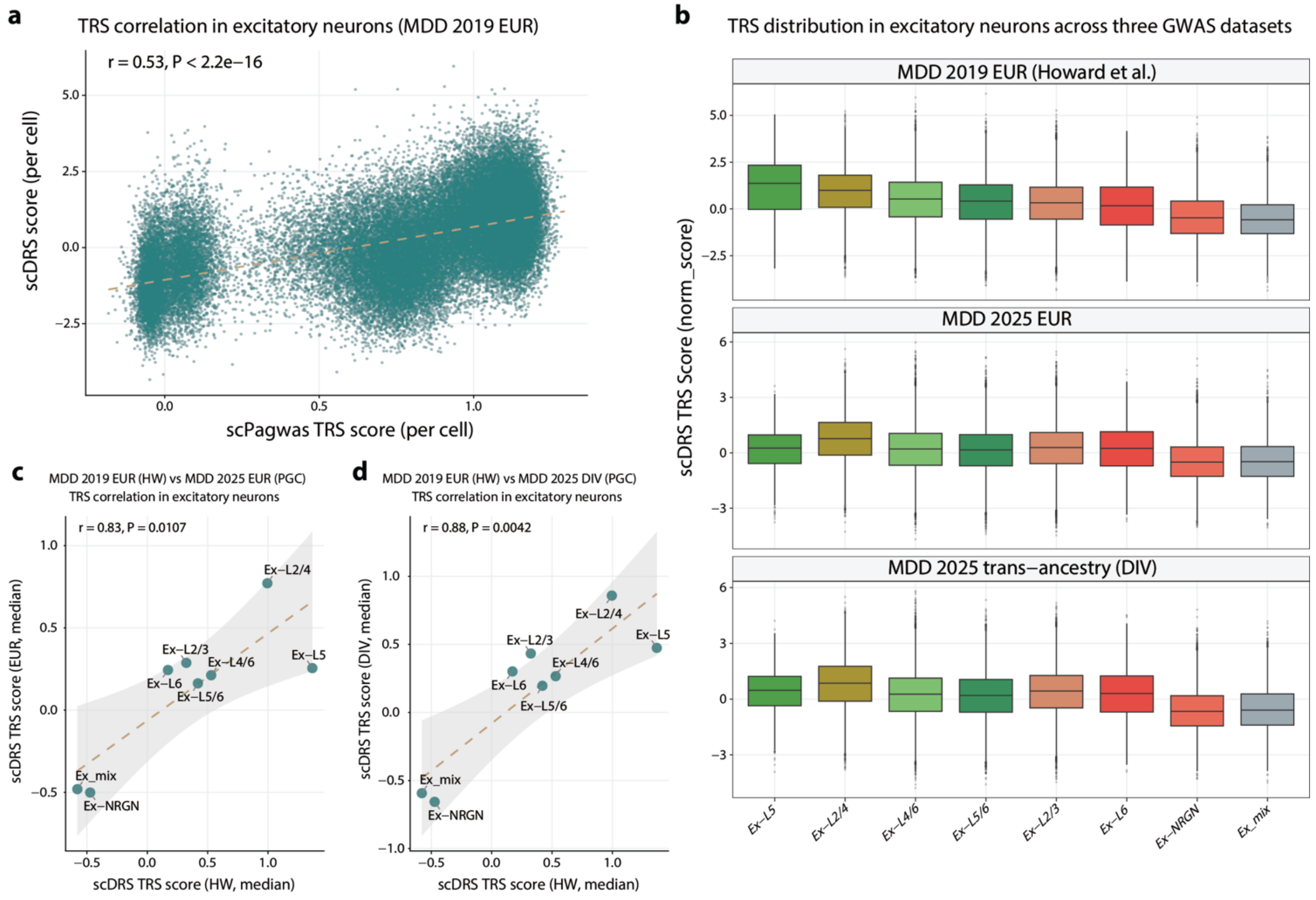


**Supplementary Figure 31 | scDRS analysis supports reproducible Ex-L2/4 excitatory-neuron signals across GWAS datasets, related to Figure 4.**

a. Single-cell-level correlation between scPagwas trait-relevance scores (TRSs) and scDRS normalized scores across excitatory neurons in the adult human brain discovery atlas, using the Howard et al. 2019 European-ancestry MDD GWAS. Each point represents one cell. The dashed line indicates the linear regression fit. scPagwas and scDRS scores showed significant single-cell-level concordance.

b. Distribution of per-cell scDRS normalized scores across excitatory neuron subtypes for three MDD GWAS resources: Howard et al. 2019 European-ancestry GWAS, PGC MDD 2025 European-ancestry GWAS and PGC MDD 2025 trans-ancestry GWAS. Box plots show the median, interquartile range and 1.5 × interquartile range; individual outlier cells are shown as points. Ex-L2/4 excitatory neurons consistently ranked among the highest-scoring excitatory subtypes across all three GWAS datasets.

c. Subtype-level correlation of median scDRS scores between the Howard et al. 2019 GWAS and the PGC MDD 2025 European-ancestry GWAS across excitatory neuron subtypes. Each point represents one subtype; the dashed line indicates the linear regression fit and shading indicates the 95% confidence interval. Subtype rankings were highly concordant between the two European-ancestry GWAS datasets.

d. Subtype-level correlation of median scDRS scores between the Howard et al. 2019 GWAS and the PGC MDD 2025 trans-ancestry GWAS across excitatory neuron subtypes. Plot conventions are as in **c**. The strong concordance supports the robustness of excitatory-neuron subtype prioritization across GWAS resources and ancestry composition, with Ex-L2/4 showing a reproducible high-ranking signal.

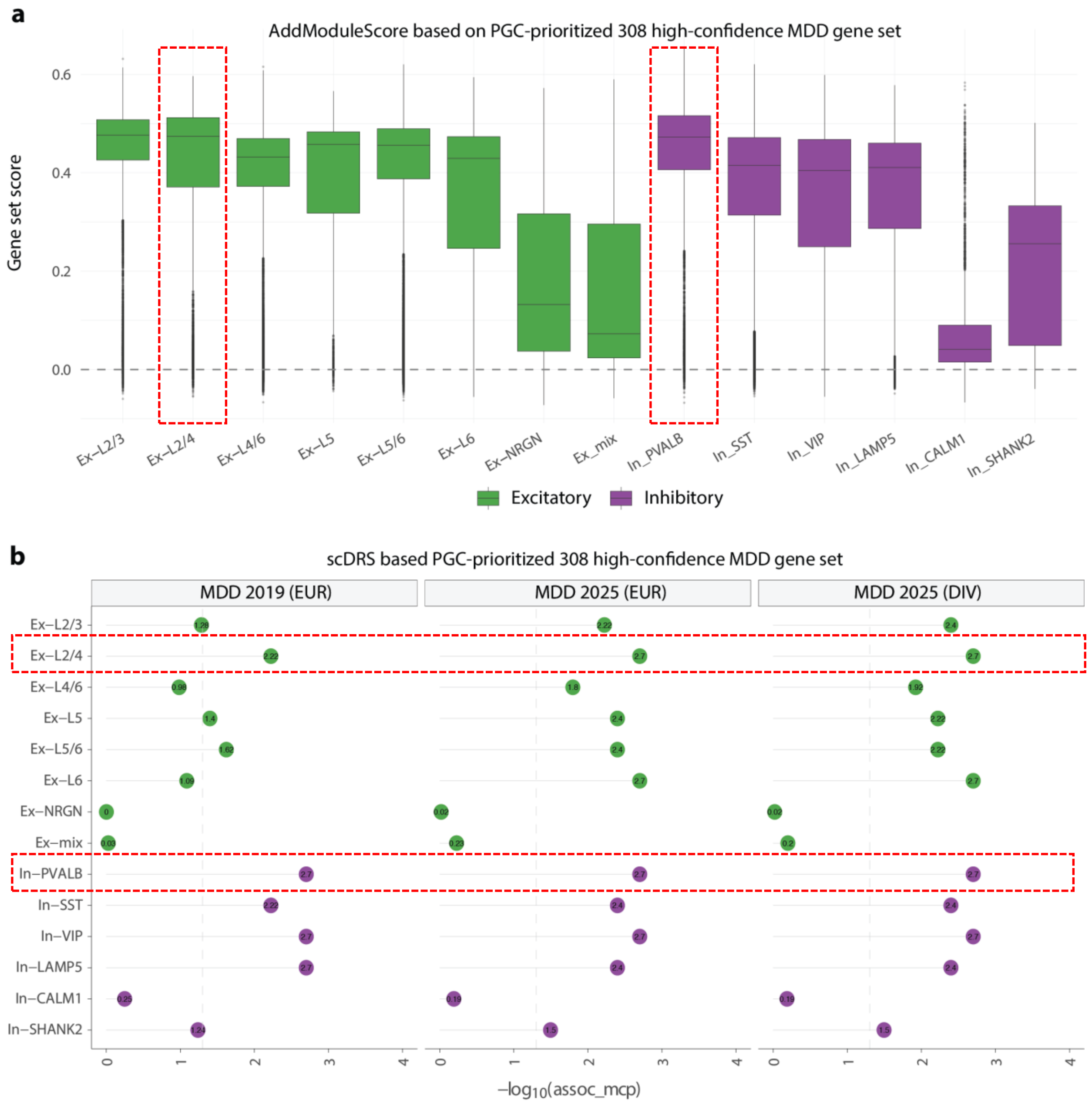


**Supplementary Figure 32 | PGC-prioritized MDD genes converge on Ex-L2/4 excitatory neurons and PVALB$^+$ inhibitory neurons, related to Figure 4.**

a. Per-cell module activity scores for the 308 high-confidence MDD genes prioritized by the latest PGC study across cortical excitatory and inhibitory neuronal subtypes in the adult human brain discovery atlas. Gene-set/module scores were calculated using Seurat *AddModuleScore*. Box plots show the median, interquartile range and 1.5 × interquartile range; individual outlier cells are shown as points. Colours denote neuronal class: excitatory neurons in green and inhibitory neurons in purple. Red dashed boxes highlight Ex-L2/4 excitatory neurons and PVALB$^+$ inhibitory neurons, which showed the highest module activity within their respective neuronal classes.

b. scDRS subtype-level association analysis using the same 308-gene set across three MDD GWAS resources: Howard et al. 2019 European-ancestry GWAS, PGC MDD 2025 European-ancestry GWAS and PGC MDD 2025 trans-ancestry GWAS. Gene sets were weighted by GWAS-specific MAGMA gene-level statistics. Each dot represents one neuronal subtype within one GWAS dataset. The x axis shows $-\log_{10}$ Monte Carlo association P values; the dashed vertical line indicates P = 0.05. Dot colours match the neuronal classes in a. Red dashed boxes indicate Ex-L2/4 excitatory neurons and PVALB$^+$ inhibitory neurons, both of which showed recurrent subtype-level associations across GWAS datasets, consistent with their elevated module activity in a.

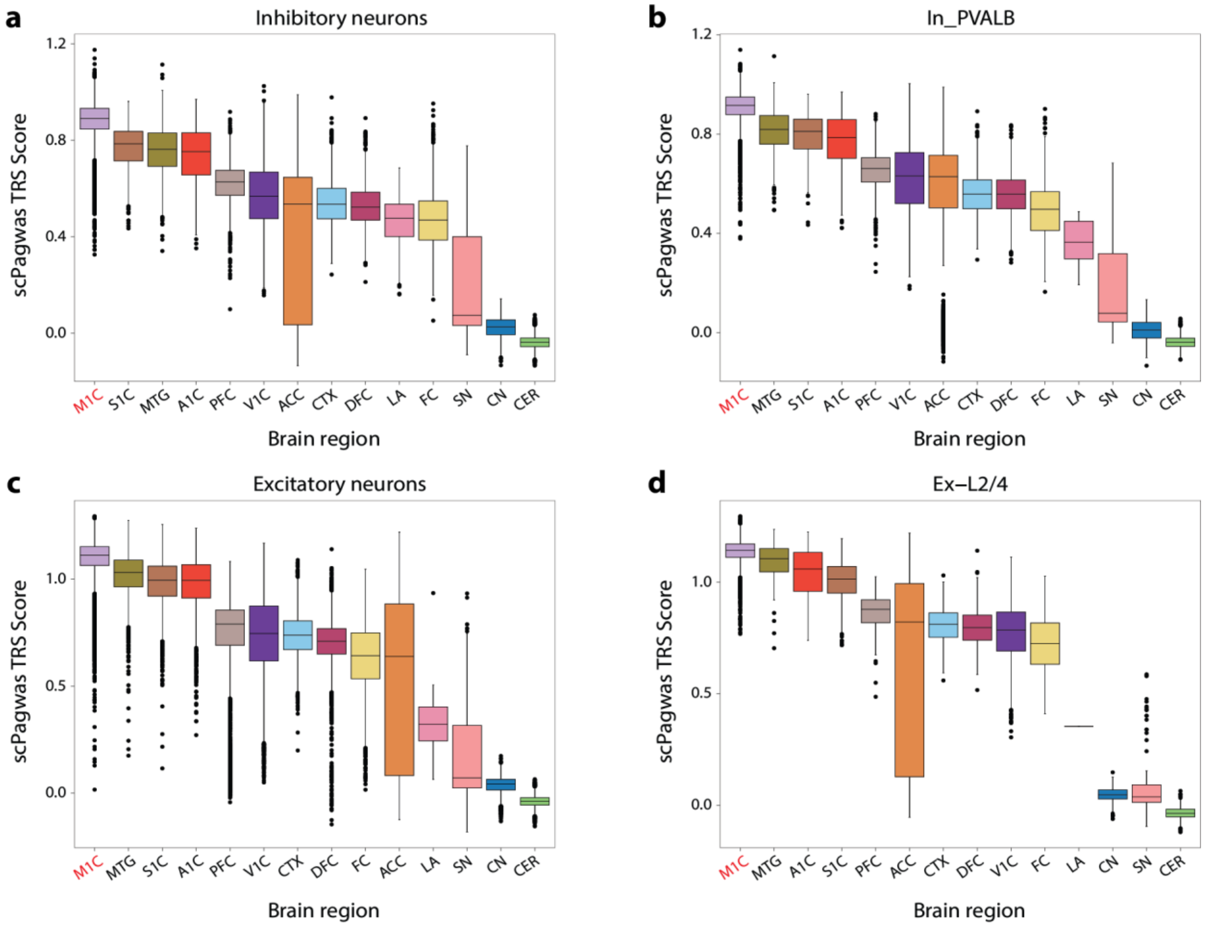


**Supplementary Figure 33 | Regional distribution of MDD trait-relevance scores across neuronal populations and prioritized neuronal subtypes, related to Figure 4.**

Box plots showing single-cell scPagwas trait-relevance scores (TRSs) using Howard et al. 2019 European-ancestry GWAS across brain regions in a, inhibitory neurons; b, $PVALB^{+}$ inhibitory neurons; c, excitatory neurons; and d, Ex-L2/4 excitatory neurons. Across both broad neuronal classes and prioritized neuronal subtypes, M1C showed consistently elevated TRS distributions relative to other brain regions, supporting its prioritization as a motor-cortical anatomical context for MDD-relevant neuronal risk. Box plots show median and interquartile range; whiskers indicate 1.5× interquartile range.

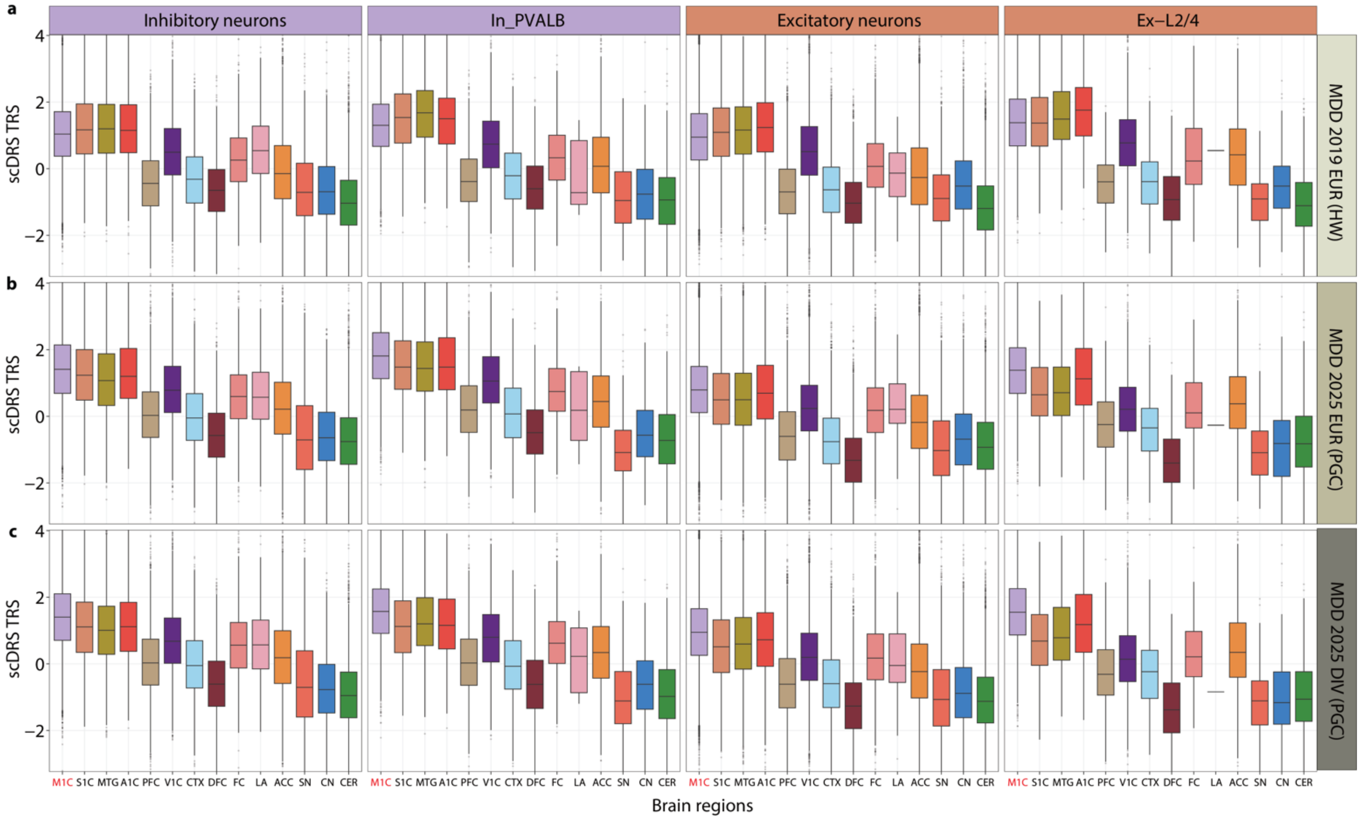


**Supplementary Figure 34 | Regional MDD trait-relevance patterns are preserved within prioritized neuronal populations across GWAS datasets, related to Figure 4.**

Per-cell scDRS normalized scores were stratified by brain region across four neuronal populations in the adult human brain discovery atlas: all inhibitory neurons, PVALB$^{+}$ inhibitory neurons, all excitatory neurons and Ex-L2/4 excitatory neurons. Rows show analyses based on three MDD GWAS resources: a, Howard et al. 2019 European-ancestry GWAS (MDD 2019 EUR); b, PGC MDD European-ancestry GWAS (MDD 2025 EUR); and c, PGC MDD trans-ancestry GWAS (MDD 2025 DIV). Box plots show the median, interquartile range and 1.5 × interquartile range; individual outlier cells are shown as points. Brain regions are ordered consistently across panels, and primary motor cortex (M1C) is highlighted in red. Across neuronal classes and prioritized neuronal subtypes, cortical regions showed higher MDD trait-relevance scores than subcortical and cerebellar regions, with M1C consistently among the highest-scoring regions across all three GWAS datasets. These results support the regional scPagwas findings and indicate that the M1C-prioritized pattern is preserved within the neuronal populations nominated by subtype-level genetic analyses, including PVALB$^{+}$ inhibitory neurons and Ex-L2/4 excitatory neurons.

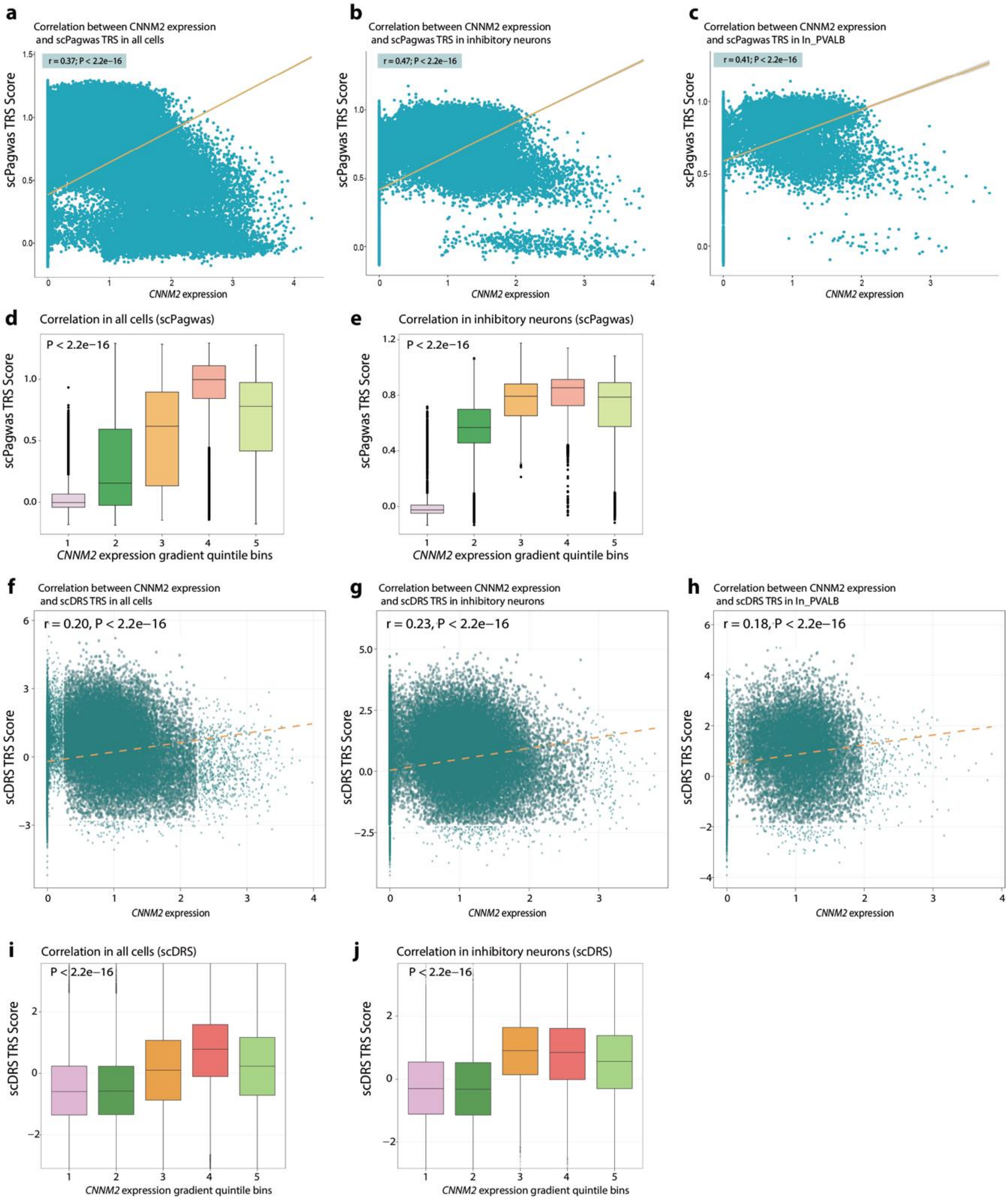


**Supplementary Figure 35 | *CNNM2* expression is associated with single-cell MDD genetic relevance scores in PVALB$^+$ inhibitory neurons, related to Figure 5.**

a–c, Scatter plots showing the relationship between *CNNM2* expression and scPagwas trait-relevance scores (TRSs) across all cells (a), inhibitory neurons (b) and PVALB$^+$ inhibitory neurons (c). Each point represents one cell; Pearson correlation coefficients and P values are shown.
d,e, Distribution of scPagwas TRSs across quintiles of *CNNM2* expression in all cells (d) and inhibitory neurons (e).
f–h, Scatter plots showing the relationship between *CNNM2* expression and scDRS disease-relevance scores across all cells (f), inhibitory neurons (g) and PVALB$^+$ inhibitory neurons (h). Pearson correlation coefficients and P values are shown.
i,j, Distribution of scDRS disease-relevance scores across quintiles of *CNNM2* expression in all cells (i) and inhibitory neurons (j). Analyses were based on the primary Howard et al. MDD GWAS.

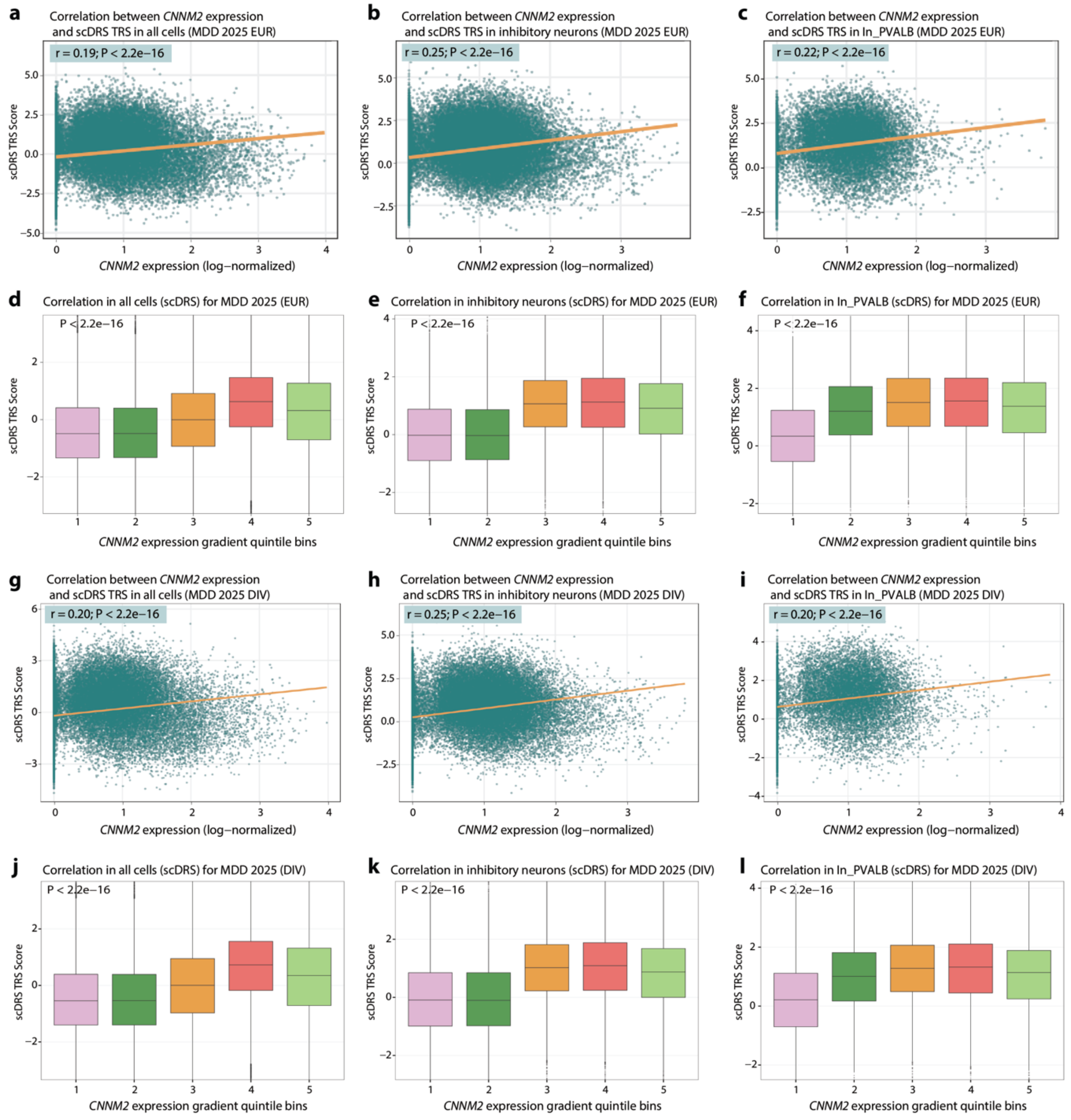


**Supplementary Figure 36 | Replication of *CNNM2* expression–MDD genetic relevance associations using updated PGC GWAS datasets, related to Figure 5.**

a–c, Scatter plots showing the relationship between *CNNM2* expression and scDRS disease-relevance scores derived from the PGC 2025 European-ancestry MDD GWAS (EUR) across all cells (a), inhibitory neurons (b) and PVALB$^+$ inhibitory neurons (c).

d–f, Distribution of scDRS disease-relevance scores across quintiles of *CNNM2* expression in all cells (d), inhibitory neurons (e) and PVALB$^+$ inhibitory neurons (f) using the PGC 2025 European-ancestry GWAS.

g–i, Scatter plots showing the relationship between CNNM2 expression and scDRS disease-relevance scores derived from the PGC 2025 diverse/trans-ancestry MDD GWAS (DIV) across all cells (g), inhibitory neurons (h) and PVALB$^+$ inhibitory neurons (i).

j–l, Distribution of scDRS disease-relevance scores across quintiles of *CNNM2* expression in all cells (j), inhibitory neurons (k) and PVALB$^+$ inhibitory neurons (l) using the PGC 2025 diverse/trans-ancestry GWAS. Pearson correlation coefficients and P values are shown for scatter plots.

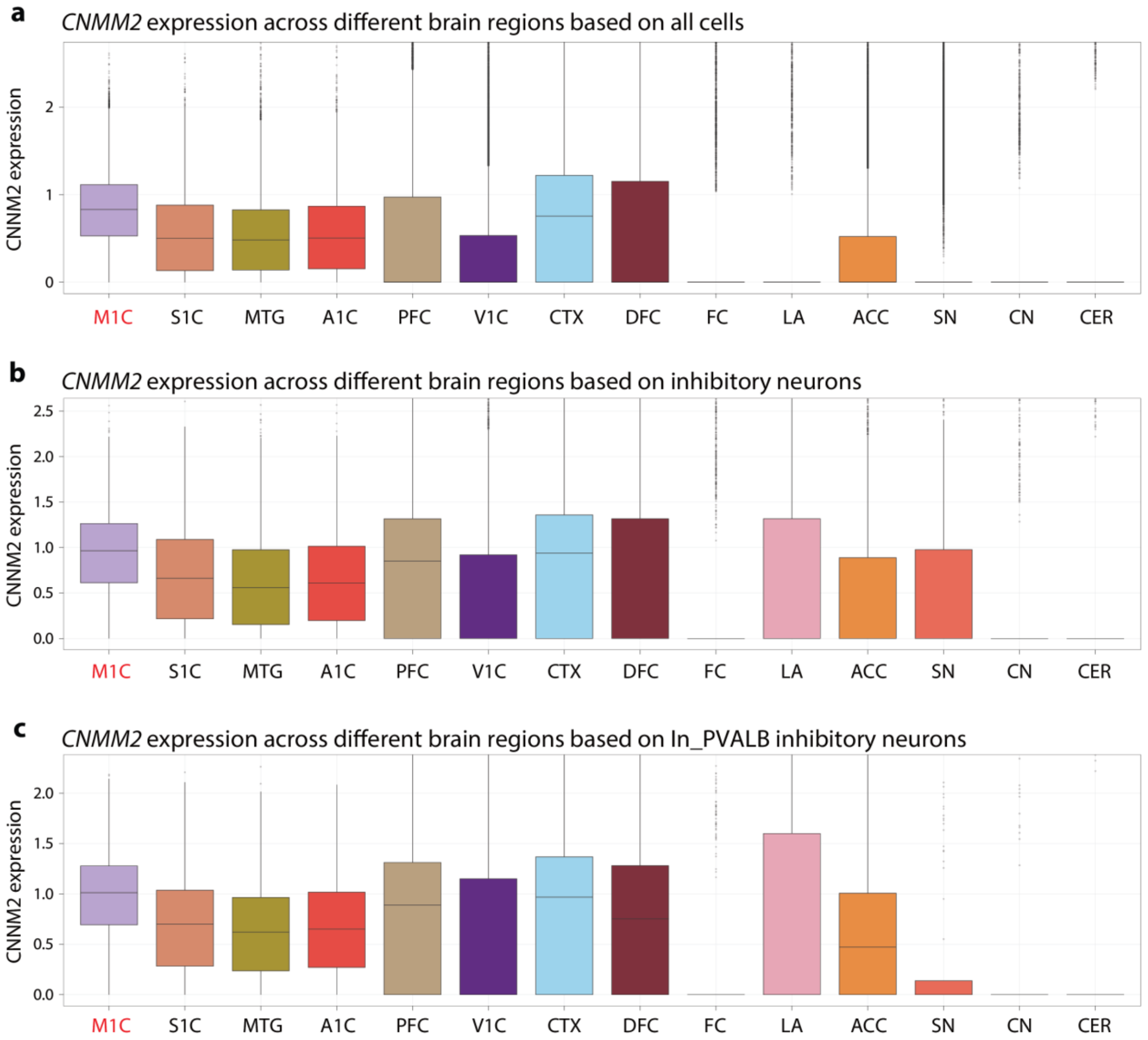


**Supplementary Figure 37 | Regional distribution of *CNMM2* expression across MDD-relevant neuronal strata, related to Figure 5.**

a–c, Box plots showing log-normalized *CNNM2* expression across 14 anatomical brain regions in the integrated human brain discovery atlas, shown for all cells (a; n = 310,833 cells from 109 donors), inhibitory neurons (b; n = 57,453 cells) and PVALB$^+$ inhibitory neurons (c; n = 16,363 cells). Regions are ordered according to the regional MDD trait-relevance hierarchy shown in Fig. X; M1C is highlighted in red. *CNNM2* expression was consistently elevated in M1C across all three strata, including within PVALB$^+$ inhibitory neurons. Box plots show the median, interquartile range and 1.5 × interquartile range; grey points denote outlier cells. Statistical comparisons were performed using one-sided Wilcoxon rank-sum tests comparing M1C with all other regions combined.

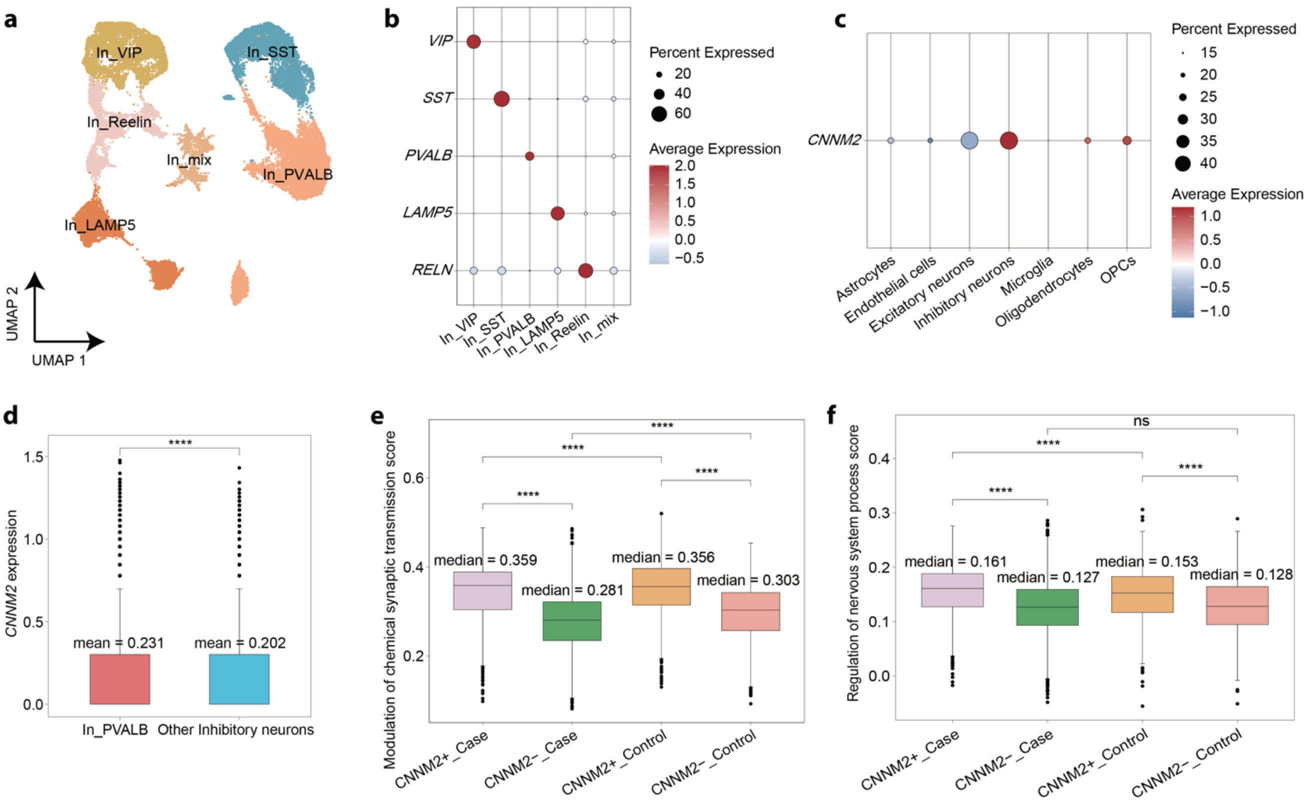


**Supplementary Figure 38 | Replication of the *CNNM2*⁺ PVALB⁺ inhibitory-neuron program in an independent human MDD case–control dataset, related to Figure 5.**

a. UMAP visualization of re-annotated inhibitory-neuron subtypes in the independent human MDD case–control dataset, showing VIP⁺, SST⁺, PVALB⁺, LAMP5⁺, RELN⁺ and mixed inhibitory-neuron populations.

b. Dot plot showing canonical marker-gene expression across inhibitory-neuron subtypes. Dot size indicates the percentage of cells expressing each gene, and colour indicates scaled average expression.

c. Dot plot showing *CNNM2* expression across major brain cell classes in the replication dataset. Dot size indicates the percentage of expressing cells, and colour indicates scaled average expression.

d. Box plot comparing *CNNM2* expression between PVALB⁺ inhibitory neurons and other inhibitory-neuron subtypes.

e,f. Module scores for *CNNM2*⁺ PVALB⁺ pathway signatures across neurons stratified by diagnosis and *CNNM2* expression status: *CNNM2*⁺ MDD, *CNNM2*⁻ MDD, *CNNM2*⁺ control and *CNNM2*⁻ control neurons. Pathway signatures include modulation of chemical synaptic transmission (e) and regulation of nervous system process (f). Box plots show median and interquartile range; whiskers indicate 1.5× interquartile range. Statistical significance was assessed using two-sided Wilcoxon rank-sum tests. **P < 0.0001; ns, not significant.

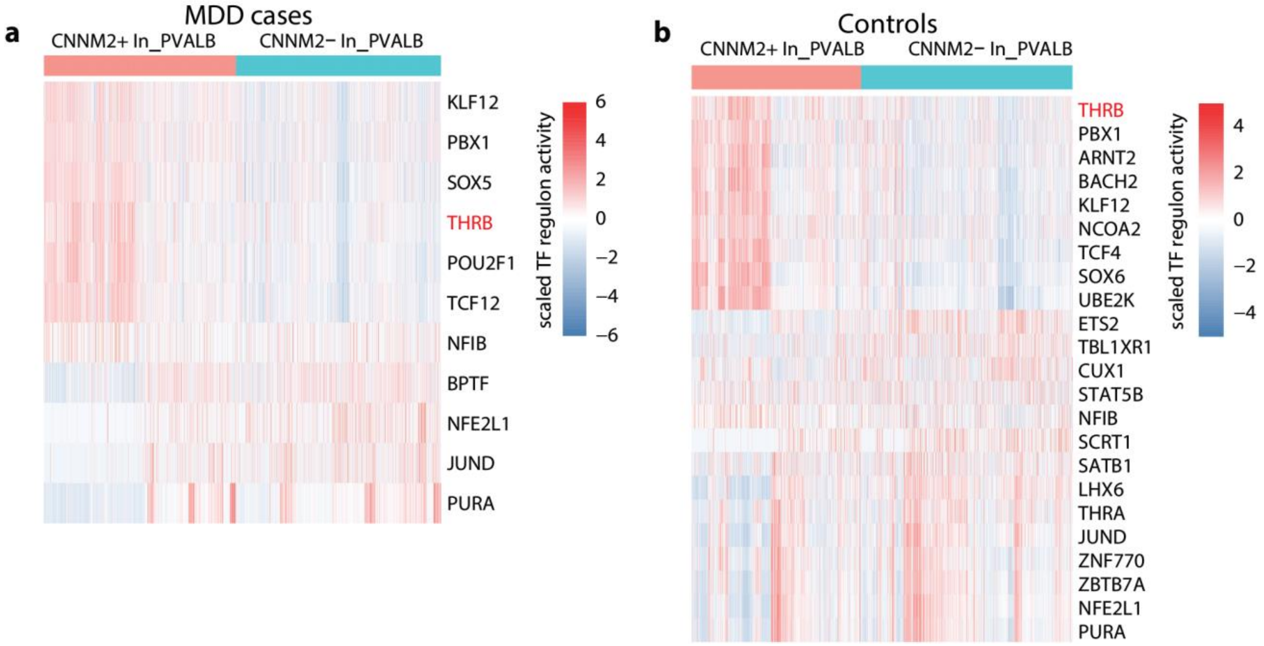


**Supplementary Figure 39 | THRB regulon activity in CNNM2⁺ PVALB⁺ inhibitory neurons in an independent human MDD case–control dataset, related to Figure 5.**

a,b. Heatmaps showing pySCENIC-inferred transcription factor regulon activity in *CNNM2*⁺ and *CNNM2*⁻ PVALB⁺ inhibitory neurons from MDD cases (a) and controls (b). Columns represent single cells grouped by *CNNM2* expression status, and rows represent transcription factor regulons. Colour indicates scaled regulon activity. THRB regulon activity was elevated in *CNNM2*⁺ PVALB⁺ neurons in both MDD cases and controls, consistent with the regulatory pattern observed in the discovery atlas.

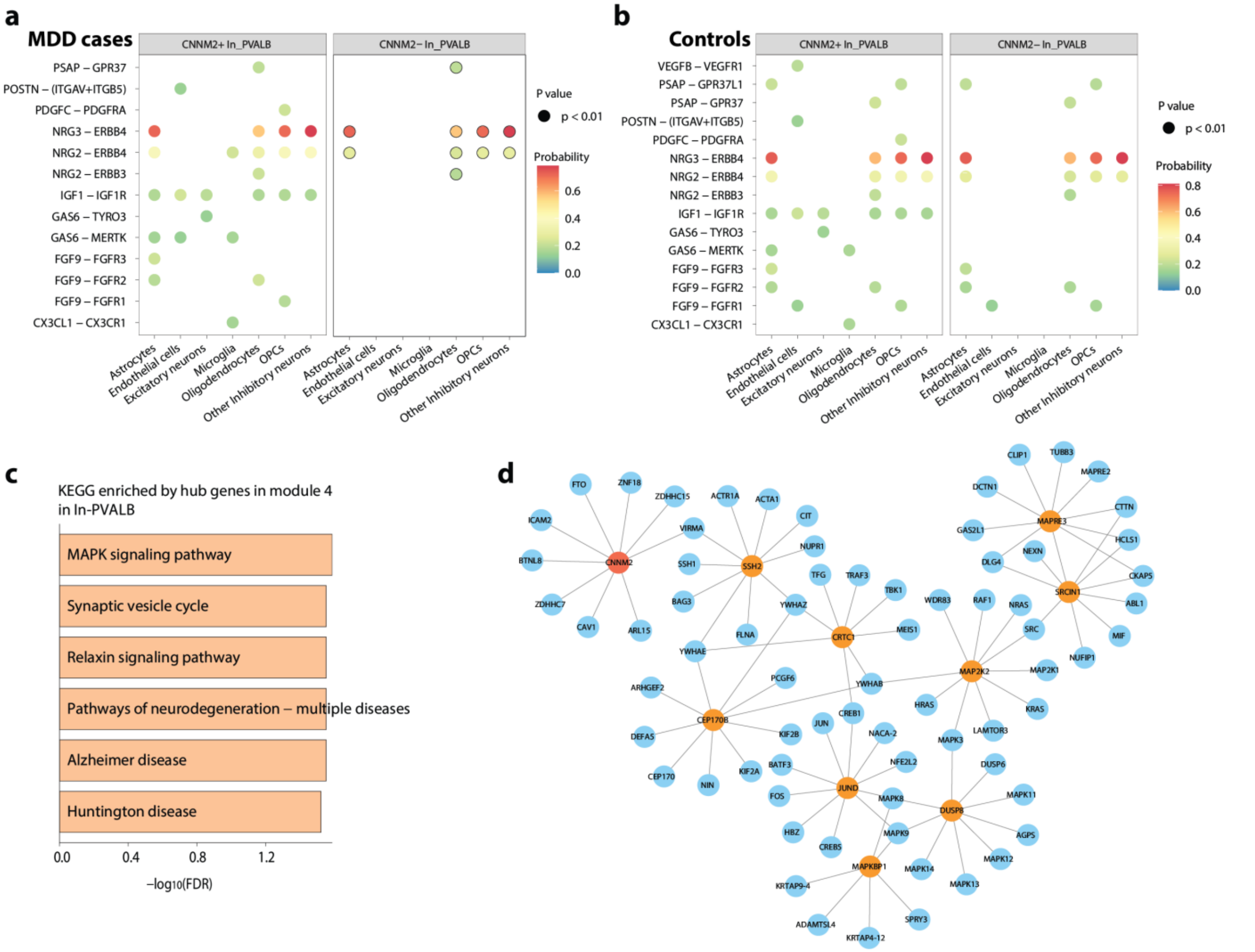


**Supplementary Figure 40 | Replication and network characterization of the CNNM2⁺ PVALB⁺ inhibitory-neuron program, related to Figure 5.**

a,b. Dot plots showing CellChat-inferred ligand–receptor interactions involving *CNNM2*⁺ and *CNNM2*⁻ PVALB⁺ inhibitory neurons in MDD cases (a) and controls (b) from the independent human MDD case–control dataset. Columns indicate interacting cell populations, rows indicate ligand–receptor pairs, dot colour denotes inferred communication probability and dot size indicates statistical significance; only significant interactions are shown ($P < 0.01$).

c. KEGG pathway enrichment of hub genes from the *CNNM2*-containing co-expression module identified by hdWGCNA in PVALB⁺ inhibitory neurons. Bar length indicates $-\log_{10}$(FDR).

d. STRING-based protein–protein interaction network linking *CNNM2* with hub genes from the *CNNM2*-associated co-expression module. The red node denotes *CNNM2*, orange nodes denote MAPK-related hub genes and blue nodes denote interacting module genes. Edges indicate protein–protein interactions inferred from STRING.

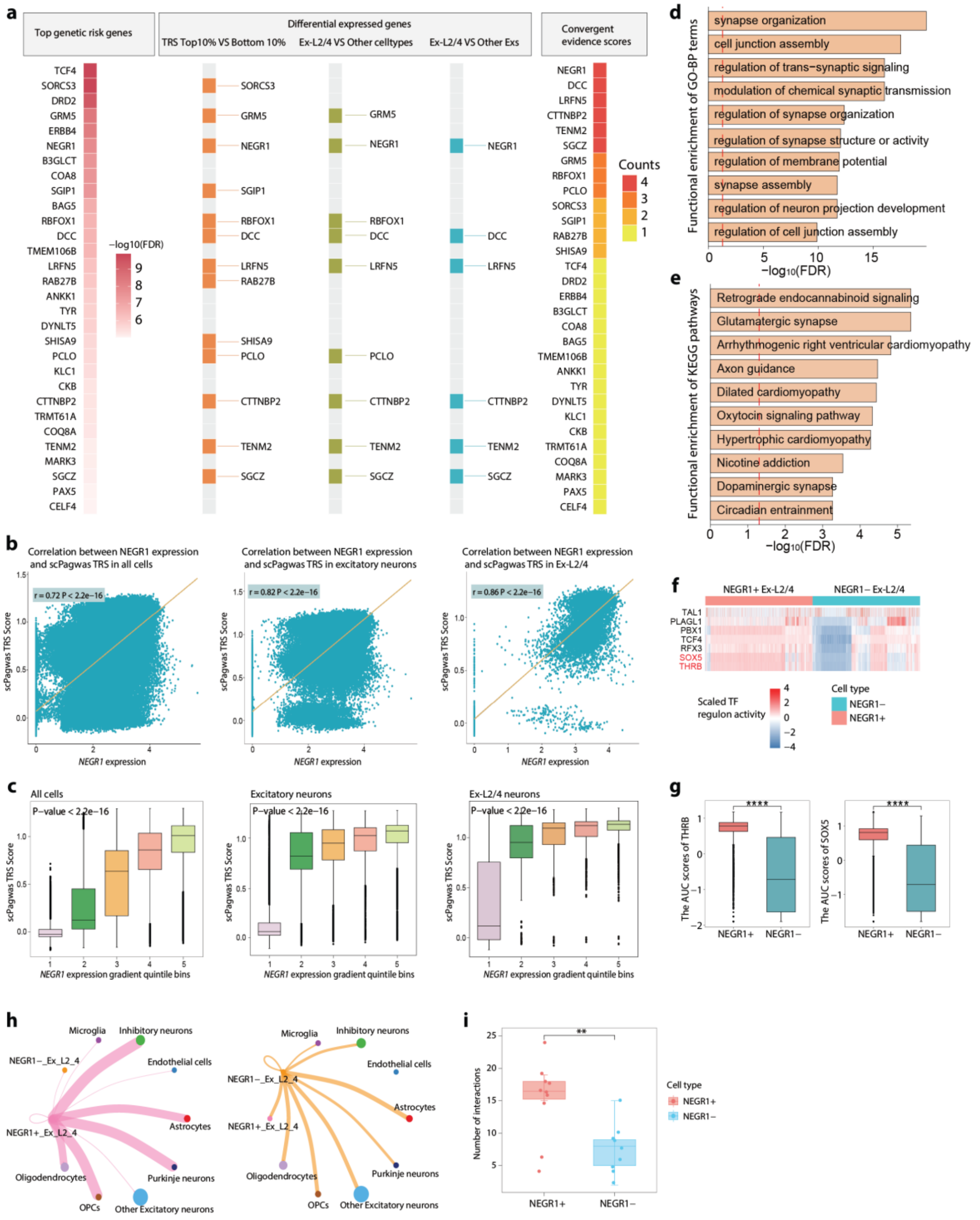


### Supplementary Figure 41 | Integrative prioritization and molecular characterization of the *NEGR1*-linked Ex-L2/4 excitatory-neuron program, related to Figure 5.

a. Integrative prioritization of candidate MDD risk genes in Ex-L2/4 excitatory neurons. Evidence layers included top MAGMA gene-wise association signals, differential expression between cells with top and bottom 10% scPagwas trait-relevance scores (TRSs), differential expression between Ex-L2/4 neurons and other major brain cell classes, and differential expression between Ex-L2/4 neurons and other excitatory-neuron subtypes. The right panel summarizes convergent evidence scores across methods.
b. Correlation between *NEGR1* expression and single-cell scPagwas TRSs across all cells, excitatory neurons and Ex-L2/4 neurons.
c. Distribution of scPagwas TRSs across *NEGR1* expression quintile bins in all cells, excitatory neurons and Ex-L2/4 neurons.
d,e. Functional enrichment analysis comparing *NEGR1*$^+$ and *NEGR1*$^-$ Ex-L2/4 neurons, showing enriched Gene Ontology biological process terms (d) and KEGG pathways (e). Bar length indicates $-\log_{10}$(FDR).
f. Heatmap of transcription factor regulon activities inferred by pySCENIC in *NEGR1*$^+$ and *NEGR1*$^-$ Ex-L2/4 neurons.
g. Box plots showing regulon activity scores for representative transcription factors, including TCF4 and THRB, in *NEGR1*$^+$ versus *NEGR1*$^-$ Ex-L2/4 neurons.
h. CellChat-inferred intercellular communication networks involving *NEGR1*$^+$ and *NEGR1*$^-$ Ex-L2/4 neurons and other annotated brain cell populations.
i. Quantification of the number of predicted ligand–receptor interactions between *NEGR1*$^+$ or *NEGR1*$^-$ Ex-L2/4 neurons and other brain cell populations. Box plots show median and interquartile range; whiskers indicate 1.5× interquartile range. Statistical significance was assessed using two-sided Wilcoxon rank-sum tests unless otherwise indicated. $P < 0.05$, P < 0.01, **P < 0.0001.

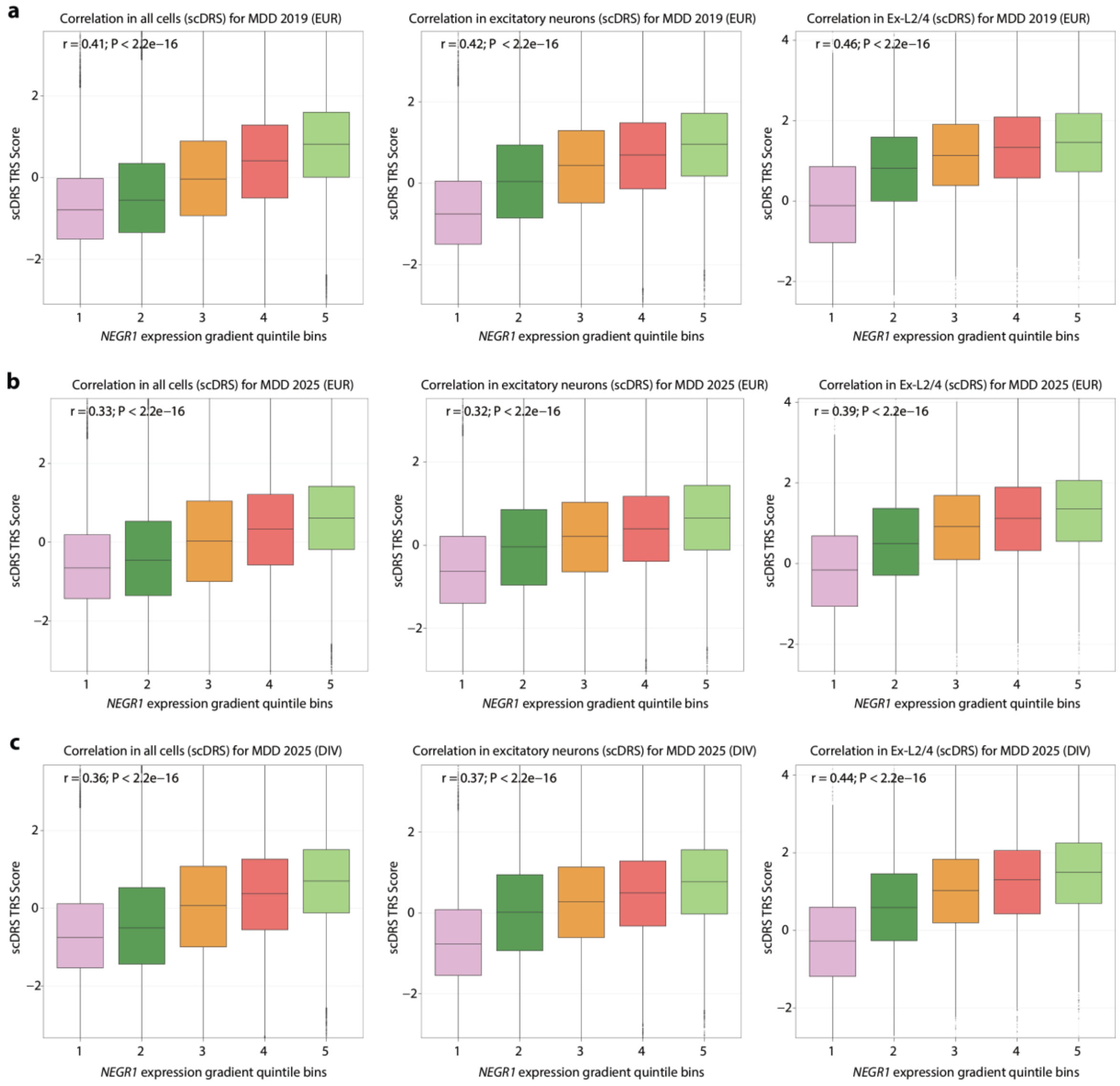


**Supplementary Figure 42 | *NEGR1* expression is associated with scDRS MDD disease-relevance scores across GWAS resources, related to Figure 5.**
a–c, Box plots showing scDRS disease-relevance scores across quintiles of *NEGR1* expression for the primary Howard et al. European-ancestry GWAS (a), the PGC 2025 European-ancestry GWAS (b) and the PGC 2025 trans-ancestry (DIV) GWAS (c). Within each GWAS, analyses are shown for all cells, excitatory neurons and Ex-L2/4 excitatory neurons. *NEGR1* expression showed consistent positive associations with scDRS disease-relevance scores across these cell strata and GWAS resources, with the largest correlations observed in Ex-L2/4 excitatory neurons. Pearson correlation coefficients and P values are shown in each panel. Box plots show the median, interquartile range and 1.5 × interquartile range.

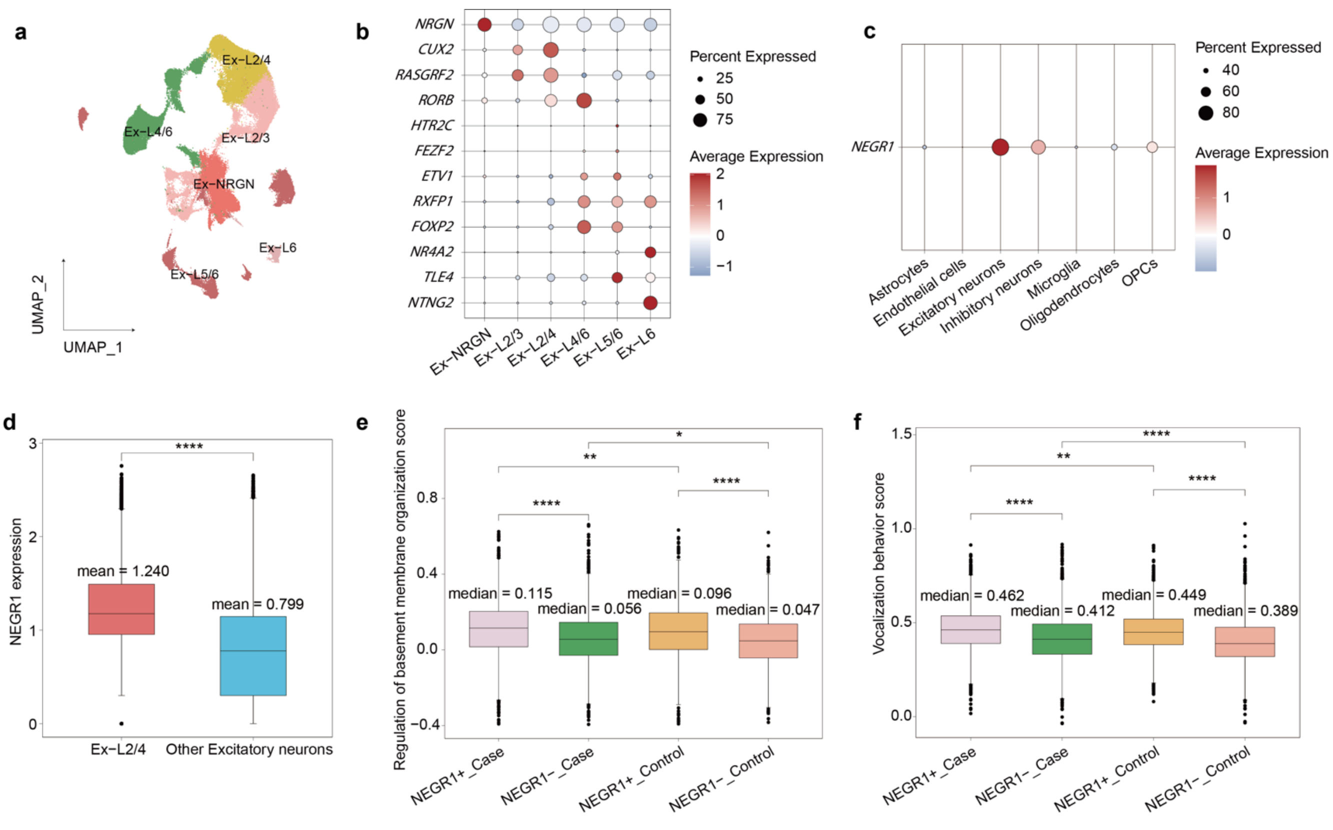


**Supplementary Figure 43 | Replication of the *NEGR1*-associated Ex-L2/4 excitatory-neuron program in an independent human MDD case–control dataset, related to Figure 5.**

a. UMAP visualization of excitatory-neuron subtypes after re-annotation in the independent MDD case–control dataset, showing Ex-L2/4, Ex-L2/3, Ex-L4/6, Ex-L5/6, Ex-L6 and Ex-NRGN populations.

b. Dot plot showing canonical marker-gene expression across excitatory-neuron subtypes. Dot size denotes the percentage of cells expressing each gene, and colour indicates scaled average expression.

c. Dot plot showing *NEGR1* expression across major brain cell classes in the replication dataset.

d. Box plot comparing *NEGR1* expression between Ex-L2/4 neurons and other excitatory-neuron subtypes.

e,f. Module scores for Ex-L2/4-associated pathway signatures across cells stratified by diagnosis and *NEGR1* expression status: *NEGR1*$^{+}$ MDD, *NEGR1*$^{-}$ MDD, *NEGR1*$^{+}$ control and *NEGR1*$^{-}$ control cells. Pathway signatures include regulation of basement membrane organization (e) and vesicle-mediated transport (f). Box plots show median and interquartile range; whiskers indicate 1.5× interquartile range. Statistical significance was assessed by a two-sided Wilcoxon rank-sum test. P < 0.05, P < 0.01, **P < 0.0001.

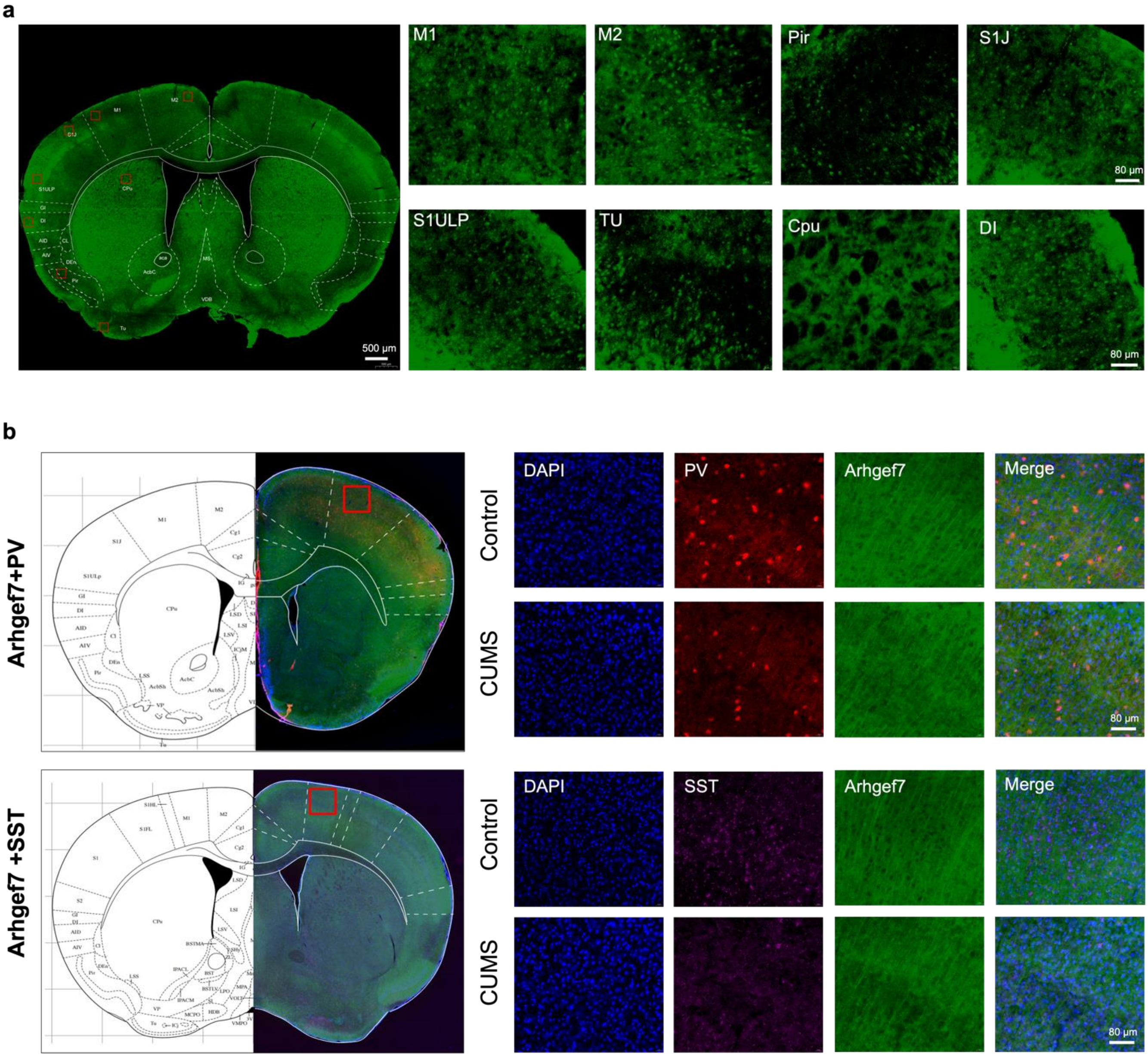


**Supplementary Figure 44 | Regional Cnnm2 expression and Arhgef7 validation in motor-cortical inhibitory neurons after chronic stress, related to Figure 6.**

a. Immunofluorescence detection of Cnnm2 protein signals distribution in the mouse whole brain. Left: Representative image of the whole-brain section (Scale bar, 500 μm). Right: Magnified images showing the expression of Cnnm2 in the primary motor cortex (M1), secondary motor cortex (M2), piriform cortex (Pir), primary somatosensory cortex jaw region (S1J), primary somatosensory cortex upper lip region (S1ULP), olfactory tubercle (TU), caudate putamen (CPu), and substantia innominata (DI) (Scale bar, 80 μm).

b. Expression of Arhgef7 in PVALB$^{+}$ (PV$^{+}$) and SST$^{+}$ inhibitory neurons. Left: Representative whole brain section indicating the immunofluorescent staining of Arhgef7 and PV$^{+}$/SST$^{+}$ neurons. Right: Representative immunofluorescent images of PV$^{+}$ neurons, Arhgef7$^{+}$ neurons, and their colocalization in M1 region in both Control and CUMS groups. Scale bar, 80 μm.

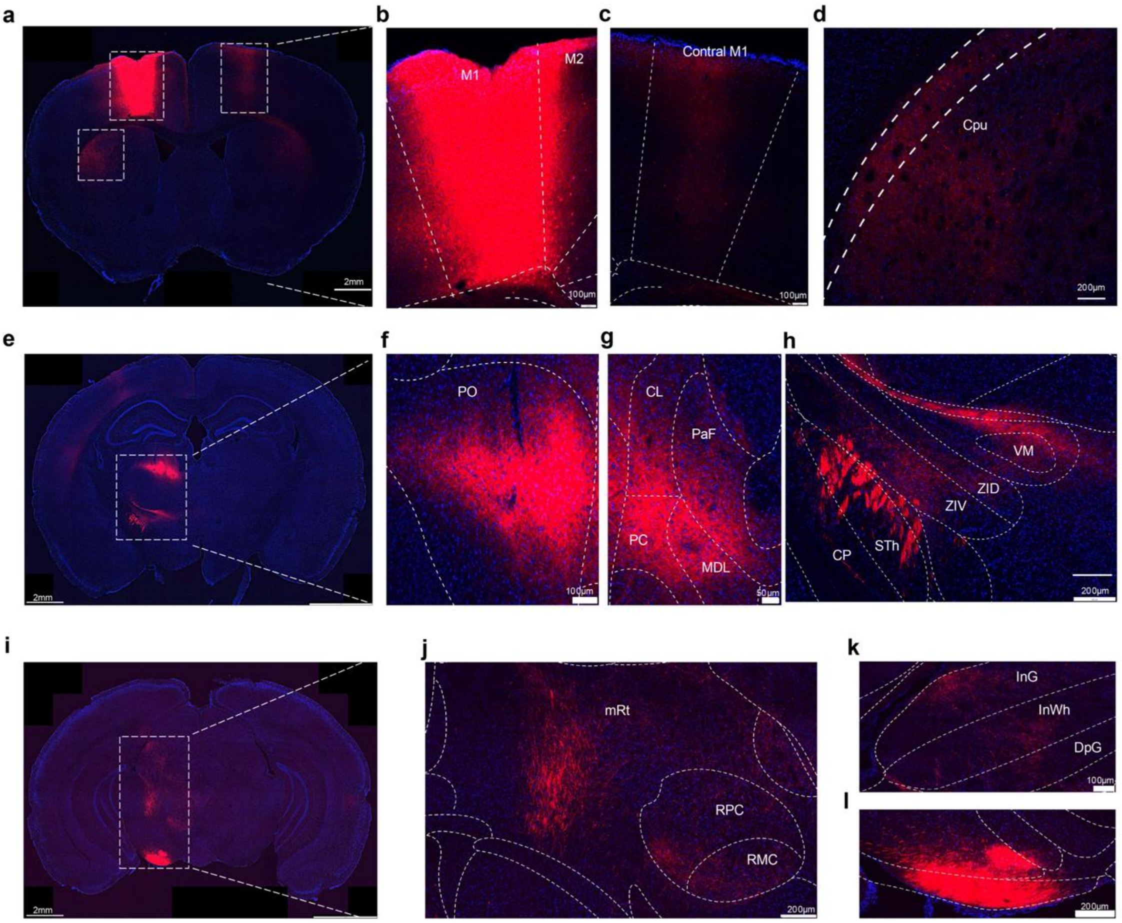


**Supplementary Figure 45 | Brain regions receiving direct projections from the primary motor cortex (M1), related to Figure 7.**

a. Three weeks after AAV2/9-hSyn-mCherry was injected into the M1, immunofluorescence staining detected mCherry-labeled nerve fibers in the secondary motor cortex (M2), contralateral M1 and caudate putamen (Cpu).

b-d. Magnified images of mCherry-positive regions in panel a.

e. For another coronal brain slice, mCherry-positive fibers were observed in the posterior thalamic nucleus (PO), centro-lateral thalamic nucleus (CL), parafascicular thalamic nucleus (PaF), paracentral thalamic nucleus (PC), medio-dorsal thalamic nucleus (MDL), ventro-medial thalamic nucleus (VM), dorsal zona incerta (ZID), ventral zona incerta (ZIV), subthalamic nucleus (STh), and cerebral peduncle (CP).

f-l. Magnified images of mCherry-positive regions in panel e.

j. mCherry-positive fibers were also detected in the intermediate gray layer of the superior colliculus (InG), intermediate white layer of the superior colliculus (InWh), deep gray layer of the superior colliculus (DpG), mesencephalic reticular nucleus (mRt), red nucleus (RN), and cerebral peduncle (CP).

k-l. Magnified images of mCherry-positive regions in panel j.

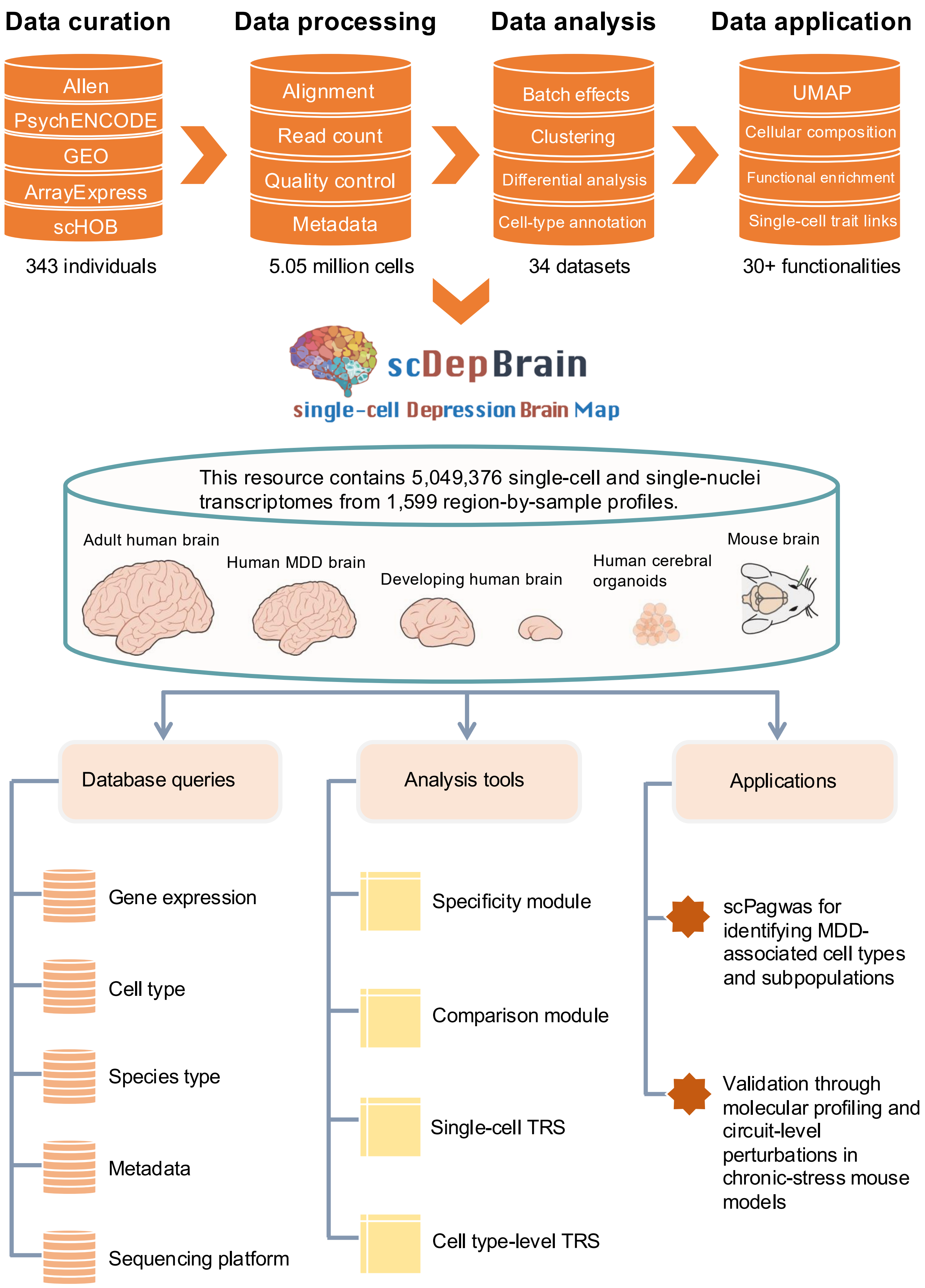


**Supplementary Figure 46 | See next page for caption.**

**Supplementary Figure 46 | Workflow and functional modules of scDepBrain for exploring MDD-associated brain cellular programs, related to Figure 1.**
Schematic overview of scDepBrain construction and application. Public brain single-cell and single-nucleus transcriptomic datasets were collected from multiple resources, including Allen Brain Map, PsychENCODE, GEO, ArrayExpress and scHOB. The collected datasets were processed through a standardized workflow, including count-matrix collection, metadata harmonization, quality control, batch-effect correction, clustering, differential-expression analysis and cell-type annotation. Processed datasets and precomputed results were integrated into scDepBrain, an interactive single-cell Depression Brain Map comprising 5,049,376 single-cell and single-nucleus transcriptomes from 1,599 region-by-sample profiles across adult human brain, human MDD brain, developing human brain, human cerebral organoid and mouse brain datasets. scDepBrain provides three major functional components: database queries, analysis tools and application modules. Database query functions allow users to browse datasets, metadata, cell types, genes and downloadable resources. Analysis modules support UMAP visualization, gene-expression exploration, cellular-composition comparison, marker-gene inspection, differential-expression analysis, pathway enrichment and MDD genetic-prioritization results. Application modules enable cross-dataset exploration of MDD-associated cell types, trait-relevance scores, candidate genes and cellular programs, providing a reusable platform for hypothesis generation and experimental follow-up in MDD research.

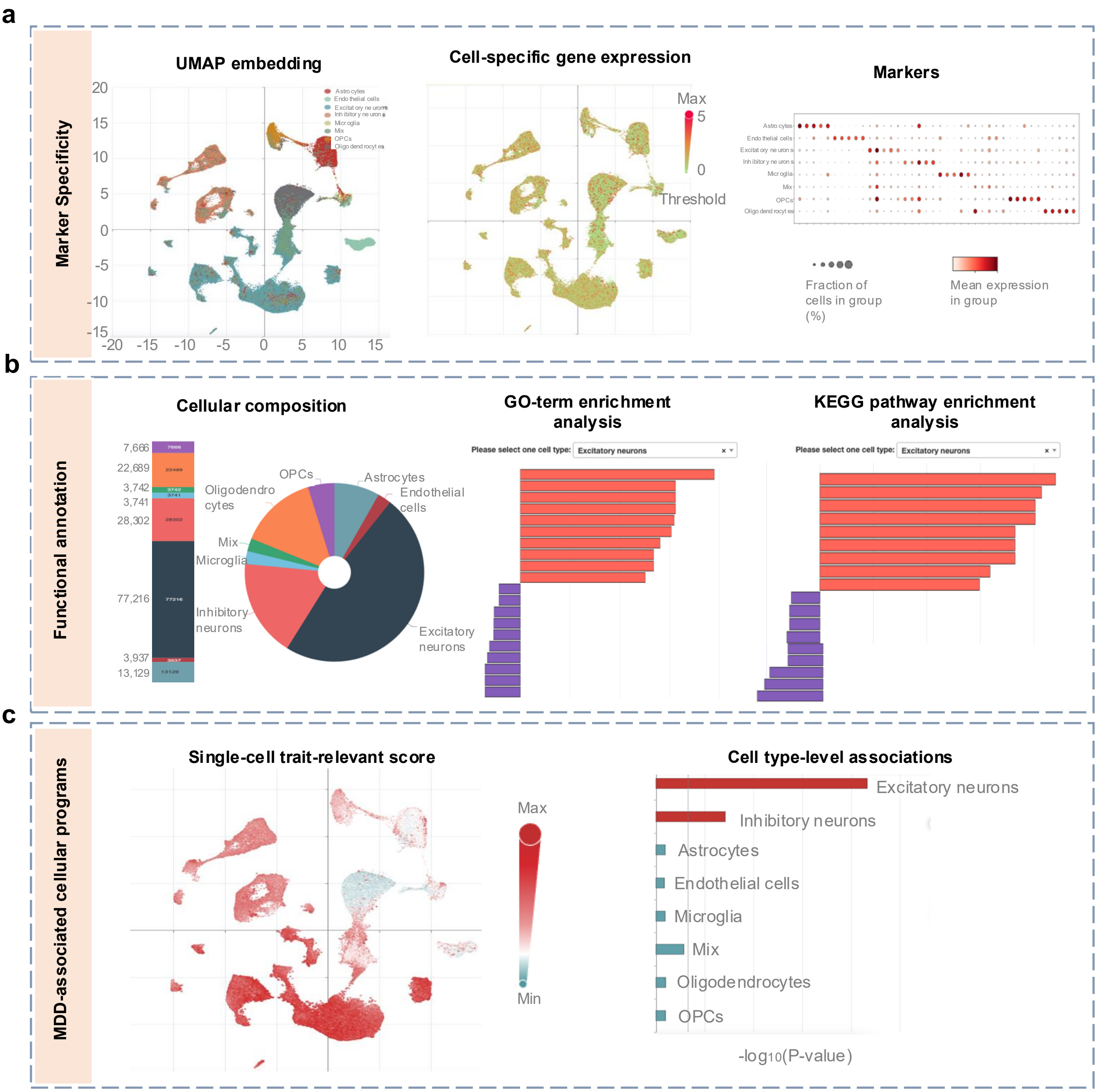


**Supplementary Figure 47 | Screen captures of online functional modules in scDepBrain, related to Figure 1.**

a. Marker specificity module. This module enables users to explore cell annotations and marker-gene expression within each selected brain single-cell or single-nucleus dataset. It includes UMAP visualization of annotated cell populations, UMAP projection of user-selected gene expression and dot-plot visualization of marker genes across cell types or subtypes. Dot size indicates the percentage of cells expressing each gene and colour indicates average expression.

b. Functional annotation module. This module provides dataset-level summaries and downstream annotation results, including cellular-composition visualization, cell-type-specific differential-expression results and GO/KEGG pathway-enrichment analysis. Users can select datasets, cell types or genes of interest to compare cellular composition, identify marker or differentially expressed genes and explore enriched biological pathways.

c. MDD-associated cellular program module. This module displays genetic-prioritization results derived from integration of MDD GWAS with single-cell transcriptomic data, including single-cell trait-relevance score projections on UMAP and cell-type-level MDD association results. Together, these modules allow users to query MDD-associated cell types, genes, pathways and trait-relevance patterns across discovery and replication brain datasets.
